# Dexamethasone 12 mg versus 6 mg for patients with COVID-19 and severe hypoxia: an international, randomized, blinded trial

**DOI:** 10.1101/2021.07.22.21260755

**Authors:** Marie W. Munch, Sheila N. Myatra, Bharath Kumar Tirupakuzhi Vijayaraghavan, Sanjith Saseedharan, Thomas Benfield, Rebecka R. Wahlin, Bodil S. Rasmussen, Anne Sofie Andreasen, Lone M. Poulsen, Luca Cioccari, Mohd S. Khan, Farhad Kapadia, Jigeeshu V. Divatia, Anne C. Brøchner, Morten H. Bestle, Marie Helleberg, Jens Michelsen, Ajay Padmanaban, Neeta Bose, Anders Møller, Kapil Borawake, Klaus T. Kristiansen, Urvi Shukla, Michelle S. Chew, Subhal Dixit, Charlotte S. Ulrik, Pravin R. Amin, Rajesh Chawla, Christian A. Wamberg, Mehul S. Shah, Iben S. Darfelt, Vibeke L. Jørgensen, Margit Smitt, Anders Granholm, Maj-Brit N. Kjær, Morten H. Møller, Tine S. Meyhoff, Gitte K. Vesterlund, Naomi E. Hammond, Sharon Micallef, Abhinav Bassi, Oommen John, Anubhuti Jha, Maria Cronhjort, Stephan M. Jakob, Christian Gluud, Theis Lange, Vaijayanti Kadam, Klaus V. Marcussen, Jacob Hollenberg, Anders Hedman, Henrik Nielsen, Olav L. Schjørring, Marie Q. Jensen, Jens W. Leistner, Trine B. Jonassen, Camilla M. Kristensen, Esben C. Clapp, Carl J. S. Hjortsø, Thomas S. Jensen, Liv S. Halstad, Emilie R. B. Bak, Reem Zaabalawi, Matias Metcalf-Clausen, Suhayb Abdi, Emma V. Hatley, Tobias S. Aksnes, Emil Gleipner-Andersen, Arif F. Alarcón, Gabriel Yamin, Adam Heymowski, Anton Berggren, Kirstine la Cour, Sarah Weihe, Alison H. Pind, Janus Engstrøm, Vivekanand Jha, Balasubramanian Venkatesh, Anders Perner, COVID STEROID 2 trial collaborators

## Abstract

**IMPORTANCE:** Dexamethasone 6 mg daily is recommended for up to 10 days in patients with severe and critical COVID-19, but a higher dose may benefit those with more severe disease.

**OBJECTIVE:** To assess the effects of dexamethasone 12 mg versus 6 mg daily in patients with COVID-19 and severe hypoxia.

**DESIGN, SETTING, PARTICIPANTS:** We conducted a parallel-grouped, stratified, blinded randomized trial including 1000 adults with confirmed COVID-19 receiving at least 10 l/min of oxygen or mechanical ventilation in 26 hospitals in Europe and India from August 2020 to May 2021.

**INTERVENTIONS:** Patients were randomized 1:1 to masked intravenous dexamethasone 12 mg or dexamethasone 6 mg daily for up to 10 days.

**MAIN OUTCOME MEASURES:** The primary outcome was the number of days alive without life support (i.e. invasive mechanical ventilation, circulatory support or renal replacement therapy) at 28 days. The primary analysis was adjusted for stratification variables (site, age below 70 years and invasive mechanical ventilation).

**RESULTS:** After exclusion of 18 patients who withdrew consent, there were 982 patients (613 in Europe, 369 in India) in the intention-to-treat population. We had primary outcome data for 971 patients; 491 assigned to dexamethasone 12 mg and 480 assigned to dexamethasone 6 mg. The median number of days alive without life support was 22.0 days (interquartile range 6.0-28.0) in the 12 mg group and 20.5 days (4.0-28.0) in the 6 mg group (adjusted mean difference 1.3 days, 95% confidence interval (CI), 0.0-2.6; P=0.066). Mortality at 28 days was 27.1% and 32.3% in patients assigned to 12 mg and 6 mg, respectively (adjusted relative risk 0.86, 99% CI, 0.68-1.08). Serious adverse reactions, including septic shock and invasive fungal infections, occurred in 11.3% versus 13.2% of patients in the 12 mg group and 6 mg group, respectively (adjusted relative risk 0.85, 99% CI, 0.55-1.32).

**CONCLUSIONS AND RELEVANCE:** Among patients with COVID-19 and severe hypoxia, dexamethasone 12 mg did not result in statistically significantly more days alive without life support at 28 days than dexamethasone 6 mg. However, the confidence interval around the point estimate should be considered when interpreting the results of this trial.

**TRIAL REGISTRATION:** ClinicalTrials.gov (NCT04509973) and Clinical Trial Registry India (2020/10/028731).

---

Patients with critical COVID-19 are characterized by severe pulmonary inflammation and hypoxia, which often leads to use of high flow oxygen, mechanical ventilation and, in case of further disease progression, circulatory support and renal replacement therapy.^1^

Dexamethasone is recommended by the World Health Organization for patients with severe and critical COVID-19^2^ based on a prospective meta-analysis of 7 randomized trials reporting reduced short-term mortality with the use of systemic corticosteroids.^3^ The largest of these trials, the Randomised Evaluation of COVID-19 Therapy (RECOVERY) trial, demonstrated a mortality benefit with 6 mg of dexamethasone per day for up to 10 days.^4^ Among the remaining 6 trials in the meta-analysis,^3^ most evaluated daily doses of corticosteroids that were above 6 mg of dexamethasone (median dose in dexamethasone equivalents 12 mg, range 6 to 16 mg).^5-8^ Higher doses of dexamethasone have also been reported beneficial in a randomized trial in non-COVID-19 patients with acute respiratory distress syndrome.^9^ Pharmacodynamic studies suggest dose-dependent activation of the corticosteroid receptor with increasing doses up to 60 mg of prednisone (equivalent to 12 mg of dexamethasone).^10^

Taken together, higher doses of dexamethasone than the recommended 6 mg daily may benefit patients with COVID-19 who have more severe disease. However, there are concerns about adverse reactions with the use of higher doses of corticosteroids,^11^ particularly reports of severe fungal infections, such as mucormycosis, in patients with COVID-19 treated with corticosteroids.^12,13^

We conducted the COVID STEROID 2 trial to evaluate the efficacy and safety of higher dose dexamethasone in hospitalized adults with COVID-19 and severe hypoxia. We hypothesized that a higher daily dose of dexamethasone (12 mg) as compared to the currently recommended daily dose (6 mg) would increase the number of days alive without life support at 28 days in these patients.

## Methods

### Trial design and oversight

The COVID STEROID 2 trial was an investigator-initiated, international, parallel-grouped, stratified, blinded randomized clinical trial. The trial protocol was approved by the Ethics Committee of the Capital Region of Denmark and institutionally at each trial site. Before enrolment was completed, we published the trial protocol and statistical analysis plan,^14^ which is available in Supplement 1. Trial conduct was overseen by the Collaboration for Research in Intensive Care, Copenhagen, Denmark, and for the Indian sites by the George Institute for Global Health, New Delhi, India and Sydney, Australia. A data safety monitoring committee (DSMC) oversaw the safety of the trial participants and conducted one planned interim analysis.

### Trial sites and patients

Patients underwent screening and randomization between August 27, 2020 and May 20, 2021 at 31 sites in 26 hospitals (11 in Denmark, 12 in India, 2 in Sweden, and 1 in Switzerland). In 2 of the Danish hospitals there were multiple sites at ICUs and departments of infectious diseases and pulmonary medicine.

Eligible patients were 18 years or older, hospitalized with confirmed SARS-CoV-2 infection on (i) supplementary oxygen at a flow of at least 10 l/min independent of delivery system, (ii) non-invasive ventilation or continuous positive airway pressure for hypoxia or (iii) invasive mechanical ventilation. We excluded patients who used systemic corticosteroids for other indications than COVID-19 in doses higher than 6 mg dexamethasone equivalents or had used systemic corticosteroids for COVID-19 for 5 days or more, those with invasive fungal infection or active tuberculosis, those with known hypersensitivity to dexamethasone and those who were pregnant. The full definitions of inclusion and exclusion criteria are presented in eMethods in Supplement 2.

### Consent procedure

We obtained informed consent from the patients or their legal surrogates according to national regulations. In many institutions, enrolment was allowed as an emergency procedure (e.g. consent from a doctor who was independent of the trial) followed by consent from relatives and the patient to continue participation. If consent was withdrawn or not granted, permission was sought from the patient or relatives to continue collection and use of trial data.

### Randomization, masking and interventions

Randomization was performed with a centralized, computer-generated allocation sequence stratified by trial site, age below 70 years, and use of invasive mechanical ventilation at the time of screening. Eligible patients were randomly allocated in a 1:1 ratio to dexamethasone 12 mg or 6 mg daily using permuted blocks of varying sizes. Treatment assignments were concealed from patients, clinicians, investigators, trial statisticians, the DSMC and the Management Committee when they wrote the first version of the abstract (eMethods in Supplement 2).

Dexamethasone 12 mg (as dexamethasone phosphate 14.4 mg) or 6 mg (as dexamethasone phosphate 7.2 mg) was suspended in sodium chloride 0.9% (eFigure 1 in Supplement 2) and administered as a masked bolus injection (total volume 5 ml) intravenously once daily for up to 10 days from randomization. We allowed the use of betamethasone at sites where dexamethasone was not available (one hospital in Sweden), as these are diastereomers and likely equipotent.^15^ A team of unblinded trial staff, who were not involved in the care of trial patients, the entry of outcome data or the statistical analysis, prepared the masked trial medication from shelf medication at local hospital pharmacies (the trade names are presented in eFigure 1 in Supplement 2). They were instructed not to reveal the treatment allocation unless the participant was subject to emergency unblinding. If the patient had used dexamethasone for COVID-19 prior to enrolment, the intervention period was shortened so that no patients received dexamethasone for more than 10 days per protocol. All other interventions were at the discretion of the clinicians, except that we recommended against the use of other immunosuppressive agents for COVID-19. From January 9 2021, we accepted the use of tocilizumab following the preprint publication of the interleukin-6 receptor antagonist (IL-6-RA) domain of the Randomized, Embedded, Multifactorial Adaptive Platform Trial for Community Acquired Pneumonia (REMAP-CAP) trial.^16^

**Figure 1.**
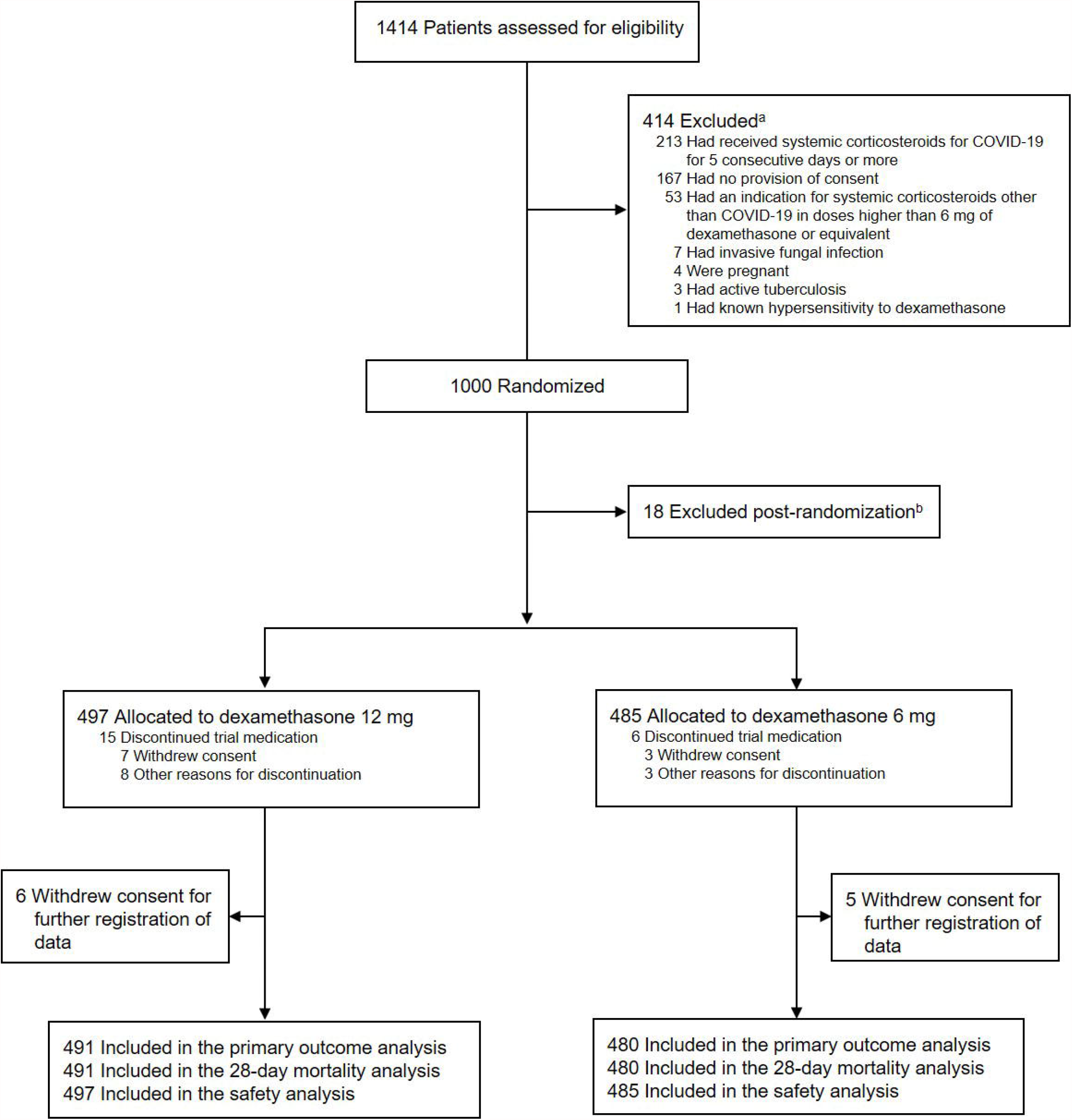
Screening, Randomization and Follow-up of Patients in the COVID STEROID 2 trial. ^a^33 patients met more than 1 criterion. ^b^13 patients were excluded before the first dose of trial medication was administered because consent was withdrawn. Another 5 patients were excluded as required by the Swiss Ethics Committee during the intervention period because consent was not granted. Thus 18 patients in total were not included in any analyses because of lack of consent for the use of data.

### Data collection and monitoring

The trial investigators or their delegates reported any serious adverse events to the coordinating centers and entered baseline characteristics, process variables and outcome data from patient files into web-based case report forms for each of the days 1 to 14 and a single day 28 follow-up form. Where available, regional and national registries were used for follow-up, and patients or surrogates were contacted directly if needed. Trial data were monitored at the sites, including consent and source data verification, by independent monitors according to a prespecified monitoring plan and centrally by staff from the coordinating centers.

### Outcomes

The primary outcome was the number of days alive without life support (i.e. invasive mechanical ventilation, circulatory support or renal replacement therapy) at 28 days after randomization (all outcome definitions are presented in eMethods in Supplement 2). Secondary outcomes were death from any cause at 28 days after randomization and the number of patients with one or more serious adverse reactions at 28 days (i.e. new episodes of septic shock, invasive fungal infection, clinically important gastrointestinal bleeding, or anaphylactic reaction to dexamethasone). Six additional secondary outcomes will be assessed at day 90 or day 180 after randomization and reported later.^14^

### Statistical analysis

We estimated that 1000 patients were required for the trial to have 85% power to show a relative reduction of 15% in 28-day mortality combined with a 10% reduction in time on life support at a 2-sided alpha-level of 5%, assuming that 30% would die and 10% still be on life support at day 28 in the control group.^1^ The DSMC statistician conducted the interim analysis after the first 500 patients had been followed for 28 days. The alpha values for the interim analysis and the final analysis were 0.0054 and 0.0492, respectively, as per O’Brien-Fleming bounds.^17^ We therefore considered P < 0.0492 statistically significant and correspondingly used 95.08% (rounded to 95%) confidence intervals (CI) for the primary outcome analyses. For the secondary outcomes, we considered P < 0.01 statistically significant and correspondingly used adjusted 99% CIs because of multiple comparisons.

The statistical analyses were performed according to the statistical analysis plan with some modifications (the changes are presented in Supplement 1).^14^ We conducted the primary analyses in the intention-to-treat population, defined as all randomized patients except 18 patients whom did not consent to use of their data. Another 8 patients were erroneously randomized (eTable 1a in Supplement 2); they were included and analyzed. In the per-protocol population, we excluded patients having one or more major protocol violations (eTable 2 in Supplement 2).

In the primary analysis, we analyzed the number of days alive without life support within the 28-day period using the Kryger Jensen and Lange test^18^ adjusted for stratification variables with results presented as adjusted means and medians with 95% CIs. We did the following secondary analysis of the primary outcome, (i) one adjusted for the stratification variables and additional predefined risk factors present at baseline (history of ischemic heart disease or heart failure, diabetes mellitus, chronic pulmonary disease, use of immunosuppressive therapy with the last 3-months, use of circulatory support and use of renal replacement therapy), (ii) one in the per-protocol population, and (iii) analyses in prespecified subgroups, which were defined at baseline by enrolment region (Europe or India), age (below 70 years or above), chronic use of systemic corticosteroids or not, presence or absence of limitations in care, use of invasive mechanical ventilation or not, use of IL-6-RA or not and use of dexamethasone for up to 2 days or 3 to 4 days prior to randomization.

For the secondary outcomes, we used logistic regression adjusted for the stratification variables and g-computation or generalized linear models with log links and binomial error distributions. We also performed unadjusted Fisher’s exact tests.

We made logical imputations for partly missing primary outcome data in 2 patients (eMethods in Supplement 2). For those 11 patients, who had partly missing primary outcome data because consent was withdrawn during the 28-day data collection period, we made best-worst and worst-best imputations of the days they had missing data (eTable 4 in Supplement 2); these 11 patients were included in the analysis of serious adverse reactions without imputation of missing data. We performed analyses using R software, versions 3.6.3 and 4.0.2.

Post hoc we supplemented the primary outcome analysis with one of bootstrapped (50,000 samples) adjusted mean differences because the observed distribution of the primary outcome was markedly skewed (41.4% of all participants had 28 days alive without life support). In another post hoc analysis decided before database lock, we analyzed the primary outcome assigning dead participants the worst possible outcome (i.e. zero days alive without life support) as done in a previous trial (eTable 5 in Supplement 2).^16^

## Results

### Participants

From August 27, 2020 to May 20, 2021, we screened 1414 patients, randomized 1000 and included 982 in the intention-to-treat population (485 patients in Denmark, 369 in India, 79 in Sweden and 49 in Switzerland; Figure 1 and eFigure 2 and 3 in Supplement 2); 497 were assigned to receive dexamethasone 12 mg and 485 to receive dexamethasone 6 mg. We obtained data for the primary outcome for 971 of the 982 patients in the intention-to-treat population (Figure 1). Patient characteristics at baseline were largely similar in the 2 groups (Table 1).

**Figure 2.**
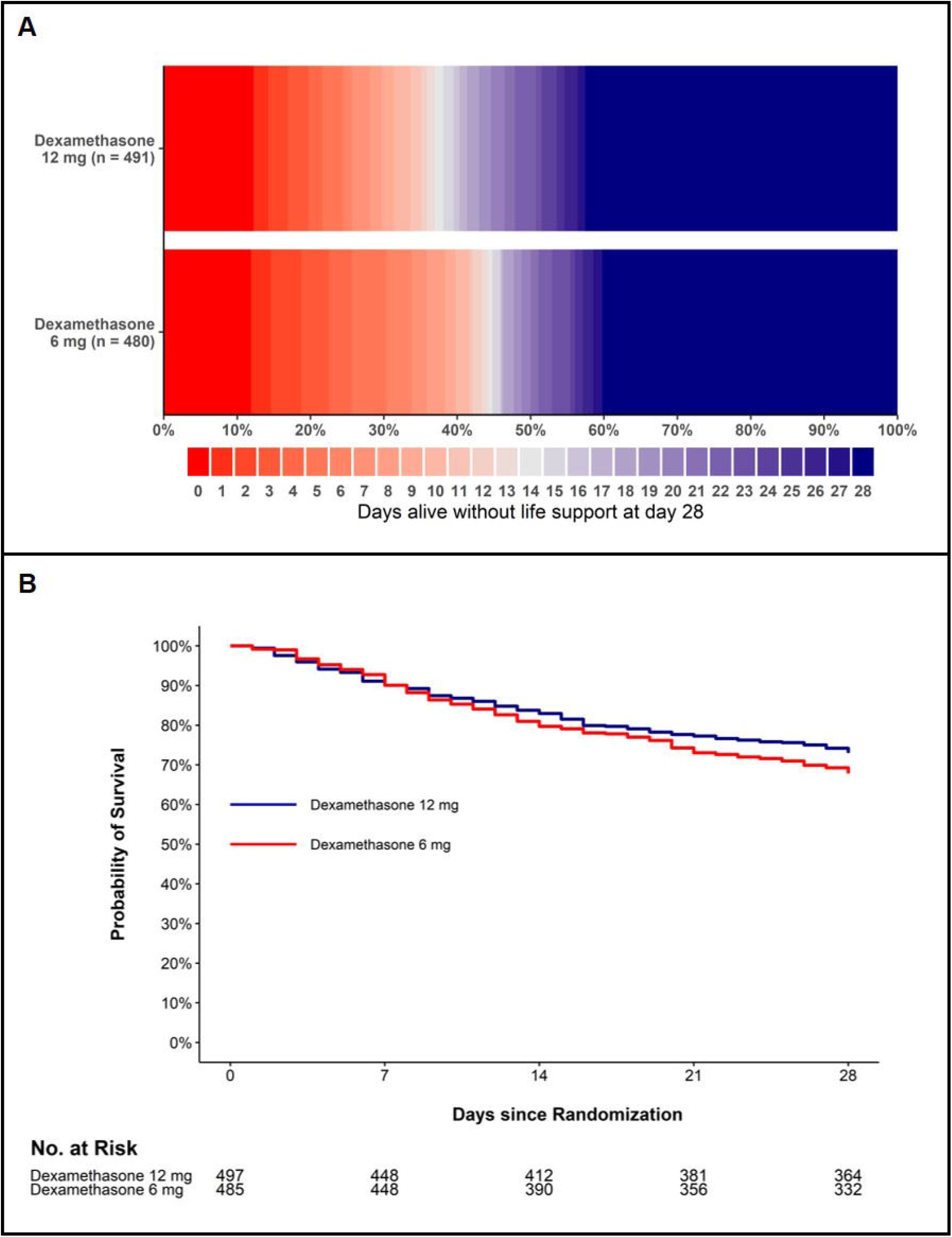
Distributions of the Primary Outcome and Survival Curves up to Day 28. Panel A shows the number of days alive without life support as horizontally stacked proportions in the 2 intervention groups in the intention-to-treat population (n=971; 11 patients had missing primary outcome data). Red represents worse outcomes, and blue represents better outcomes. Panel B shows the survival curves censored at day 28 for the 2 intervention groups in the intention-to-treat population (n=982). The 11 patients who were not followed for the full 28 days (6 patients in the dexamethasone 12 mg group and 5 patients in the dexamethasone 6 mg group) were included in the survival curves until the last day they were known to be alive; at this time point they were censored.

**Figure 3.**
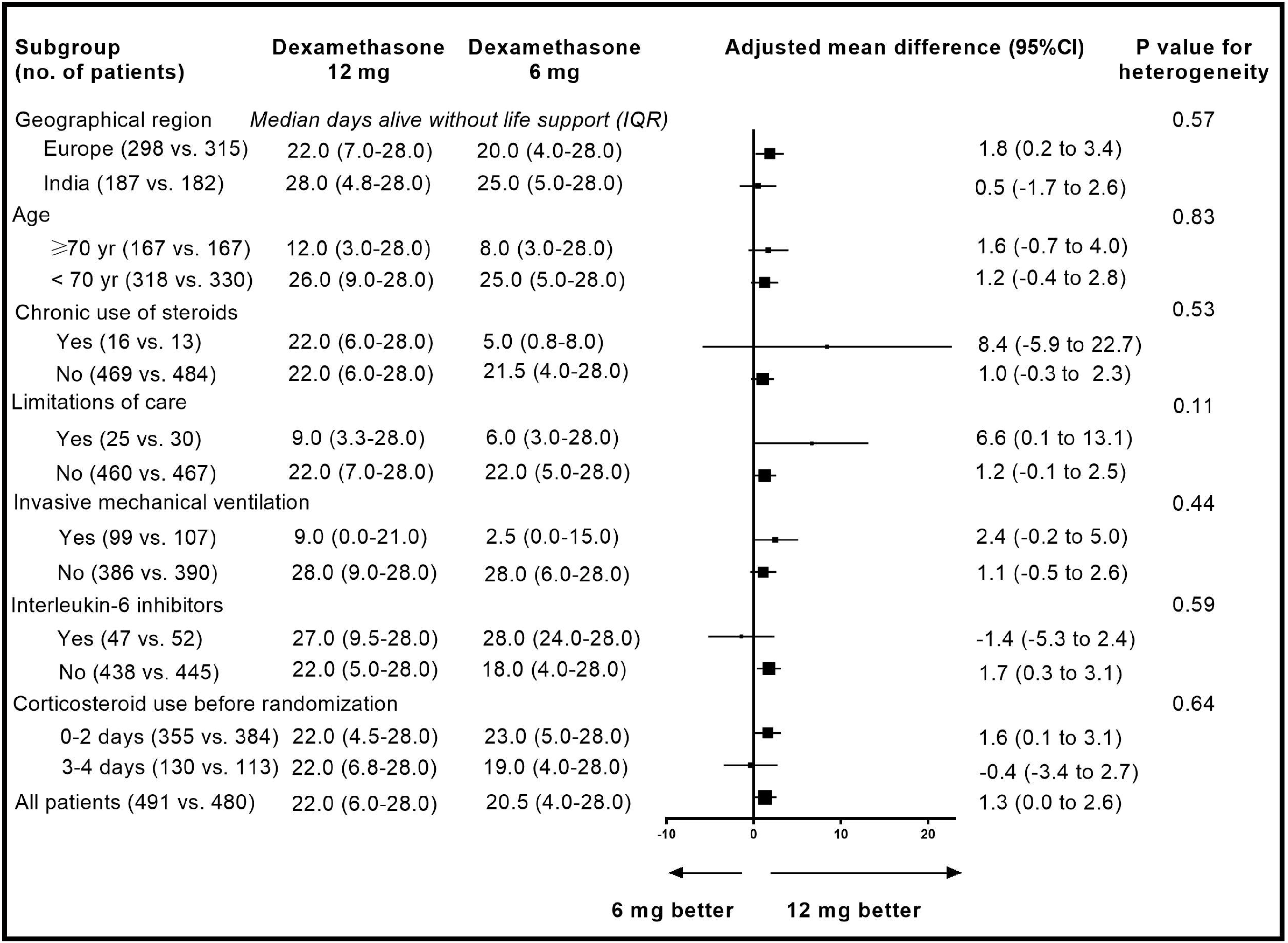
Adjusted Mean Differences in the Primary Outcome in the Predefined Subgroups. Shown are the adjusted mean differences with 95% confidence intervals (CI) for the primary outcome, the number of days alive without life support at day 28, in the dexamethasone 12 mg group compared with the dexamethasone 6 mg group in all patients and in the 7 pre-defined subgroups.

**Table 1.**
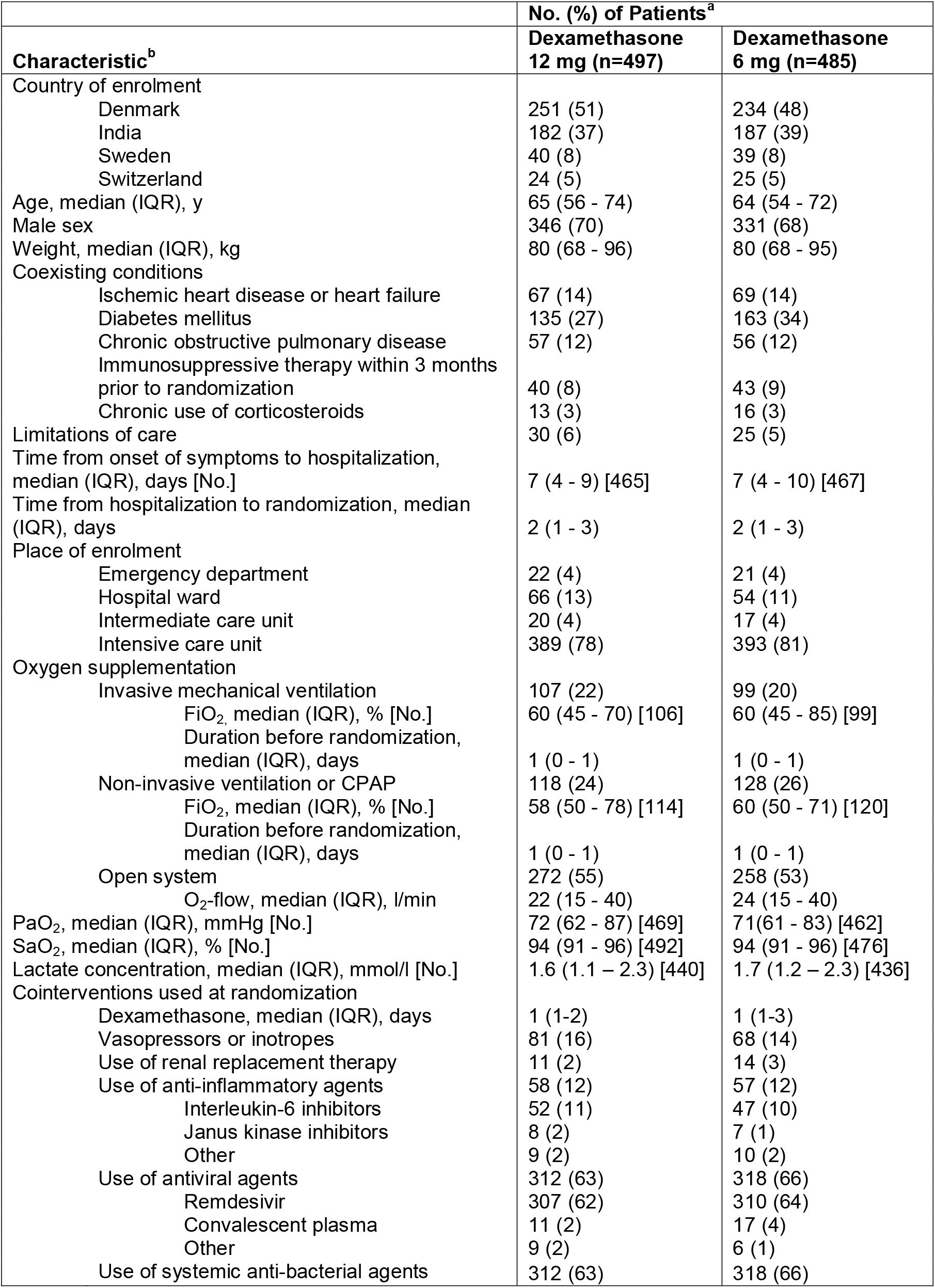
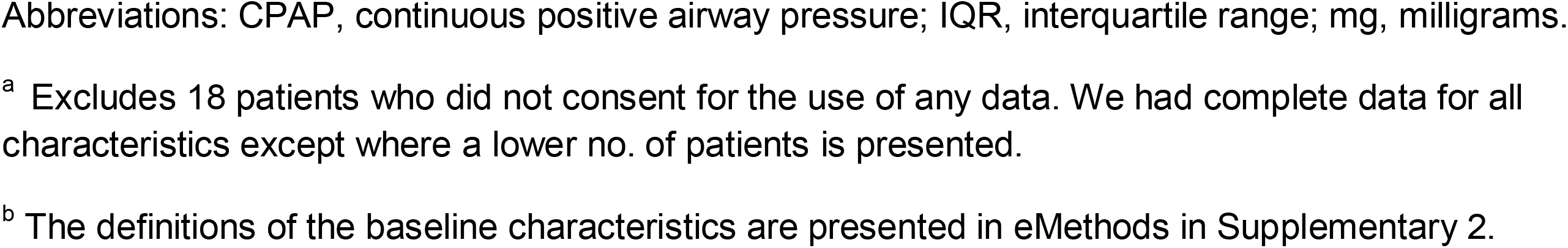
Patient Baseline Characteristics.

| Characteristic<sup>b</sup> | No. (%) of Patients<sup>a</sup> |  |
| --- | --- | --- |
|  | Dexamethasone<br>12 mg (n=497) | Dexamethasone<br>6 mg (n=485) |
| Country of enrolment |  |  |
| Denmark | 251 (51) | 234 (48) |
| India | 182 (37) | 187 (39) |
| Sweden | 40 (8) | 39 (8) |
| Switzerland | 24 (5) | 25 (5) |
| Age, median (IQR), y | 65 (56 - 74) | 64 (54 - 72) |
| Male sex | 346 (70) | 331 (68) |
| Weight, median (IQR), kg | 80 (68 - 96) | 80 (68 - 95) |
| Coexisting conditions |  |  |
| Ischemic heart disease or heart failure | 67 (14) | 69 (14) |
| Diabetes mellitus | 135 (27) | 163 (34) |
| Chronic obstructive pulmonary disease | 57 (12) | 56 (12) |
| Immunosuppressive therapy within 3 months<br>prior to randomization | 40 (8) | 43 (9) |
| Chronic use of corticosteroids | 13 (3) | 16 (3) |
| Limitations of care | 30 (6) | 25 (5) |
| Time from onset of symptoms to hospitalization,<br>median (IQR), days [No.] | 7 (4 - 9) [465] | 7 (4 - 10) [467] |
| Time from hospitalization to randomization, median<br>(IQR), days | 2 (1 - 3) | 2 (1 - 3) |
| Place of enrolment |  |  |
| Emergency department | 22 (4) | 21 (4) |
| Hospital ward | 66 (13) | 54 (11) |
| Intermediate care unit | 20 (4) | 17 (4) |
| Intensive care unit | 389 (78) | 393 (81) |
| Oxygen supplementation |  |  |
| Invasive mechanical ventilation | 107 (22) | 99 (20) |
| FiO<sub>2</sub>, median (IQR), % [No.] | 60 (45 - 70) [106] | 60 (45 - 85) [99] |
| Duration before randomization,<br>median (IQR), days | 1 (0 - 1) | 1 (0 - 1) |
| Non-invasive ventilation or CPAP | 118 (24) | 128 (26) |
| FiO<sub>2</sub>, median (IQR), % [No.] | 58 (50 - 78) [114] | 60 (50 - 71) [120] |
| Duration before randomization,<br>median (IQR), days | 1 (0 - 1) | 1 (0 - 1) |
| Open system | 272 (55) | 258 (53) |
| O<sub>2</sub>-flow, median (IQR), l/min | 22 (15 - 40) | 24 (15 - 40) |
| PaO<sub>2</sub>, median (IQR), mmHg [No.] | 72 (62 - 87) [469] | 71 (61 - 83) [462] |
| SaO<sub>2</sub>, median (IQR), % [No.] | 94 (91 - 96) [492] | 94 (91 - 96) [476] |
| Lactate concentration, median (IQR), mmol/l [No.] | 1.6 (1.1 - 2.3) [440] | 1.7 (1.2 - 2.3) [436] |
| Cointerventions used at randomization |  |  |
| Dexamethasone, median (IQR), days | 1 (1-2) | 1 (1-3) |
| Vasopressors or inotropes | 81 (16) | 68 (14) |
| Use of renal replacement therapy | 11 (2) | 14 (3) |
| Use of anti-inflammatory agents | 58 (12) | 57 (12) |
| Interleukin-6 inhibitors | 52 (11) | 47 (10) |
| Janus kinase inhibitors | 8 (2) | 7 (1) |
| Other | 9 (2) | 10 (2) |
| Use of antiviral agents | 312 (63) | 318 (66) |
| Remdesivir | 307 (62) | 310 (64) |
| Convalescent plasma | 11 (2) | 17 (4) |
| Other | 9 (2) | 6 (1) |
| Use of systemic anti-bacterial agents | 312 (63) | 318 (66) |

Abbreviations: CPAP, continuous positive airway pressure; IQR, interquartile range; mg, milligrams.
<sup>a</sup> Excludes 18 patients who did not consent for the use of any data. We had complete data for all characteristics except where a lower no. of patients is presented.
<sup>b</sup> The definitions of the baseline characteristics are presented in eMethods in Supplementary 2.

### Trial and concomitant interventions

Both groups had received dexamethasone for a median of 1 day before enrolment. The use of respiratory, circulatory and renal support and other anti-inflammatory, antiviral, and anti-bacterial agents were similar between groups at baseline (Table 1).

The assigned trial intervention was received as per protocol by 461 of 497 patients (92.7%) in the 12 mg group and by 446 of 485 (91.9%) in the 6 mg group (eTable 2 in Supplement 2). The duration of the trial intervention was similar in the 2 groups (median 7 days, IQR 5.0 to 9.0 and 7 days, 6.0 to 9.0, respectively, eTable 2 in Supplement 2). In the intervention period, 10 of 497 patients (2.0%) in the 12 mg group and 9 of 485 (1.9%) in the 6 mg group received open-label corticosteroids (eTable 2 in Supplement 2). Nine (1.8%) patients in the dexamethasone 12 mg group and 11 (2.3%) in the dexamethasone 6 mg group were discharged from hospital within 28 days against medical advice (eTable 10 in Supplement 2).

### Primary outcome

At 28 days after randomization, the median number of days alive without life support was 22.0 days (IQR 6.0 to 28.0) in the dexamethasone 12 mg group and 20.5 days (4.0 to 28.0) in the dexamethasone 6 mg group (adjusted mean difference, 1.3; 95% CI 0.0 to 2.6; P=0.066) (Figure 2 and Table 2). The results were similar in the pre-planned (Table 2 and eTables 3 and 4 in Supplement 2) and the post hoc sensitivity analyses (Table 2 and eTable 5 in Supplement 2). In predefined subgroup analyses, we found no statistically significant heterogeneity in the effect of the trial intervention on the primary outcome (Figure 3). The single components of the composite primary outcome were similar between allocation groups (Table 2 and eTable 6 in Supplement 2).

**Table 2.**
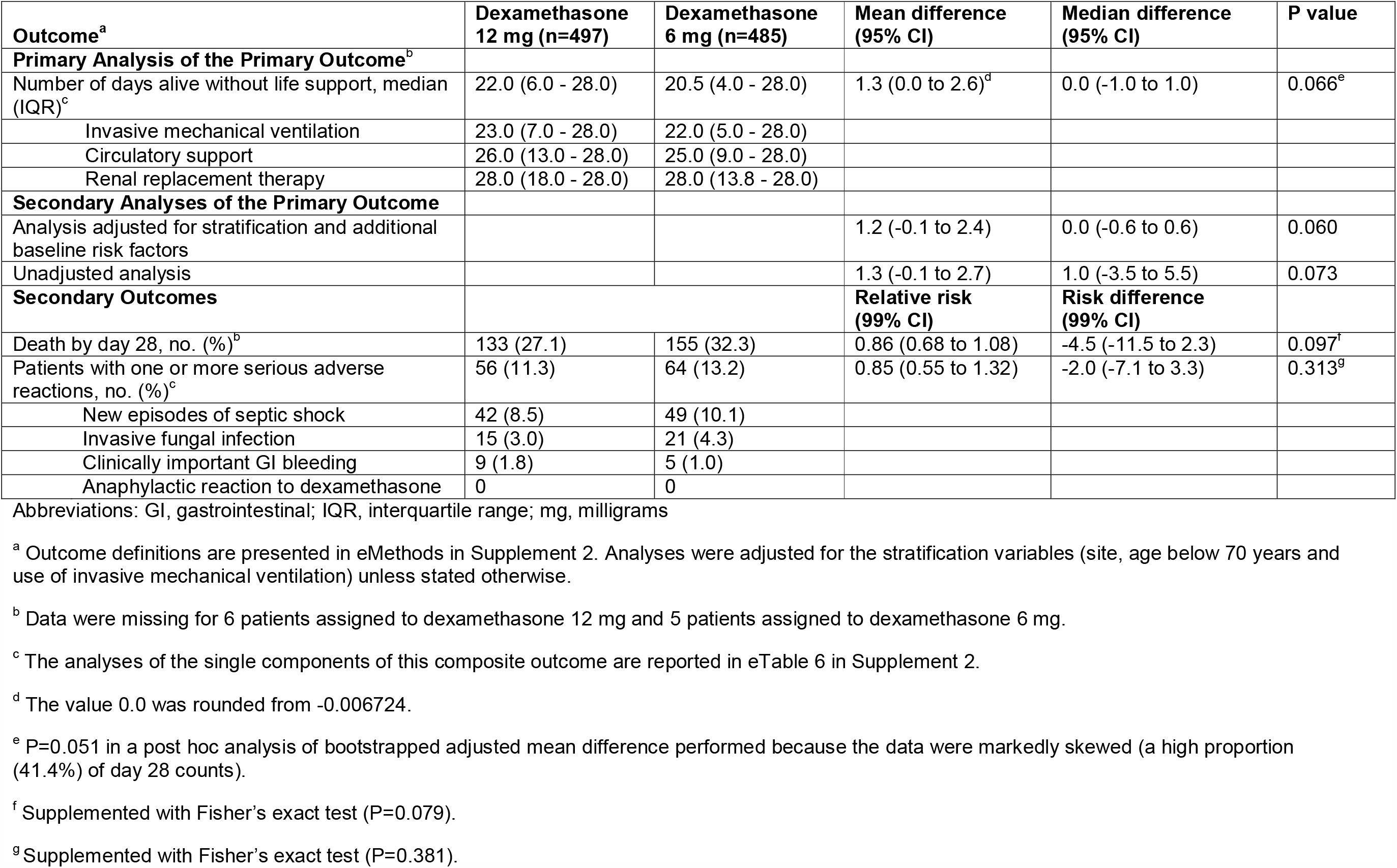
Outcomes at Day 28 in the Intention-To-Treat Population.

| **Outcome<sup>a</sup>** | **Dexamethasone**<br>**12 mg (n=497)** | **Dexamethasone**<br>**6 mg (n=485)** | **Mean difference**<br>**(95% CI)** | **Median difference**<br>**(95% CI)** | **P value** |
| --- | --- | --- | --- | --- | --- |
| **Primary Analysis of the Primary Outcome<sup>b</sup>** |  |  |  |  |  |
| Number of days alive without life support, median (IQR)<sup>c</sup> | 22.0 (6.0 - 28.0) | 20.5 (4.0 - 28.0) | 1.3 (0.0 to 2.6)<sup>d</sup> | 0.0 (-1.0 to 1.0) | 0.066<sup>e</sup> |
| Invasive mechanical ventilation | 23.0 (7.0 - 28.0) | 22.0 (5.0 - 28.0) |  |  |  |
| Circulatory support | 26.0 (13.0 - 28.0) | 25.0 (9.0 - 28.0) |  |  |  |
| Renal replacement therapy | 28.0 (18.0 - 28.0) | 28.0 (13.8 - 28.0) |  |  |  |
| **Secondary Analyses of the Primary Outcome** |  |  |  |  |  |
| Analysis adjusted for stratification and additional baseline risk factors |  |  | 1.2 (-0.1 to 2.4) | 0.0 (-0.6 to 0.6) | 0.060 |
| Unadjusted analysis |  |  | 1.3 (-0.1 to 2.7) | 1.0 (-3.5 to 5.5) | 0.073 |
| **Secondary Outcomes** |  |  | **Relative risk**<br>**(99% CI)** | **Risk difference**<br>**(99% CI)** |  |
| Death by day 28, no. (%)<sup>b</sup> | 133 (27.1) | 155 (32.3) | 0.86 (0.68 to 1.08) | -4.5 (-11.5 to 2.3) | 0.097<sup>f</sup> |
| Patients with one or more serious adverse reactions, no. (%)<sup>c</sup> | 56 (11.3) | 64 (13.2) | 0.85 (0.55 to 1.32) | -2.0 (-7.1 to 3.3) | 0.313<sup>g</sup> |
| New episodes of septic shock | 42 (8.5) | 49 (10.1) |  |  |  |
| Invasive fungal infection | 15 (3.0) | 21 (4.3) |  |  |  |
| Clinically important GI bleeding | 9 (1.8) | 5 (1.0) |  |  |  |
| Anaphylactic reaction to dexamethasone | 0 | 0 |  |  |  |

Abbreviations: GI, gastrointestinal; IQR, interquartile range; mg, milligrams
<sup>a</sup> Outcome definitions are presented in eMethods in Supplement 2. Analyses were adjusted for the stratification variables (site, age below 70 years and use of invasive mechanical ventilation) unless stated otherwise.
<sup>b</sup> Data were missing for 6 patients assigned to dexamethasone 12 mg and 5 patients assigned to dexamethasone 6 mg.
<sup>c</sup> The analyses of the single components of this composite outcome are reported in eTable 6 in Supplement 2.
<sup>d</sup> The value 0.0 was rounded from -0.006724.
<sup>e</sup> P=0.051 in a post hoc analysis of bootstrapped adjusted mean difference performed because the data were markedly skewed (a high proportion (41.4%) of day 28 counts).
<sup>f</sup> Supplemented with Fisher's exact test (P=0.079).
<sup>g</sup> Supplemented with Fisher's exact test (P=0.381).

### 28-day mortality

At 28 days, a total of 133 of 491 patients (27.1%) in the dexamethasone 12 mg group and 155 of 480 (32.3%) in the dexamethasone 6 mg group had died (adjusted relative risk, 0.86; 99% CI 0.68 to 1.08) (Figure 2, Tables 2 and eTable 7 in Supplement 2).

### Serious adverse reactions and events

At 28 days, 56 of 497 patients in the dexamethasone 12 mg group (11.3%), as compared with 64 of 485 patients (13.2%) in the dexamethasone 6 mg group, had one or more serious adverse reactions (adjusted relative risk, 0.85; 99% CI 0.55 to 1.32) (Table 2); the components of the composite outcome are presented in Table 2 and eTables 6 and 7 in Supplement 2; none had anaphylactic reactions to dexamethasone.

The total numbers of patients with one or more serious adverse reactions (registered as the above secondary outcome or reported by investigators) or serious adverse events (reported by investigators) were 108 (21.7%) and 130 (26.8%) in the 12 mg and 6 mg groups, respectively (eTable 8 in Supplement 2). Extracorporeal membrane oxygenation was used in 3 (0.6%) and 14 (2.9%) patients in the 12 mg and 6 mg groups, respectively (eTable 9 in Supplement 2).

## Discussion

In this international, blinded, randomized trial in adults with COVID-19 and severe hypoxia, treatment with dexamethasone 12 mg did not result in statistically significantly more days alive without life support at 28 days as compare with dexamethasone 6 mg, but the lower limit of the 95% CI around the point estimate was 0.0 suggesting that the results are most compatible with benefit from 12 mg. We found no heterogeneity of the intervention effect in any of the prespecified subgroups. The 28-day mortality result was more uncertain as the CI around the point estimate was wide and compatible both with marked benefit and with harm. The number of patients with serious adverse reactions (i.e. septic shock, invasive fungal infection and clinically important gastrointestinal bleeding) appeared similar between the groups.

Our results suggest that the 12 mg dose of dexamethasone may offer additional anti-inflammatory effects without an increase in serious adverse reactions and events, including septic shock and invasive fungal infections. However, our trial was not powered to assess differences in infectious complications between groups.

Our results are in line with those of other trials assessing treatments providing anti-inflammatory effect in addition to that of dexamethasone 6 mg in patients with COVID-19; most patients also received dexamethasone as part of usual care in the IL-6-RA domains of the REMAP-CAP and RECOVERY trials.^16,19^ Both these trials showed improvements in organ support-free days and/or short-term mortality; the absolute mortality benefit with tocilizumab was 8 percent points in REMAP-CAP and 4 percent points in RECOVERY.^16,19^ In our trial, adjusted 28-day mortality was 4.5 percent points lower in the dexamethasone 12 mg group than in the 6 mg group, but this was not statistically significantly different. These potential differences may be due to differences in anti-inflammatory modulation, patient populations, outcome definitions, statistical frameworks (REMAP-CAP used Bayesian statistics) or sample sizes and event rates. Additional analyses of our trial (outcomes at day 90 and 180 and a Bayesian analysis of day 28 and 90 outcomes),^14,20^ and a planned prospective meta-analysis of trials assessing higher versus standard dose dexamethasone in patients with COVID-19 and hypoxia^21^ may allow more firm inferences.

The strengths of our trial include the pragmatic protocol, the relatively large sample size, the inclusion of most of the eligible patients, the allocation concealment and blinding, the complete follow-up, the variety of hospitals and countries involved and that patients were enrolled in both Europe (62%) and India (38%) reflecting different patient characteristics and risk factors, practice patterns and health care systems. Septic shock and invasive fungal infections were specified secondary safety outcomes and therefore accurately captured. We challenged the results in multiple sensitivity analyses, in those of the per protocol population and in the prespecified subgroups and found similar results as in the primary analysis. Together, these characteristics increase the internal and external validity of our results.

### Limitations

We had limited power to detect differences in some of the outcomes and in the subgroup analyses. Some baseline variables may have differed between the groups, e.g. diabetes, but a pre-defined secondary analysis adjusting for diabetes and other important risk factors supported the primary result. As expected, the distribution of the primary outcome data was not normal, but it was unexpected that day 28 would have 41.4% of the counts. To mitigate this, we used a newly developed statistical test that accounts for excess zeroes and post hoc used bootstrapping to challenge the results further.^18^ The sample size estimation for the primary outcome was based on expected relative differences in 28-day mortality and time on life support of 15% and 10%, respectively, that may be considered large. Changes in the treatment of COVID-19 during trial (e.g. increased use of IL-6-RAs) may have influenced the results.

## Conclusions

Among patients with COVID-19 and severe hypoxia, dexamethasone 12 mg did not result in statistically significantly more days alive without life support at 28 days than dexamethasone 6 mg. However, the confidence interval around the point estimate should be considered when interpreting the results of this trial.

## Data Availability

Fully de-identified data will be available 1-year after the publication of the trial data upon application to the Management Committee

http://www.cric.nu/covid-steroid-2-trial-documents/

## Role of the Funder/Supporter

Neither the funder nor the supporter had roles in the design of the protocol, trial conduct, or the analyses or reporting of the data.

## Group information

### Management Committee

Anders Perner (Sponsor and Chair of Management Committee), Marie Warrer Munch (International Coordinating Investigator), Maj-Brit Nørregaard Kjær (International Project Manager), Anders Granholm (National investigator - Denmark), Morten Hylander Møller (National investigator - Denmark), Tine Sylvest Meyhoff (National investigator - Denmark), Gitte Kingo Vesterlund (National Project Manager - Denmark), Marie Helleberg (National investigator - Denmark), Thomas Benfield (National investigator - Denmark), Balasubramanian Venkatesh (Co-chair), Naomi Hammond (Senior Research Fellow and Operations Lead - Australia), Sharon Micallef (Project Manager - Australia), Vivekanand Jha (Co-chair), Bharath Kumar Tirupakuzhi Vijayaraghavan (National Investigator - India), Sheila Nainan Myatra (National Coordinating Investigator - India), Maria Cronhjort (National Investigator - Sweden), Rebecka Rubenson Wahlin (National Coordinating Investigator - Sweden), Stephan Jakob (National Investigator - Switzerland), Luca Cioccari (National Coordinating Investigator - Switzerland), Christian Gluud (Trialist), and Theis Lange (Statistician).

### Management Committee for Indian Sites

Balasubramanian Venkatesh (Co-chair), Vivekanand Jha (Co-chair), Sheila Nainan Myatra (National Coordinating Investigator), Naomi Hammond (Senior Research Fellow and Operations Lead – Australia), Abhinav Bassi (Project Manager - India), Sharon Micallef (Project Manager - Australia), Anubhuti Jha (Site Coordinator – India), Mallikarjuna Kunigari (Program Manager – India), Bharath Kumar Tirupakuzhi Vijayaraghavan (National Investigator), Oommen John (Senior Research Fellow – India), Ashwani Kumar (Research Associate – Australia and India), Anders Perner (Sponsor), Marie Warrer Munch (International Coordinating Investigator), Maj-Brit Nørregaard Kjær (International Project Manager).

### Regional Coordinating Centre for India

The George Institute for Global Health, Sydney, Australia: Naomi Hammond, Ashwani Kumar, Sharon Micallef, Dorrilyn Rajbhandari, Balasubramanian Venkatesh

### Local Coordinating Centre for India

The George Institute for Global Health, New Delhi, India: Sumaiya Arfin, Abhinav Bassi, Nikita Bathla, Anubhuti Jha, Vivekanand Jha, Oommen John, Rajesh Joshi, Mallikarjuna Kunigari.

### Independent Data Monitoring and Safety Committee

Christian Hassager (Clinician), Manu Shankar-Hari (Trialist), and Susanne Rosthøj (Statistician).

## The COVID STEROID 2 trial investigators

see Supplement 2

## Additional Contributions

We would like to thank all patients and relatives for agreeing to participate in the COVID STEROID 2 trial; all clinical and research staff at the participating hospitals; the regulatory authorities in the participating countries for the expedite handling of the protocol; and the funding sources. We would also like to thank all clinical and research staff involved in the design and conduct of the ‘Low-dose hydrocortisone in patients with Covid-19 and severe hypoxia (COVID STEROID) trial’ which preceded the COVID STEROID 2 trial.

