## Supplementary material for "Dexamethasone 12 mg versus 6 mg for patients with COVID-19 and severe hypoxia: an international, randomized, blinded trial": Suppl. 1

This supplement contains the following items:

1. Original protocol, final protocol, summary of changes to protocol
2. Original statistical analysis plan, summary of changes to statistical analysis plan

### Higher vs. Lower Doses of Dexamethasone in Patients with COVID-19 and Severe Hypoxia: the COVID STEROID 2 trial

**Management Committee**

Anders Perner, sponsor

Morten Hylander Møller

Marie Warrer Petersen

Maj-Brit Nørregaard Kjær

Tine Sylvest Meyhoff

Marie Helleberg

Anders Granholm

Gitte Kingo Vesterlund

Thomas Benfield

Steffen Christensen

Maria Cronhjort

Rebecka Rubenson Wahlin

Stephan Jakob

Luca Cioccari

Balasubramanian Venkatesh

Vivekanand Jha

Bharath Kumar Tirupakuzhi Vijayaraghavan

Sheila Nainan Myatra

Naomi Hammond

Oommen John

Abhinav Bassi

Sharon Micallef

Christian Gluud, trialist

Theis Lange, statistician

**Protocol version and date**

1.7, 17.08.2020

**Applicable protocol registration numbers**

ClinicalTrials.gov identifier NCT04509973

Ethics committee number H-20051056

EudraCT number 2020-003363-25

Danish Medicines Agency number 2020-07-16

### 1 Abstract

**Background**

Severe acute respiratory syndrome coronavirus-2 (SARS-CoV-2) is causing a pandemic of coronavirus disease 2019 (COVID-19) with many patients developing severe hypoxic respiratory failure. Many patients have died, and healthcare systems in several countries have been or will be overwhelmed because of a surge of patients needing hospitalisation and intensive care. The care in COVID-19 is primarily supportive, including respiratory and circulatory support.

Preliminary results from the Randomised Evaluation of COVid-19 thERapY (RECOVERY) trial have reported a reduction in 28-day mortality with low-dose dexamethasone (6 mg) once daily versus no intervention in hospitalised patients with COVID-19; an effect that may have been more pronounced in patients with increasing hypoxia. Yet, higher doses of dexamethasone may be beneficial in patients with non-COVID-19 acute respiratory distress syndrome. At present, it is unclear what dose of dexamethasone is most beneficial in patients with COVID-19 and severe hypoxia, and clinical equipoise exists.

**Objectives**

We aim to assess the effects of higher (12 mg) vs lower doses (6 mg) of intravenous dexamethasone on the number of days alive without life-support in adult patients with COVID-19 and severe hypoxia.

**Design**

International, parallel-group, centrally randomised, stratified, blinded, clinical trial.

**Inclusion and exclusion criteria**

We will screen all adult patients who have documented COVID-19 receiving at least 10 L/min of oxygen independent of delivery system OR mechanical ventilation. We will exclude patients who have an indication for systemic use of higher doses of corticosteroids (above 6 mg dexamethasone or equivalent) for other indications than COVID-19, who have received corticosteroids for COVID-

19 for 5 consecutive days or more, who have invasive fungal infection, who have active tuberculosis, who have known hypersensitivity to dexamethasone, who are pregnant, and those in whom informed consent cannot be obtained.

**Experimental intervention**

Dexamethasone 12 mg once daily for up to 10 days will be given as bolus injection.

**Control intervention**

Dexamethasone 6 mg once daily for up to 10 days will be given as bolus injection.

**Outcomes**

The primary outcome is days alive without life support (invasive mechanical ventilation, circulatory support, or renal replacement therapy) at day 28. Secondary outcomes are serious adverse reactions (new episode of septic shock, invasive fungal infection, clinically important gastrointestinal bleeding, or anaphylactic reaction to dexamethasone) at day 28; days alive without life support at day 90; days alive and out of hospital at day 90; all-cause mortality at day 28, day 90 and 180 days; and health-related quality of life at 180 days.

**Statistics**

The primary outcome will be compared using the Kryger Jensen and Lange test and reported as differences in means and medians along with 95% confidence intervals adjusted for the stratification variables (site, use of invasive mechanical ventilation, and age). Secondary binary outcomes will be analysed using binomial regression models with log links with results quantified as adjusted relative risks supplemented with adjusted risk differences, both with 95% confidence intervals.

**Trial size and testing strategy/design**

At maximum, we will randomise 1000 participants. The independent data monitoring and safety committee will conduct an interim analysis after 500 participants have been followed for 28 days.

The alpha values for the interim analysis and the final analysis are 0.0054 and 0.0492, respectively as by the O'Brien-Fleming bounds, which preserves type I error at the usual 5%. In both analyses, the Kryger Jensen and Lange test will be employed to compare the groups on the primary outcome.

**Estimated timeline**

- August 2020, authority approvals and 1$^{st}$ patient randomised

- December 2020, interim analysis

- Mid 2021, last patient randomised and primary report on 28-day outcomes submitted.

- Late 2021, report on 90-day outcomes submitted

- Mid 2022, report on 180-day outcomes submitted

### 2 Administrative information

#### *2.1 Local and International Sponsors*

**International Sponsor/Central Coordinating Centre**

Dept. of Intensive Care

Rigshospitalet

Blegdamsvej 9
2100 Copenhagen Ø

+453545 7167

**International Sponsor Contact**

Anders Perner, senior staff specialist and professor in intensive care medicine

Dept. of Intensive Care

Rigshospitalet

Blegdamsvej 9

2100 Copenhagen Ø

+45 3545 8333

**Local sponsor /Coordinating Centre, Sweden**

Dept. of Clinical Science and Education

Södersjukhuset, Karolinska Institutet
Sjukhusbacken 10

11883 Stockholm

**Local Sponsor contact, Sweden**

Maria Cronhjort, senior staff specialist in intensive care medicine

Dept. of Anaesthesia and Intensive Care

Södersjukhuset, Karolinska Institutet
Sjukhusbacken 10

11883 Stockholm

**Local sponsor /Coordinating Centre, Switzerland**

Department of Intensive Care Medicine

Bern University Hospital (Inselspital)

Freiburgstrasse 15

3010 Bern

+41 31 632 53 00

**Local Sponsor contact, Switzerland**

Stephan Jakob, senior staff specialist and professor in intensive care medicine

Department of Intensive Care Medicine

Bern University Hospital (Inselspital)

Freiburgstrasse 15

3010 Bern

**Local sponsor /Coordinating Centre, India**

The George Institute for Global Health, New Delhi, India

Plot No. 57, Second Floor, Corporation Bank Building

Nagarjuna Circle, Punjagutta, Hyderabad - 500 082 | Telangana | India

T +91 40 3099 4444

F +91 40 3099 4400

**Local Sponsor contact, India**

Vivekanand Jha

The George Institute for Global Health, New Delhi, India

#### *2.2 Local investigators and clinical trial sites*

**Denmark**

Marie Warrer Petersen, Dept. of Intensive Care

Marie Helleberg, Dept. of Infectious Diseases

Vibeke Jørgensen, Dept. of Thoracic Anaesthesiology

Margit Smitt, Dept. of Neuroanaesthesiology

Rigshospitalet

Blegdamsvej 9

2100 Copenhagen Ø

Klaus Tjelle, Dept. of Anaesthesia and Intensive Care

Thomas Benfield, Dept. of Infectious Diseases

Charlotte Suppli Ulrik, Dept. of Respiratory Medicine

Hvidovre Hospital, University of Copenhagen

2650Hvidovre

Anne Sofie Andreasen, Dept. of Anaesthesia and Intensive Care

Herlev Hospital, University of Copenhagen

2730 Herlev

Thomas Mohr, Dept. of Intensive Care

Gentofte Hospital, University of Copenhagen

2900 Hellerup

Morten Bestle, Dept. of Anaesthesia and Intensive Care

North Zealand Hospital, University of Copenhagen
3400 Hillerød

Lone Poulsen, Dept. of Anaesthesia and Intensive Care

Zealand University Hospital, Køge

4600 Køge

Thomas Hildebrandt, Dept. of Anaesthesia and Intensive Care

Zealand University Hospital, Roskilde

4000 Roskilde

Anders Møller, Dept. of Anaesthesia

Slagelse Hospital

4200 Slagelse

Christoffer G. Sølling, Dept. of Anaesthesia and Intensive Care

Viborg Hospital

8800 Viborg

Anne Brøchner, Dept. of Anaesthesia and Intensive Care

Kolding Hospital

6000 Kolding

Iben Strøm Darfelt, Dept. of Anaesthesia

Regional Hospital West Jutland, Herning

7400 Herning

Bodil S. Rasmussen, Dept. of Anaesthesia and Intensive Care

Aalborg University Hospital

9000 Aalborg

Steffen Christensen, Dept. of Anaesthesia and Intensive Care

Aarhus University Hospital

8200 Aarhus

Thomas Strøm, Dept. of Anaesthesia and Intensive Care

Odense University Hospital

5000 Odense

**Sweden**

Maria Cronhjort, Dept. of Anaesthesia and Intensive Care

Rebecka Rubenson Wahlin, Dept. of Anaesthesia and Intensive Care

Carl-Johan Treutiger, Dept. of Infectious diseases

Buster Mannheimer, Dept. Internal Medicine

Jacob Hollenberg, Dept of Cardiology

Södersjukhuset, 11883 Stockholm

118 83 Stockholm

Johan Mårtensson, Dept. of Anaesthesia and Intensive Care

Pontus Naucler, Dept. of Infectious diseases

Karolinska Universitetssjukhuset, Solna

171 76 Stockholm

Åke Norberg, Dept. of Anaesthesia and Intensive Care

Karolinska Universitetssjukhuset, Huddinge

141 86 Stockholm

Olof Wall, Dept. of Anaesthesia and Intensive Care

Sara Tehrani, Dept. Internal Medicine

Magnus Hedenstierna, Dept. of Infectious diseases

Danderyds Sjukhus

182 88 Stockholm

Andreas Wiklund, Dept. of Anaesthesia and Intensive Care

Capio St Görans Sjukhus

112 81 Stockholm

Michelle Chew, Dept. of Anaesthesia and Intensive Care

Universitetssjukhuset i Linköping

581 85 Linköping

Fredrik Schiöler, Dept. of Anaesthesia and Intensive Care

Vrinnevisjukhuset, Norrköping

603 79 Norrköping

Fredrik Sjövall, Dept. of Anaesthesia and Intensive Care

Anna Nilsson, Dept. of Infectious diseases

Skånes universitetssjukhus (SUS) Malmö

214 28 Malmö

Jonathan Oras, Dept. of Anaesthesia and Intensive Care

Magnus Gisslen, Dept. of Infectious diseases

Sahlgrenska universitetssjukhuset

413 45 Göteborg

**Switzerland**

Stephan Jakob, Department of Intensive Care Medicine

Luca Cioccari, Department of Intensive Care Medicine

Bern University Hospital (Inselspital)

3010 Bern

**India**

BK Tirupakuzhi Vijayaraghavan

Apollo Hospitals

600 006 Chennai

JV Divatia

Tata Memorial Hospital

400 012 Mumbai

Farhad N Kapadia

P.D. Hinduja National Hospital & Medical Research Centre

400 016 Mumbai

Pravin Amin

Bombay Hospital & Medical Research Centre

400 020 Mumbai

Sanjith Saseedharan

S L Raheja Fortis Hospital

400 016 Mumbai

Amit Shobhavat

K. J. Somaiya Super Specialty Hospital

400 022 Mumbai

Rajesh Chawla

Indraprastha Apollo Hospital

110 076 New Delhi

Omender Singh

Max Super Specialty Hospital, Saket

110017 New Delhi

Urvi Shukla

Symbiosis University Hospital and Research Centre

412 115 Pune

Binila Chacko

Christian Medical College Vellore
632 004 Vellore

Kedar Toraskar

Wockhardt hospitals

400011 Mumbai

Syed Moied Ahmed

Jawahar Lal Nehru Medical College, AMU

202002 Aligarh

Saif Mohd. Khan

Rajendra Institute of Medical Sciences

834009 Ranchi

Kapil Borawake

Vishwaraj Hospital

412201 Pune

Neeta Bose Nayak

Gotri General Hospital

390021 Vadodara

Zubair Umer Mohamed

Amrita Institute of Medical Sciences

682 041 Kochi

#### *2.3 Methodological trial sites*

**Coordinating centre**

Dept of Intensive Care

Rigshospitalet

Blegdamsvej 9
2100 Copenhagen Ø

+453545 7167

**Methods centre**

Copenhagen Trial Unit, Centre for Clinical Intervention Research

Rigshospitalet
Tagensvej 22

2200 Copenhagen N

**Statistical centre**
Section of Biostatistics
University of Copenhagen
Øster Farimagsgade 5
1014 Copenhagen K

**Independent Data Monitoring and Safety Committee (IDMSC)**

Christian Hassager, Professor in cardiology, Copenhagen University Hospital, Denmark

Manu Shankar-Hari, Clinician Scientist, Reader and Consultant in Intensive Care Medicine, National Institute for Health Research and Kings College, London, United Kingdom

Susanne Rosthøj, Statistician from University of Copenhagen

### 3 List of abbreviations

AE, Adverse Event

AR, Adverse Reaction

ARDS, Acute Respiratory Distress Syndrome

CI, Confidence Interval

CPAP, Continuous Positive Airway Pressure

CRIC, Collaboration for Research in Intensive Care

CTU, Copenhagen Trial Unit

IDMSC, Independent Data Monitoring and Safety Committee

eCRF, Electronic Case Report Form

EudraCT, European Union Drug Regulating Authorities Clinical Trial

GI, Gastrointestinal

GLM, Generalised Linear Model

GRADE, Grading of Recommendations Assessment, Development and Evaluation

HR, Hazard Ratio

HRQoL, Health Related Quality of Life

ICH-GCP, International Conference on Harmonisation on Good-Clinical -Practice

ICMJE, International Committee of Medical Journal Editors

ICU, Intensive Care Unit

IL-6, Interleukin 6

IQR, Interquartile Range

ITT, Intention-to-treat

IV, Intravenous

MD, Mean Difference

OR, Odds Ratio

QoL, Quality of Life

RCT, Randomised clinical trial

RR, Relative Risk

RRI, Relative Risk Increase

RRR, Relative Risk Reduction

SAR, Serious Adverse Reaction

SAE, Serious Adverse Event

SARS, Severe Acute Respiratory Syndrome

SD, Standard Deviation

SR, Systematic review

SSC, Surviving Sepsis Campaign

SUSAR, Suspected Unexpected Serious Adverse Reaction

WHO, World Health Organization

### 4 Introduction and background

#### 4.1 Severe acute respiratory syndrome coronavirus 2/Coronavirus Disease 19

In December 2019, the Wuhan Municipal Health Committee in China identified an outbreak of viral pneumonia cases of unknown cause (1). A novel coronavirus was soon identified as the cause of the disease (1). This novel virus has been named severe acute respiratory syndrome coronavirus 2 (SARS-CoV-2) (2) and the disease caused by the virus has been designated coronavirus disease 2019 (COVID-19) (3). Since the initial outbreak in China in December 2019, SARS-CoV-2 has spread globally and COVID-19 has been declared a pandemic by the World Health Organization (WHO)(4). Currently, the number of reported patients with COVID-19 and associated deaths are, as of July 14, 2020, more than 13.100.000 and 573.000, respectively (5). There are currently large outbreaks in the US, Brazil, Russia, and India with many severely ill patients admitted to hospitals and intensive care units (ICUs).

SARS-CoV-2 causes respiratory tract infection (6). The symptoms vary from mild to severe pneumonia and from mild to severe acute respiratory distress syndrome (ARDS) (6). Current estimates suggest that up to 40% of hospitalised COVID-19 patients develop ARDS (6-10). Further, 20-35% of those patients admitted to the ICU may develop septic shock (6, 8, 9, 11, 12). Both conditions are associated with high morbidity and mortality (6, 13).

#### 4.2 Corticosteroids in COVID-19

The current care in COVID-19 is primarily supportive including oxygen, mechanical ventilation, and general intensive care (14). Many patients are treated with various antiviral drugs or immunomodulatory agents, including corticosteroids (15). Until recently, clinical equipoise existed regarding the use of systemic corticosteroids for COVID-19. The Surviving Sepsis Campaign guidelines on the management of critically ill adults with COVID-19 recommended use of low-dose corticosteroids for shock reversal over no use (weak recommendation, low quality of evidence) and use of corticosteroids over no use for those with ARDS (weak recommendation, low quality of evidence) (11). In contrast, the WHO and the Infectious Diseases Society of America (IDSA) recommended against the use corticosteroids in COVID-19 (16, 17).

A preliminary report from the Randomised Evaluation of COVid-19 thERapY (RECOVERY) trial was released on June 22, 2020 (18). In the RECOVERY trial, 6,425 hospitalised patients with suspected or confirmed COVID-19 were randomised to open-label dexamethasone 6 mg daily for up to 10 days vs. usual care (18). The preprint of the preliminary results reported an overall relative reduction of 17% in 28-day mortality (age-adjusted rate ratio 0.83, 95% confidence interval (CI) 0.74 to 0.92) with indications of greatest benefit among those patients requiring invasive mechanical ventilation (rate ratio 0.65, 95% CI 0.51 to 0.82) (18). These results are supported by similar findings in a recently updated systematic review including patients with non-COVID-19 ARDS (risk ratio (RR) 0.72, 95% CI 0.55 to 0.93) (19).

International collaborative research initiatives have been formed with the aim of harmonising and coordinating data collection to enable prospective meta-analyses of the ongoing randomised trials of corticosteroids for COVID-19. The results of these meta-analyses are still not available. As of June 22, 2020, 16 trials assessing corticosteroids for COVID-19 were registered at ClinicalTrials.gov, many of which have already commenced enrolment (18, 20-34). Of these, the COVID STEROID trial is initiated by the same Sponsor and Management Committee of the COVID STEROID 2 trial (31). The COVID STEROID trial assesses low-dose hydrocortisone 200 mg daily vs. placebo in patients with COVID-19 and severe hypoxia. The trial was commenced on April 15, 2020, but paused after randomising 30 patients on June 16, 2020, due to the press release from the RECOVERY trial (35). The decision to continue or stop the COVID STEROID trial will be made after the peer-reviewed publication of the RECOVERY trial is available as well as results from an ongoing prospective meta-analysis of trials assessing corticosteroids for COVID-19.

#### *4.3 Type and dose of corticosteroids for sepsis, ARDS and COVID-19*

The choice of type, dose, and duration of corticosteroids for treatment of sepsis and ARDS is controversial. Various regimens have been used in different trials (Table 1). Generally, studies with short-course high-dose corticosteroids for sepsis did not show a reduction in mortality or showed increased mortality, whereas studies employing longer-course low-dose steroids showed shock reversal and potentially also a reduction in mortality (36). Clinical guidelines published in 2018 stated that the optimal corticosteroid dose and duration of treatment are still uncertain (37). A later dose-response meta-analysis suggested that long-course (7 days) low-dose (200–300 mg per day) hydrocortisone treatment with cumulative dose ≥1,000 mg was beneficial for the reduction of 28-day mortality in patients with sepsis (38).

Similarly, a meta-analysis of corticosteroids for ARDS was inconclusive regarding short-course high-dose treatment (>30 mg dexamethasone or equivalent per day), whereas early initiation of longer course low-dose corticosteroids (≤30 mg dexamethasone or equivalent per day) reduced the duration of mechanical ventilation and mortality (39).

In the RECOVERY trial, low-dose dexamethasone (6 mg) versus no intervention was shown to reduce 28-day mortality in hospitalised patients with suspected or confirmed COVID-19 (18). The findings from the RECOVERY trial have been implemented in a COVID-19 treatment guideline from the National Institutes of Health (NIH) and the updated guideline from IDSA in which dexamethasone 6 mg is recommended for COVID-19 patients receiving supplemental oxygen or mechanical ventilation (40, 41). Therefore, dexamethasone 6 mg is likely to be part of the standard care of COVID-19 patients receiving supplemental oxygen or mechanical ventilation in most hospitals as observed in our clinical practice survey (results below). The remaining ongoing trials of corticosteroids for COVID-19 assess different corticosteroids (i.e. dexamethasone, methylprednisolone, hydrocortisone, or prednisone) with varying daily doses used (median dexamethasone equivalent dose 15 mg, interquartile range (IQR) 10-16 mg) (20-34). However, the results of these trials have not yet been published (20-34), leaving the optimal dosing for COVID-19 uncertain.

In trials in non-COVID-19 ARDS, the doses used (median dexamethasone equivalent dose 12 mg, IQR 9-16 mg (42-48)) have been within the dosing regimens used in the COVID-19 trials. Of note, higher doses of dexamethasone has previously been used in a clinical trial in non-COVID ARDS suggesting benefit and no obvious harm (42).

In short-term use in healthy volunteers, dose-dependent activation of the corticosteroid receptor has been observed for increasing doses up to 60 mg of prednisone (equivalent to 12 mg of dexamethasone) suggesting that doses up to 12 mg of dexamethasone may offer additional anti-inflammatory effects (49). In that study, the adverse effects were independent of the dosing (49).

We, the COVID STEROID 2 trial investigators, have done a survey of clinical practice in early July 2020 at 26 potential COVID STEROID 2 trial sites after the preprint publication of the RECOVERY trial results. All sites had used corticosteroids for patients with COVID-19; at most sites (95%), all patients had received corticosteroids. Most sites used mainly dexamethasone, and the median steroid dose (in dexamethasone equivalents) used at sites in patients with COVID-19 was 9.6 mg (IQR 6.0 – 15.0 mg).

In a concomitant survey of clinical preferences done early July 2020 among doctors at potential COVID STEROID 2 trial sites after the preprint publication of the RECOVERY trial results, 86% of 250 responding doctors would always or most times use steroids in patients with COVID-19 and hypoxia; 56% would use 6 mg of dexamethasone or equivalent, and 36% would use a dose above 6 mg (unpublished results). As for preferences for an upcoming trial, most doctors (95%) would enrol their patients with severe COVID-19 into a trial of steroids, and most (55%) into one of 12 mg vs. 6 mg dexamethasone (unpublished results).

**Type and dose of corticosteroid in the COVID STEROID 2 trial**

For the COVID STEROID 2 trial, participants in the experimental intervention arm will receive intravenous dexamethasone 12 mg for up to 10 days or until discharge from the participating trial site without tapering. Dexamethasone has previously been used without tapering in a clinical trial assessing an even higher dose of dexamethasone (median 15 mg for 10 days) for non-COVID ARDS with potential benefit and without obvious harm (42).

Participants in the control intervention arm will receive intravenous dexamethasone 6 mg daily for up to 10 days or until discharge from the participating trial site without tapering, which is the exact protocol used in the RECOVERY trial (18). The RECOVERY trial investigators have not yet reported data on adverse events (18).

**Table 1. Estimates on the effects of corticosteroid vs. placebo/no treatment in critically ill patients with severe infection and/or severe respiratory failure:** Most data are from recently updated systematic reviews (SRs) of randomised clinical trials (RCTs), except those from viral acute respiratory distress syndrome (ARDS) and COVID-19.

| | Septic shock (50) | ARDS without COVID-19 (19) | Community acquired pneumonia (51) | Viral ARDS (11) | COVID-19(18) |
|---|---|---|---|---|---|
| Evidence base | SR of 22 RCTs, including 7297 participants | SR of 7 RCTs, including 851 participants | SR of 13 RCTs, including 2005 participants | SR of 10 observational studies on other corona viruses | Predefined subgroup of patients receiving invasive mechanical ventilation in 1 RCT, including 1007 participants in this subgroup |
| Corticosteroid used | Hydrocortisone 18 trials<br>Methylprednisolone 2 trials | Methylprednisolone 3 trials<br>Hydrocortisone 2 trials<br>Inhaled budesonide 2 trials<br>Dexamethasone 1 trial | Hydrocortisone 6 trials<br>Methylprednisolone or prednisolone 5 trials<br>Dexamethasone 1 trials<br>Prednisone 1 trial | Not reported | Dexamethasone |
| Daily dose (dexamethasone-equivalent) | 7.5-11.3 mg/day | 4-32 mg/day | 7.5-15 mg/day | Not reported | 6 mg |
| Mortality | RR 0.98 [0.89 to 1.08] | RR 0.72 [0.55 to 0.93] | RR 0.67 [0.45 to 1.01] | OR 0.83 [0.32 to 2.17] | Rate ratio 0.65 [0.51 to 0.82] |
| Admission to ICU | - | - | RR 0.69 [0.46 to 1.03] | | |
| Need for ventilation | - | - | RR 0.45 [0.26 to 0.79] | | |
| Days ventilated | MD −0.75 [−1.34 to -0.17] days | MD –4.8 [–7.0 to –2.6] days | - | | |
| Days in shock | MD −1.52 [−1.71 to -1.32] days | - | - | | |
| Days in ICU | MD −0.75 [−1.34 to −0.17] days | MD 0.1 [–3.0 to 3.2] days | - | | |
| Days in hospital | MD −0.87 [−2.17 to 0.44] days | MD –3.6 [–7.2 to –0.02] days | MD -1.22 [-2.08 to -0.35] days | | |
| Adverse events<br>Secondary infections<br>Hyperglycaemia | <br>RR 1.05 [0.95 to 1.16]<br>RR 1.11 [1.07 to 1.16] | <br>RR 0.82 [0.67 to 1.02]<br>RR 1.12 [1.01 to 1.24] | <br>-<br>RR 1.49 [1.01 to 2.19] | | |

| | | | |
|---|---|---|---|
| Gastrointestinal bleeding<br>Encephalopathy<br>Neuromuscular weakness | RR 1.09 [0.80 to 1.46]<br>RR 1.99 [0.37 to 10.84]<br>- | RR 0.71 [0.30 to 1.73]<br>-<br>RR 0.85 [0.62 to 1.18] | RR 0.82 [0.33 to 1.62]<br>RR 1.65 [0.88 to 3.08]<br>- |

ARDS: acute respiratory distress syndrome; SR: systematic review; RCTs: randomised clinical trials; mg: milligrams; RR: relative risk; OR: odds ratio; HR: hazard ratio; MD: mean difference; ICU: intensive care unit

#### *4.4 Ethical justification and trial rationale*

Patients with COVID-19 and severe hypoxia (the hallmark of ARDS) are at high risk of death (6, 7). Until recently, the care for these patients was exclusively supportive, including respiratory and circulatory support.

The RECOVERY trial reported a reduction in 28-day mortality with low-dose dexamethasone (6 mg) for hospitalised patients with suspected or confirmed COVID-19. Yet, higher doses of dexamethasone (median 15 mg) may be beneficial in non-COVID-19 ARDS (42), and higher doses were also used in the other trials of corticosteroids in COVID-19 (median dose 12 mg). Also, in a contemporary survey of the COVID STEROID 2 trial sites after the preprint publication of the RECOVERY trial had been published, 95% of sites used steroids in all patients, most often as dexamethasone in doses above 6 mg (median 9.6 mg (IQR 6.0 to 15.0)), a result supported by those of a concomitant survey of clinician's preferences (unpublished results). Taken together, it is unclear which dose of dexamethasone is most beneficial to COVID-19, and clinical equipoise exists among clinicians and researchers.

The present trial will be conducted to the highest of methodological standards with ongoing assessment of the known serious adverse reactions to corticosteroid, including a planned interim analysis. Any serious adverse reactions for single participants and the group of participants receiving higher vs. lower dose of dexamethasone will be assessed and handled. The control group will receive the exact same protocol as in the RECOVERY trial in addition to usual clinical care. We, the COVID STEROID 2 trial group, find the trial justifiable both medically and ethically.

The patients to be enrolled in the COVID STEROID 2 trial cannot consent due to the combination of severe infection and severe hypoxia. COVID-19 with severe hypoxia is a medical emergency that requires immediate interventions including life-supportive interventions. Therefore, we cannot delay enrolment and need to use the consent procedures for emergency research.

Informed consent will be obtained according to national law in the participating countries. In Denmark, we will use the consent procedures for temporarily incompetent patients for all patients enrolled in the COVID STEROID 2 trial (https://www.retsinformation.dk/Forms/R0710.aspx?id=192671). Here, patients will be enrolled after informed consent from a doctor (first trial guardian), who is independent of the trial, who has knowledge of the clinical condition and who is familiar with the trial protocol to such extent that he/she can judge for each patient, if it will be reasonable to enrol the patient in the trial. As soon as possible after enrolment, consent will be obtained from the patient's next of kin and another doctor (second trial guardian). The second trial guardian is also independent of the trial, has knowledge of the clinical condition, and is familiar with the trial protocol to such extent that he/she can judge for each patient, if it will be reasonable to enrol the patient in the trial. Participants, who regain competence, will be asked for informed consent as soon as possible (Appendix 6, 18.6). The process leading to informed consent will follow all applicable regulations. The consenting parties will be provided with written and oral information about the trial allowing them to make an informed decision about participation in the trial. Written information and the consent form will be subject to review and approval by the ethical committee system. The consenting party can at any time, without further explanation, withdraw consent.

#### *4.5 Trial conduct*

The COVID STEROID 2 trial will comply with the published trial protocol, the Helsinki Declaration in its latest version (52), the International Conference on Harmonization on Good-Clinical-Practice (GCP) guidelines (53), General Data Protection Regulation, and national laws (including Databeskyttelsesloven in Denmark). The Management Committee of the trial will oversee the conduct. We have written the protocol in accordance with the Standard Protocol Items: Recommendations for Interventional Trials (SPIRIT) 2013 Statement (54) and will register the trial in the ClinicalTrials.gov and European Union Drug Regulating Authorities Clinical Trials (EudraCT) registries before the enrolment of the first participant. No substantial deviation from the protocol will be implemented without prior review and approval of the regulatory authorities except where it may be necessary to eliminate an immediate hazard to the trial participants. In such case, the deviation will be reported to the authorities within 7 days.

Enrolment will start after the approval by the Ethics Committee, the Danish Medicines Agency and the Capital Region Knowledge Center for Data Compliance (legal department). We will publish the approved protocol online at the Collaboration of Research in Intensive Care's website at www.cric.nu and submit a manuscript with main points of the protocol including description of design, rationale and the detailed statistical analysis plan to a peer-reviewed medical journal.

### 5 Trial objectives

The objective of the *Higher vs. Lower Doses of Dexamethasone in Patients with COVID-19 and Severe Hypoxia – COVID STEROID 2 trial* is to assess the effects 12 mg vs. 6 mg of intravenous dexamethasone on the number of days alive without life-support and other patient-centered outcomes in adult patients with COVID-19 and severe hypoxia. We hypothesise that dexamethasone 12 mg will increase the number of days alive without life support as compared to dexamethasone 6 mg in patients with COVID-19 and severe hypoxia.

### 6 Trial design

The COVID STEROID 2 trial is an investigator-initiated, international, parallel-group, blinded, centrally randomised, stratified, clinical trial.

#### *6.1 Randomisation*

Patients with COVID-19 fulfilling all inclusion criteria and no exclusion criteria will be randomised. The 1:1 randomisation will be centralised and web-based according to the computer-generated allocation sequence list, stratification variable (trial site, the use of invasive mechanical ventilation (y/n), age below 70 years (y/n)), and varying block size at Copenhagen Trial Unit (CTU) to allow immediate and concealed allocation to one of the two intervention groups. The allocation sequence list will exclusively be known to the data manager at CTU and will be unknown the unblinded trial site staff preparing the trial medication (section 6.2), to the clinicians, to the investigators and statistician conducting the analysis. Each trial participant will be allocated a unique screening number.

#### *6.2 Blinding*

We will mask the allocation for the participants, the clinical staff, the trial site staff registering the outcome data, the trial Management Committee, and the trial statistician, who will conduct the analyses with the two intervention groups coded as e.g. 0 and 1. A dedicated team of trial site staff (medical-, pharmacy- or nurse students or pharmacists, research nurses or doctors) who are certified in medicine handling procedures will unblinded prepare the trial medication and perform daily data entry about the administration of the trial medications including any protocol violations. This unblinded team of trial site staff will not be involved in the care of trial participants, outcome assessment, or in the statistical analyses. They will be instructed not to reveal the allocation under any circumstances.

**Trial medication preparation**

We will use shelf-medications from the hospital department's pharmacy for both intervention and control medication. The local trade names used in the COVID STEROID 2 trial are presented in Appendix 9, 18.9.

To ensure blinding, the trial medications will be prepared by the unblinded trial site staff, and the participants and clinical staff will thus remain blinded to the treatment allocation. For each participant, the trial medication will be prepared once daily and administered as a bolus injection.

**Preparation of experimental intervention: dexamethasone 12 mg**

The experimental intervention is dexamethasone 12 mg daily given as bolus injection for up to 10 days. We will allow the use of betamethasone 12 mg at sites where dexamethasone is not available, as these are diastereomers and likely equipotent (55).

Dexamethasone phosphate is a clear colourless solution and comes in vial of 1 and 5 ml (4 mg per ml, which equals 3.33 mg of dexamethasone). For each participant allocated to the experimental intervention, 3.6 ml of dexamethasone phosphate will be drawn into one 5 ml syringe together with 1.4 ml isotonic saline (0.9%) to a total volume of 5 ml and a concentration of 2.88 mg/ml of dexamethasone phosphate, which equals a total of 12 mg of dexamethasone. The trial medication will be delivered daily to the clinical staff by the unblinded trial staff and administered as one bolus injection.

**Preparation of control intervention: dexamethasone 6 mg**

The control intervention is dexamethasone 6 mg daily given as bolus injection for up to 10 days. We will allow the use of betamethasone 6 mg at sites where dexamethasone is not available, as these are diastereomers and likely equipotent (55).

For each participant allocated to the control intervention, 1.8 ml of dexamethasone phosphate (4 mg per ml, which equals 3.33 mg of dexamethasone) will be mixed with 3.2 ml of isotonic saline (0.9%) to a total volume of 5 ml and a concentration of 1.44 mg/ml of dexamethasone phosphate, which equals a total of 6 mg of dexamethasone. The dexamethasone solution will be delivered daily to the clinical staff by the unblinded trial staff and administered as one bolus injection.

#### *6.3 Unblinding*

**Unblinding of the intervention for a participant**

The intervention may be unblinded if deemed necessary by the treating clinician or the investigator for treatment or safety reasons. The sponsor or his delegate will break the blind for a participant if there is clinical suspicion of an unexpected serious adverse reaction (SUSAR) and judge the 'expectedness' of this according to the product information. Any SUSAR will be reported to the authorities accordingly.

Unblinding of the intervention for a participant can be performed around the clock by contacting the sponsor or his delegate. The sponsor or his delegate will contact the unblinded trial site staff from whom the trial allocation is available, and the intervention will be discontinued. The primary investigator at the site will be informed about the participant's allocation.

**Unblinding of the entire trial**

The Management Committee may stop and unblind the trial if there are clear indications that one intervention is superior to the other based on the recommendations from the independent Data Monitoring and Safety Committee (IDMSC) or other relevant data.

The members of the IDMSC will remain blinded unless 1) they request otherwise or 2) the interim analysis has provided strong indications of one intervention being beneficial or harmful.

#### *6.4 Participant timeline*

We will strive to enrol participants as soon as they fulfil the inclusion criteria, and no later than within 5 days of initiation of standard care corticosteroids for COVID-19. The allocated intervention will be continued so that participants in total receives 10 days of corticosteroids or until discharge from the participating site or death (whichever occurs first). Thus, no participant will receive corticosteroid for COVID-19 for more than 10 consecutive days. We will follow the patients for 28 days after randomisation and identify survivors at days 90 and 180 in electronic patient records or in registries. At day 180, we will contact surviving participants or their next of kin for health-related quality of life (HRQoL) follow-up.

**End of trial**

The trial will end when the last patient enrolled has completed 180-day follow up (last-patient last-visit). We will report the end-of-trial no later than 90 days after the last-patient last-visit to the Danish Medicines Agency and Ethics Committee.

### 7 Selection of participants

All patients admitted to an active trial site will be considered for participation. Patients will be eligible if they comply with the inclusion and exclusion criteria (full definitions are presented in Appendix 3, 18.3).

#### *7.1 Inclusion criteria*

All the following criteria must be fulfilled:

- Aged 18 years or above **AND**
- Confirmed SARS-CoV-2 (COVID-19) requiring hospitalisation **AND**
- Use of one of the following:
    - Invasive mechanical ventilation **OR**
    - Non-invasive ventilation or continuous use of continuous positive airway pressure (CPAP) for hypoxia **OR**
    - Oxygen supplementation with an oxygen flow of at least 10 L/min independent of delivery system

#### *7.2 Exclusion criteria*

We will exclude patients who fulfil any of the following criteria:

- Use of systemic corticosteroids for other indications than COVID-19 in doses higher than 6 mg dexamethasone equivalents
- Use of systemic corticosteroids for COVID-19 for 5 days consecutive days or more

- Invasive fungal infection
- Active tuberculosis
- Fertile woman (<60 years of age) with positive urine human gonadotropin (hCG) or plasma-hCG
- Known hypersensitivity to dexamethasone
- Previously randomised into the COVID STEROID 2 trial
- Informed consent not obtainable

We will not exclude patients enrolled in other interventional trials unless the protocols of the two trials collide. We will establish co-enrolment agreements when possible with the sponsor/investigator to maintain an updated list of trials approved for co-enrolment (Appendix 7, 18.7).

#### *7.3 Participant discontinuation and withdrawal*

The procedure for handling withdrawal of consent from a participant will follow national regulations. In Denmark, the procedure will be as follows.

**Discontinuation and withdrawal at the choice of the participant or the proxy**
A participant, who no longer wishes to participate in the trial, can withdraw his/her consent at any time without need of further explanation, and without consequences for further treatment.

For incapacitated participants, consent can be withdrawn at any time by the person(s), who has given proxy-consent. To limit the amount of missing data, we will collect as much data as possible from each participant. Therefore, the investigator will ask the participant or the proxy if they allow continued data registration and follow-up at day 180.

**Discontinuation and withdrawal at the choice of the investigator**
A participant may have the intervention stopped by the clinician or investigator at any time, if:
- The participant experiences intolerable adverse reactions or events (including Serious Adverse Reactions (SARs) or Suspected Unexpected Serious Adverse Reactions (SUSARs)) suspected to be related to the trial intervention.
- The clinicians in conjunction with the coordinating investigator decide it to be in the interest of the participant

- Withdrawal from active therapy
- The participant is subject to compulsory hospitalisation.

In these participants, the collection of data and the follow-up will continue, and the participant will remain in the intention-to-treat population.

**Discharge**

The trial allocation will be stopped when patients are discharged or transferred to a non-participating hospital department. The patient will still be followed through the electronic health records, including registration of data for days alive without life support and day alive and out of hospital. Participants who are discharged or transferred to a department participating in the COVID STEROID 2 trial will continue the allocated intervention at the new trial site for a total treatment duration of 10 days from randomisation. If the participant is readmitted to a COVID STEROID 2 trial site from a non-participating hospital department within 10 days of randomisation, the allocation will also resume for a total treatment duration of up to 10 days from randomisation depending on the number of days with corticosteroid treatment before randomisation.

### 8 Selection of trial sites and personnel

#### *8.1 Trial sites and setting*

Trial sites will be hospitals in Denmark, Sweden, Switzerland and India. Trial sites are listed in the section *Administrative information* (p. 4). This section will be updated during the trial, and authorities will be notified.

#### *8.2 Trial personnel*

All clinical staff caring for patients will be eligible to care for and give the interventions to the trial participants. The primary trial personnel are constituted of a dedicated team of medical-, pharmacy- or nurse students or research nurses or doctors who will be trained and certified in all trial-related procedures. The screening will be done by the clinical doctors. When a candidate patient is identified, the clinical team will alert the trial staff, who will seek consent and thereafter screen the patient in the eCRF.

Medical students will be eligible to screen and enrol patients in the eCRF, obtain informed consent, prepare trial medication and perform data entry. Nurse and pharmacy students and pharmacists will be eligible to obtain informed consent, prepare trial medication and perform data entry; nurse- and pharmacy students and pharmacists can only screen and enrol of patients in the eCRF if a named doctor or medical student checks and signs the inclusion notes. All participating trial sites will receive written and oral instructions about the trial procedures. A 24-hour hotline will be available for trial-related questions.

#### *8.3 Trial interventions*

The intervention period is up to 10 days from randomisation or until hospital discharge or death, whichever comes first. The intervention period will be adjusted for each participant so that the number of consecutive days with the use of corticosteroid for COVID-19 before randomisation is subtracted from the 10-day intervention period (e.g. a participant who has received corticosteroid for COVID-19 for 3 consecutive days prior to randomisation will receive 7 days of the trial intervention).

#### *8.4 Experimental intervention*

Intravenous bolus injection of dexamethasone 12 mg. We will allow the use of betamethasone 12 mg at sites, where dexamethasone is not available.

#### *8.5 Control intervention*

Intravenous bolus injection of dexamethasone 6 mg. We will allow the use of betamethasone 6 mg at sites, where dexamethasone is not available.

#### *8.6 Co-interventions*

All participants in the trial will be given co-interventions at discretion of the treating clinicians. We will recommend against the use of additional corticosteroids (systemically or as inhalation) and other anti-inflammatory agents (e.g. IL-6 inhibitors) in all trial participants.

Based upon an updated critical appraisal of the literature, the Management Committee endorses and encourages co-enrolment in the COVID STEROID 2 trial (Appendix 7, 18.7). Co-enrolment agreements will be established when possible with the sponsor/investigator to maintain an updated list of trials approved for co-enrolment (Appendix 7, 18.7).

#### *8.7 Concomitant interventions*

All other interventions will be allowed as per the clinical team including those affecting CYP3A4, because it is not clinical practice at the trial sites to change the use or dosing of dexamethasone or betamethasone with concomitant use of CYP3A4 inhibitors or inducers.

#### *8.8 Monitoring of participants*

The participant will be monitored closely due to the severity of their illness. The level of monitoring will be as per the clinical standard of the trial sites including continuous monitoring of oxygen saturation and pulse when severe hypoxia is present; 1-2 hourly measurements of blood pressure and respiratory rate when severe hypoxia is present; and 8-hourly measurement of body temperature; daily measurement of blood values including C-reactive protein (CRP), leukocyte count, hemoglobin, creatinine, urea and electrolytes, pH, atrial blood gases, lactate, and blood glucose. Additional measurements will be done on clinical indications including microbiological cultures, markers of candida infections and electrocardiograms (ECGs).
These data will not be registered in the COVID STEROID 2 trial eCRF but will be available in the participant's health care records for the Sponsor and/or the authorities if needed.

#### *8.9 Criteria for modification of interventions for a given trial participant*

The clinical team may at any time violate the protocol if they find it to be in the best interest of the participant. We will have a COVID STEROID 2 trial hotline to enable discussion around-the-clock between the clinicians caring for trial participants and the COVID STEROID 2 trial team regarding protocol related issues. Protocol violations will be registered and reported.

#### 8.10 Assessment of participant compliance

We will monitor protocol compliance at the trial site through the electronic case report form (eCRF) and alert trial sites in the case of clear violations (central monitoring). In addition, the trial will be externally monitored according to the GCP Directive and the monitoring plan (section 13).

#### 8.11 Intervention accountability

Both the trial intervention and control medications are routinely used for in-hospital treatment of patients and we will use shelf-medication from the department's pharmacy. The trial medication will only be handled by the trained trial staff and the clinical staff who are trained and certified for the caring for patients. The methods used for trial medication preparations are described in 6.2.

**Trial medications**

The list of local brands used in Denmark, Sweden, Switzerland and India are presented in Appendix 18.9.

<u>Experimental intervention</u>

Active medication: dexamethasone phosphate, solution for injection, 4 mg/ml, ATC code: H02AB02

At sites where dexamethasone is not available: betamethasone sodium phosphate, solution for injection, 5.3 mg/ml, ATC code: H02AB01

Solution: isotonic saline, solution for intravenous injection, 9mg/ml, ATC code: B05BB01. Each ml contains 9 mg of saline in sterile water. Content of electrolytes/l: 154mmol chloride, 154 mmol natrium. Osmolarity 308mmol/l.

<u>Control intervention</u>

Active medication: dexamethasone phosphate, solution for injection, 4 mg/ml ATC code: H02AB02

At sites where dexamethasone is not available: betamethasone sodium phosphate, solution for injection, 5.3 mg/ml, ATC code: H02AB01

Solution: isotonic saline, solution for intravenous injection, 9mg/ml, ATC code: B05BB01. Each ml contains 9 mg of saline in sterile water. Content of electrolytes/l: 154mmol chloride, 154 mmol natrium. Osmolarity 308mmol/l.

**Labelling**

When the trial drug is prepared, it will be labelled with a COVID STEROID 2-trial sticker, making clinical personnel aware that the syringe contains trial medication. The sticker will hold information about the participant's data, the trial medicines, the date and time of preparation, the expire date and time, the signature of the trial staff preparing the medications and a telephone number for the COVID STEROID 2-trial 24-h hotline (the labels are presented in Appendix 4, 18.4). To ensure blinding of the clinicians, the sticker will not hold information about the BATCH / LOT numbers of the trial medications. The BATCH / LOT numbers will instead be noted in a trial medication log. This log will only be available to the unblinded research staff.

### 9 Outcome measures

#### *9.1 Primary outcome*

Days alive without life support (i.e. invasive mechanical ventilation, circulatory support or renal replacement therapy (including days in between intermittent renal replacement therapy)) from randomisation to day 28.

#### *9.2 Secondary outcomes*

- Number of participants with one or more serious adverse reactions (SARs) at day 28 defined as new episodes of septic shock, invasive fungal infection, clinically important gastrointestinal (GI) bleeding or anaphylactic reaction to IV dexamethasone
- All-cause mortality at day 28
- All-cause mortality at day 90
- Days alive without life support at day 90
- Days alive and out of hospital at day 90
- All-cause mortality at day 180

- HRQoL at day 180 using EQ-5D-5L and EQ-VAS

### 10 Safety

#### *10.1 Definitions*

In the COVID STEROID 2 trial, we will use the definitions below (56):

**Adverse event (AE)**

Any undesirable medical event occurring to a participant during a clinical trial, which does not necessarily have a causal relationship with the intervention.

**Adverse reaction (AR)**

Any undesirable and unintended medical response related to the intervention occurring to a participant during a clinical trial.

**Serious adverse event (SAE)**

Any adverse event that results in death, is life-threatening, requires hospitalisation or prolongation of existing hospitalisation, results in persistent or significant disability or incapacity, or is a congenital anomaly or birth defect.

**Serious adverse reaction (SAR)**

Any adverse reaction that results in death, is life-threatening, requires hospitalisation or prolongation of existing hospitalisation, results in persistent or significant disability or incapacity, or is a congenital anomaly or birth defect. The SARs are identified in the Danish Summary of Products Characteristics (SmPC) for dexamethasone.

**Suspected unexpected serious adverse reaction (SUSAR)**

Any suspected adverse reaction which is both serious and unexpected (the nature or severity of which is not consistent with SmPC for dexamethasone).

#### 10.2 Risk and safety issues in the COVID STEROID 2 trial

The trial participants will be hospitalised patients for whom adverse events and reactions are documented routinely in the patient health record (i.e. notes, charges and laboratory reports). We will record the occurrence of SARs in the 28 days following randomisation for all participants and report them as an outcome measure.

For all participants, we will register daily the presence or absence of potential SARs according to intravenous dexamethasone in the Danish SmPC, which are serious and relevant to short course use in critically ill patients, i.e. new episodes of septic shock, invasive fungal infections, clinically important GI bleeding and anaphylaxis.

#### 10.3 Assessment of adverse events

**Timing**

In all participants, we will assess the occurrence of SARs in the 28 days following randomisation (the maximum intervention period is 10 days; 28 days allow for at least another 18 days of assessment after the intervention, which is clinically relevant in short course use in critically ill patients.

**Classification of an event**

We will make no inferences about a causal relationship between the intervention and the SARs but register the occurrence in the two groups and report them in the final report according to the definition given above.

As for any SAE, the investigators will report them to the sponsor or his delegate within 24 hours. If such a SAE is deemed both unexpected and related to the intervention by the investigator, it will be considered a SUSAR and reported as such. If the sponsor does not adjudicate the SAE as related to the intervention, this will also be noted in the SUSAR report.

**Reporting**

Any SAE adjudicated to be related to the trial intervention by the investigator, will be reported within 24 hours to the Sponsor or his delegate. If deemed a SUSAR by the sponsor, he will report it to the Danish Medicine Agency, the Ethics Committee and all trial sites within 7 days. No later than 8 days after the reporting, the Sponsor will inform the Danish Medicines Agency of relevant information on the Sponsor's and the investigator's follow-up action to the life-threatening or fatal SUSAR. Any other SUSARs will be reported to the Danish Medicines Agency no later than 15 days from the time when the Sponsor is informed.

Once a year, the sponsor will submit a list of all SARs that have occurred at all sites during the trial period and a report on safety of the trial subjects to the Danish Medicines Agency and National Ethics Committee.

The sponsor will notify the Danish Medicines Agency when the trial has been completed (no later than 90 days thereafter) and if earlier than planned, the reasons for stopping the trial.

In addition, we will report all SARs defined in 9.2 as outcome measures and all SUSARs in the final trial report and the results of the trial will be reported on EudraCT within 12 months of 'last-patient-last-visit'.

### 11 Procedures, assessments and data collection

#### *11.1 Screening*

All patients admitted to a participating trial site with confirmed COVID-19 and severe hypoxia (as defined in section 7.1) will be eligible for screening. The screening will be done by the clinical doctors. When a candidate patient is identified, the clinical team will alert the trial staff, who will seek consent and thereafter screen the patient in the eCRF.

For all fertile women under 60 years of age, screening for hCG in urine or plasma will be done before enrolment in the trial. If a hCG-test has already been done under the current admission, we will use the test result of this for screening for pregnancy.

#### *11.2 Procedures of informed consent*

Participants will be enrolled after consent by proxy is obtained according to national regulations. We will follow the normal procedures for collection of informed consent and any modifications to these as approved by the Ethics Committee due to restrictions in physical visits to the hospitals during the current SARS-CoV-2 epidemic. The procedure for informed consent in Denmark is described in Appendix 6 (18.6).

#### *11.3 Data collection*

The screening of participants will be done by the clinical team as described in 10.1. The clinicians will pass on information about eligible participants to the COVID STEROID 2 trial site staff who will hereafter obtain informed consent from the first trial guardian (the Danish procedure).

After informed consent is obtained, the data below (10.4) will be obtained by the trial site staff from the participant’s hospital files, national/regional/hospital registers (source data as defined per site and region) and interview with participant or next of kin and entered into the web-based eCRF (the server hosting the database is located at CTU, Rigshospitalet, Region Hovedstaden). For participants transferred from a trial site to a non-trial site, data related to the outcomes will be collected from either hospital files (if accessible) or investigator contact to the non-trial site or health care registers.

#### *11.4 Variables*

All variables are defined in Appendix 3 (18.3).

**Screening variables**

Inclusion and exclusion criteria (7.2 and 7.3)

Number of consecutive days of systemic use of corticosteroids for COVID-19 up to the day of screening

**Baseline variables**

- Sex
- Age at enrolment (date of birth)
- Date of admission to hospital
- Number of days with symptoms before hospital admission
- Department at which the participant was included:
    - Emergency department
    - Hospital ward
    - Intermediate care unit
    - Intensive care unit
- Use of respiratory support at randomisation:
    - Closed system (y/n): Invasive mechanical ventilation or non-invasive ventilation or continuous use of CPAP
        - If yes, latest FiO2 prior to randomisation
        - If yes, no. of days of mechanical ventilation prior to randomisation
    - Open system with an oxygen flow ≥10 L/min (y/n)
        - If yes, maximum supplemental oxygen flow on an open system at randomisation (+/- 1 h)
- Limitations of care (i.e. not for invasive mechanical ventilation, circulatory support, renal replacement therapy, cardio-pulmonary resuscitation) at the time of randomisation (y/n)
- Chronic use of systemic corticosteroids for other indications than COVID-19 (y/n)
- Treatment for COVID-19 during current hospital admission prior to randomisation:
    - Agents with potential anti-viral action:
        - Remdesivir
        - Convalescent plasma
        - Other
    - Anti-bacterial agent (y/n)
    - Agents with potential anti-inflammatory action:
        - Janus kinase inhibitor(y/n)
        - IL-6 inhibitors (y/n)

- Other

- Chronic co-morbidities:
  - History of ischaemic heart disease or heart failure (y/n)
  - Diabetes Mellitus (y/n)
  - Chronic pulmonary disease (y/n)
  - Immunosuppressive therapy within the last 3-months (y/n)

- Blood values, interventions and vital parameters:
  - Participant weight
  - $PaO_2$ and $SaO_2$ in the most recent arterial blood gas sample prior to inclusion **OR** $SpO_2$ from pulse oximeter if arterial blood gas sample is not available
  - Circulatory support (infusion of vasopressor/inotropes) within the last 24 hours prior to randomisation (y/n)
  - Renal replacement therapy within the last 72 hours prior to randomisation (y/n)
  - Highest plasma lactate within the last 24 hours prior to randomisation

**Daily during admission for the first 14 days after randomisation (day forms)**

- Invasive mechanical ventilation (y/n)

- Circulatory support (continuous infusion of vasopressor/inotropes for a minimum of 1 hour) on this day (y/n)

- Any form of renal replacement therapy on this day including days between intermittent renal replacement therapy (y/n)

- SAR(s) on this day (y/n for each)
  - New episodes of septic shock
  - Invasive fungal infection
  - Clinically important GI bleeding
  - Anaphylactic reaction to IV dexamethasone

**Daily registration of major protocol violations up to 10 days (from day 1 and up to 10 days)**

- Use of open-label systemic corticosteroids on this day (y/n)

-Trial intervention (y/n): did the participant receive the trial medication on this day? (yes/no)

- If no, apply reasons: by error/lack of resources, other reason

**Discharge form**

- Died in hospital

- Discharged from hospital

- Discharged to another ward participating in the COVID STEROID 2 trial

- Discharged to another ward not participating in the COVID STEROID 2 trial

**Follow-up 28 days after randomisation**

- Death (y/n, if yes: date of death)
- Number of days on invasive mechanical ventilation from day 15-28
- Number of days with circulatory support (infusion of vasopressor/inotropes for a minimum of 1 hour) from day 15-28
- Number of days on renal replacement therapy from day 15-28
- The occurrence of SAR(s) from day 15-28:
  - New episodes of septic shock (y/n, if yes: apply date(s))
  - Invasive fungal infection (y/n, if yes: apply date(s))
  - Clinically important GI bleeding (y/n, if yes: apply date(s))
  - Anaphylactic reaction to IV dexamethasone (y/n, if yes: apply date(s))
- Use of extracorporeal membrane oxygenation (ECMO) from randomisation to day 28 (y/n)
- Discharged against medical advice to home/other hospital/other facility (y/n)
  - If yes, apply medical condition at the time of discharge: on life support (i.e. invasive mechanical ventilation, circulatory support or renal replacement therapy), receiving supplementary oxygen (<10 L/min or ≥10 L/min) or no supportive therapy

**Follow-up 90 days after randomisation**

- Death (y/n, if yes date of death)
- Number of days on invasive mechanical ventilation from day 29-90
- Number of days with circulatory support (continuous infusion of vasopressor/inotropes for a minimum of 1 hour) from day 29-90
- Number of days on renal replacement therapy from day 29-90, including days between intermittent renal replacement therapy
- Date of discharge from hospital
- Additional hospital admissions (y/n, if yes: date of re-admission(s) and discharge(s))

**Follow-up 180 days after randomisation**

- Death (y/n, if yes date of death)

- HRQoL

- EQ-5D-5L
- EQ-VAS

### 12 Statistical plan and data analysis

The analyses will be done according to the principles stipulated in ICH-GCP guidelines (56). The protocol and detailed statistical analysis plan will be published online at www.cric.nu and in a peer-reviewed journal before the conduct of the planned interim analysis.

The primary outcome will be compared using the Kryger Jensen and Lange test and reported as differences in means and medians along with 95% confidence intervals adjusted for the stratification variables (site, use of invasive mechanical ventilation, and age). Secondary binary outcomes will be analysed using binomial regression models with log links adjusted for the stratification variables (site, use of invasive mechanical ventilation, and age) with results quantified as adjusted relative risks supplemented with adjusted risk differences, both with 95% confidence intervals.

#### *12.1 Sample size and power*

**Sample size estimation and testing strategy**

At maximum, we will randomise 1,000 participants. A blinded statistician will conduct an interim analysis after the first 500 participants have been followed for 28 days. The alpha values for the interim analysis and the final analysis are 0.0054 and 0.0492, respectively as by the O'Brien-Fleming bounds, which preserves type I error at the usual 5% (57). In both analyses, a Kryger Jensen and Lange test will be employed to compare the groups on the primary outcome. The trial will be stopped early if the alpha cut-off is crossed at the interim analysis. The trial has 85% power to detect a 15% relative reduction in 28-day mortality combined with a 10% reduction in time on life support among the survivors.

The mortality outcomes will be tested in a hierarchical procedure along with the primary outcome (first primary outcome, then 28 days mortality, and finally 90 days mortality) reusing the alpha if the previous test was significant. If the primary outcome is insignificant at trial conclusion, ordinary 5% level test will be employed for all additional outcomes, but the results interpreted as exploratory.

**Power estimations for secondary outcomes**

We expect to have 80% statistical power to detect the following effects for the secondary outcomes based on the trial design described above. Power is reported at the 5% level even though the two mortality outcomes are also part of the primary outcome's hierarchical testing procedure.

- A 21% relative risk reduction for the mortality at day 28 (control event rate 30%)

- A 18% relative risk reduction for the mortality at day 90 (control event rate 40%)

- A 32% relative risk reduction for the number of participants with one or more SARs (control event rate 15%)
- A 15% relative risk reduction for the mortality at day 180 (control event rate 50%)

The estimates of control event rates for mortality at day 28 originate in data of previous coronavirus studies (6, 58); the estimates of the control event rates for mortality at day 90 and the number of participants with SARs are based on best clinical estimate. We expect the following secondary outcomes to be highly skewed (non-normally distributed): Days alive out of hospital at day 90 and HRQoL at 180 days. The power estimations for these are, therefore, somewhat uncertain why we refrain from making these estimates.

#### *12.2 Statistical methods*

The analyses will be done in the intention-to-treat (ITT) population defined as all randomised participants for whom there is consent for the use of data.

The primary outcome will be compared using a Kryger Jensen and Lange test adjusted for the stratification variables (site, invasive mechanical ventilation, and age). Differences will be quantified as differences in means and medians along with 95% confidence intervals. For the binary outcomes (including mortality outcomes), we will use generalised linear models with log links and binomial error distributions adjusted for the stratification variables (site, invasive mechanical ventilation, and age) as the primary analysis (59). Differences in binary outcomes will be quantified using adjusted relative risks and secondarily adjusted risk differences along with 95% confidence intervals. This will be supplemented with Fisher's exact tests. Days alive without life support at day 90 and days alive out of hospital at day 90 will be analysed similarly to the primary outcome.

We will challenge the primary result in analyses adjusted for important baseline risk factors (age, co-morbidities, and use of life-support), in subgroups (Table 2) and in the per-protocol population being the ITT population except those having one or more major protocol violations as defined above (11.4 Variables). If there are more than 5% missing data for outcomes and/or covariates for an analysis, we will multiply impute the missing data for that analysis.

All statistical tests will be 2-tailed. Several outcome measures (including SARs and days alive without the use of life support at day 28 and 90) are composite; we will also report each component of these outcomes as recommended (56) in a supplement to the main report.

*Table 2. Heterogeneity of the intervention effects on the primary outcome will be analysed in the following subgroups based on baseline characteristics. As statistical test, we will use test of interaction in the adjusted analysis described above (p = 0.01).*

| **Subgroup** | **Definition** | **Expected direction of the interaction** |
|---|---|---|
| Elderly patients | Patients ≥70 years versus <70 years of age | Larger beneficial effect of higher dose dexamethasone in the younger population |
| Invasive mechanical ventilation | Patients who receive invasive mechanical ventilation versus oxygen by other delivery systems | Larger beneficial effect of higher dose dexamethasone in patients who receive invasive mechanical ventilation |
| Shock | Patients with shock versus without shock | Larger beneficial effect of higher dose dexamethasone in patients with shock |

| | | |
|---|---|---|
| Duration of corticosteroid use before enrolment in COVID STEROID 2 trial | Patients who received corticosteroids for COVID-19 for 0 to 2 days versus 3 to 4 days before enrolment | Larger beneficial effect of higher dose dexamethasone in patients with short duration (≤2 days) of corticosteroid use before enrolment |
| Limitations of care | Patients with limitations of care (i.e. not for invasive mechanical ventilation, circulatory support, renal replacement therapy, cardio-pulmonary resuscitation) versus without limitations of care | Larger beneficial effect of higher dose dexamethasone in patients without limitations of care |
| Chronic use of systemic corticosteroids for other indications than COVID-19 | Patients with versus without chronic use of systemic corticosteroids | Larger beneficial effect of higher dose dexamethasone in patients without chronic use of systemic corticosteroids |

**Effect measures**

We will present the effects on the primary outcome as raw mean differences as well as median differences. For binary outcomes, we will report results as raw and adjusted relative risks and absolute risk differences, computed using generalized linear models (GLMs) with appropriate link functions (log links) and binomial error-distribution. Results will be presented with 95% confidence intervals (CI) for the analyses of the primary outcome (P-value 0.05) and 99% CIs for those of the secondary outcomes (P-value 0.01) due to the multiplicity of these. Significance of results will be based on the test described under testing strategy.

**Interim analysis**

We will conduct one interim-analyses after 500 participants have been followed for 28 days.

The IDMSC will analyse the primary outcome and the occurrence of SARs as described in the charter (Appendix 5, 18.5). The IDMSC will submit their recommendations to the Management Committee, which make the final decision regarding the continuing, pausing or stopping of the trial as described in the IDMSC charter.

After completion of the interim analysis, the recommendations from the IDMSC and the conclusion reached by the Management Committee will be submitted to the Ethics Committee.

**Early stopping criteria**

We will employ O'Brien-Fleming bounds which imply a significant cut-off of 0.0054 at the interim analysis. The Kryger Jensen and Lange test will be employed to compare the groups for the primary outcome. The trial will be stopped early if the alpha cut-off is crossed at the interim analysis.

**Final analysis**

Before unblinding the interventions groups, we will submit the statistical report of primary and secondary outcomes at day 28 (i.e. days alive without life support, mortality and SAR) to the IDMSC. The IDMSC will be asked to submit their recommendations to the Management Committee on whether to submit a primary report on 28-day outcomes or await the analyses of 90-day outcomes.

### 13 Quality control and quality assurance

The sponsor and his delegates will be responsible for organising the trial sites including education of the local investigators, the trial site staff and clinical staff before the initiation of the trial. This education will be continuously documented in the site master file.

After initiation, trial site investigators will be responsible for all trial-related procedures at their site, including education of staff in trial-related procedures, recruitment and follow-up of participants and entry of data. Clinical staff at the trial sites will be responsible for screening of eligible patients and the treatment of trial participants.

#### 13.1 Monitoring

The trial will be externally monitored according to the GCP Directive and the monitoring and data verification plan including the documentation of informed consent of trial participants. The monitoring and data verification plan will be developed together with the GCP unit of Copenhagen University Hospital and adhered to by the staff monitoring all trial sites.

After the consent is obtained, Sponsor and his delegates will have access to the participants hospital files for quality control and monitoring. Sponsor will allow direct access to source data for GCP monitoring or control visits by the Danish national authorities overseeing drug trials. In addition, we will use central monitoring of site through the eCRF, including adherence to the protocol.

#### 13.2 Drug traceability measures

The registration of the batch numbers and the expiry dates of the dexamethasone and saline used, and the identity of the clinician administering the dexamethasone and saline will be registered as per standard practice at the sites. These data will not be registered in the trial documents but can be obtained by the Sponsor or the authorities if needed. We believe that this is a safe procedure because both the dexamethasone and saline used in the COVID STEROID 2 trial has been in clinical use for many years and the safety of single doses cannot be questioned. The same procedure was approved by the Danish Medicines Agency in the CLASSIC (EudraCT no. 2018-000404-42) and COVID STEROID trials (EudraCT no. 2020-001395-15).

### 14 Legal and organisational aspects

#### 14.1 Finance

##### Trial funding

The trial is funded by grants from the Novo Nordisk Foundation (DKK 5.000.000,-) and Rigshospitalet (DKK 1.875.000,-). The funding organisation has not been or will not be involved in the design, conduct, analyses, or reporting of the trial nor will it have ownership of the data. The Sponsor and trial staff have no financial affiliations to the Novo Nordisk Foundation.

##### Compensation

Dependent on the workload and preferences, the trial sites will receive case money or funds to the salary for the dedicated team of trial staff.

**Insurance**

In Denmark, the trial participants are covered by the Danish Law ‘Lov om Patientskadeerstatning’; in Sweden, by the ‘Läkemedelsförsäkringen’; and in Switzerland, by the participating hospital’s insurance. In India, insurance will be covered by the local sponsor (The George Institute for Global Health, India).

### 15 Plan for publication, authorship and dissemination

All trial results, whether positive, negative or neutral, will be published preferably in a peer-reviewed medical journal. Furthermore, the results will be published at the Collaboration for Research in Intensive Care (CRIC) home page (www.cric.nu). We will adhere to the Consolidated Standards of Reporting Trials (CONSORT) statement (60), including the accountability of all patients screened (Appendix 2, 18.2).

Before unblinding the intervention groups, the Management Committee will write two abstracts based on the statistical report with the group allocation masked, one assuming the experimental intervention group is X and the control intervention group is Y, and one assuming the opposite. Then, the allocation code will be unmasked.

Authorship will be granted according to the guidelines from the International Committee for Medical Journal Editors (ICMJE; http://www.icmje.org/recommendations/browse/roles-and-responsibilities/defining-the-role-of-authors-and-contributors.html). The listing of authors will be as follows on the primary publication: MW Petersen will be first author, SN Myatra the second, and BKT Vijayaraghavan the third author. The next authors will be the site investigators according to the number of included participants per site, and then the other members of the Management Committee. A. Perner will be the last and corresponding author.

The Management Committee may grant additional authorships depending on personal input as per the Vancouver definitions. Investigators on sites may be granted authorship on sub-study publications if they contribute significantly as per the Vancouver definitions.

The IDMSC and investigators not qualifying for authorship will be acknowledged with their names under ‘the COVID STEROID 2 trial investigators’ in an *appendix* to the final manuscript.

The funding sources will be acknowledged, but they will have no influence on the data handling or analyses, the writing of the manuscript or the decision to publish.

##### 15.1 Sub-studies

Sub-studies will be encouraged if they do not hamper the completion of the main protocol and can be conducted after approval of the specific protocol by the Management Committee and the authorities. Thus, specific protocols for any sub-studies will be submitted to and approved by the relevant authorities and ethic committees before the commencement of such studies. In Appendix 8 (18.8), any proposed sub-studies are listed.

##### 15.2 Intellectual property rights

The COVID STEROID 2 trial group owns the trial data.

##### 15.3 Organisational framework

The COVID STEROID 2 trial will be conducted and managed by the Sponsor, Management Committee, (Appendix 1, 18.1), the dedicated trial site team, the investigators, and the Research Unit at Department of Intensive Care, Rigshospitalet.

#### 16 Estimated trial timeline

- August 2020, authority approvals and 1st participant randomised

- December 2020, interim analysis

- Mid 2021, last participant randomised and primary report on 28-day outcomes submitted.

- Late 2021, report on 90-day outcomes submitted

- Mid 2022, report on 180-day outcomes submitted

25. Efficacy of Dexamethasone Treatment for Patients With ARDS Caused by COVID-19 (DEXA-COVID19) (NCT04325061). Available from: https://clinicaltrials.gov/ct2/show/NCT04325061?term=COVID+and+dexa&draw=2&rank=1.
26. Community-Acquired Pneumonia : Evaluation of Corticosteroids (CAPE_COD) (NCT02517489). Available from: https://clinicaltrials.gov/ct2/show/NCT02517489?term=COVID+hydrocortisone&draw=2&rank=5.
27. Efficacy and Safety of Corticosteroids in COVID-19 (NCT04273321). Available from: https://clinicaltrials.gov/ct2/show/NCT04273321.
28. Dexamethasone Treatment for Severe Acute Respiratory Distress Syndrome Induced by COVID-19 (DHYSCO) (NCT04347980). Available from: https://clinicaltrials.gov/ct2/show/NCT04347980.
29. Dexamethasone for COVID-19 Related ARDS: a Multicenter, Randomized Clinical Trial (NCT04395105). Available from: https://clinicaltrials.gov/ct2/show/NCT04395105.
30. Dexamethasone and Oxygen Support Strategies in ICU Patients With Covid-19 Pneumonia (COVIDICUS) (NCT04344730). Available from: https://clinicaltrials.gov/ct2/show/NCT04344730.
31. Low-dose Hydrocortisone for Patients with COVID-19 and Severe Hypoxia (NCT04348305). Available from: https://clinicaltrials.gov/ct2/show/NCT04348305?term=COVID+STEROID&draw=2&rank=1. .
32. COVID-19-associated ARDS Treated With Dexamethasone: Alliance Covid-19 Brasil III (CoDEX) (NCT04327401). Available from: https://clinicaltrials.gov/ct2/show/NCT04327401.
33. Efficacy and Safety of Corticosteroids in Oxygen-dependent Patients With COVID-19 Pneumonia (CORTICOVIDHUGO) (NCT04359511). Available from: https://clinicaltrials.gov/ct2/show/NCT04359511.
34. Corticosteroids During Covid-19 Viral Pneumonia Related to SARS-Cov-2 Infection (CORTI-Covid) (NCT04344288). Available from: https://clinicaltrials.gov/ct2/show/NCT04344288
35. University of Oxford. Press release on 16 June 2020: Low-cost dexamethasone reduces death by up to one third in hospitalised patients with severe respiratory complications of COVID-19. Available from: https://www.recoverytrial.net/files/recovery_dexamethasone_statement_160620_v2final.pdf. Accessed on 16 June 2020.
36. Minneci PC, Deans KJ, Eichacker PQ, Natanson C. The effects of steroids during sepsis depend on dose and severity of illness: an updated meta-analysis. Clin Microbiol Infect. 2009;15(4):308-18.

37. Lamontagne F, Rochwerg B, Lytvyn L, Guyatt GH, Moller MH, Annane D, et al. Corticosteroid therapy for sepsis: a clinical practice guideline. BMJ. 2018;362:k3284.
38. Lin LL, Gu HY, Luo J, Wang L, Zhang C, Niu YM, et al. Impact and Beneficial Critical Points of Clinical Outcome in Corticosteroid Management of Adult Patients With Sepsis: Meta-Analysis and GRADE Assessment. Front Pharmacol. 2019;10:1101.
39. Sun S, Liu D, Zhang H, Zhang X, Wan B. Effect of different doses and time-courses of corticosteroid treatment in patients with acute respiratory distress syndrome: A meta-analysis. Exp Ther Med. 2019;18(6):4637-44.
40. COVID-19 Treatment Guidelines Panel. Coronavirus Disease 2019 (COVID-19) Treatment Guidelines. National Institutes of
Health. Available at https://www.covid19treatmentguidelines.nih.gov/. Accessed 01 June 2020.
41. Bhimraj A, Morgan RL, Shumaker AH, Lavergne V, Baden L, Cheng VC, et al. Infectious Diseases Society of America Guidelines on the Treatment and Management of Patients with COVID-19. Available from: https://www.idsociety.org/globalassets/idsa/practice-guidelines/covid-19/treatment/idsa-covid-19-gl-tx-and-mgmt-v2.1.0.pdf. Accessed on July 10, 2020. 2020.
42. Villar J, Ferrando C, Martinez D, Ambros A, Munoz T, Soler JA, et al. Dexamethasone treatment for the acute respiratory distress syndrome: a multicentre, randomised controlled trial. Lancet Respir Med. 2020;8(3):267-76.
43. Rezk NA, Ibrahim AM. Effects of methyl prednisolone in early ARDS. Egyptian Journal of Chest Diseases and Tuberculosis. 2012;Volume 62, Issue 1:167-72.
44. Liu L, Li J, Huang YZ, Liu SQ, Yang CS, Guo FM, et al. [The effect of stress dose glucocorticoid on patients with acute respiratory distress syndrome combined with critical illness-related corticosteroid insufficiency]. Zhonghua Nei Ke Za Zhi. 2012;51(8):599-603.
45. Steinberg KP, Hudson LD, Goodman RB, Hough CL, Lanken PN, Hyzy R, et al. Efficacy and safety of corticosteroids for persistent acute respiratory distress syndrome. N Engl J Med. 2006;354(16):1671-84.
46. Meduri GU, Golden E, Freire AX, Taylor E, Zaman M, Carson SJ, et al. Methylprednisolone infusion in early severe ARDS: results of a randomized controlled trial. Chest. 2007;131(4):954-63.
47. Zhao WB, Wan SX, Gu DF, Shi B. Therapeutic effect of glucocorticoid inhalation for pulmonary fibrosis in ARDS patients. Medical Journal of Chinese People's Liberation Army. 2014;Vol. 39, No. 9.

48. Tongyoo S, Permpikul C, Mongkolpun W, Vattanavanit V, Udompanturak S, Kocak M, et al. Hydrocortisone treatment in early sepsis-associated acute respiratory distress syndrome: results of a randomized controlled trial. Crit Care. 2016;20(1):329.
49. Fleischhack G, Hartmann C, Simon A, Wulff B, Havers W, Marklein G, et al. Meropenem versus ceftazidime as empirical monotherapy in febrile neutropenia of paediatric patients with cancer. The Journal of antimicrobial chemotherapy. 2001;47(6):841-53.
50. Rygard SL, Butler E, Granholm A, Moller MH, Cohen J, Finfer S, et al. Low-dose corticosteroids for adult patients with septic shock: a systematic review with meta-analysis and trial sequential analysis. Intensive Care Med. 2018;44(7):1003-16.
51. Siemieniuk RA, Meade MO, Alonso-Coello P, Briel M, Evaniew N, Prasad M, et al. Corticosteroid Therapy for Patients Hospitalized With Community-Acquired Pneumonia: A Systematic Review and Meta-analysis. Ann Intern Med. 2015;163(7):519-28.
52. World Medical Association. WMA Declaration of Helsinki - Ethical Principles for Medical Research involving Human Subjects. Available from: https://www.wma.net/policies-post/wma-declaration-of-helsinki-ethical-principles-for-medical-research-involving-human-subjects/20132013/. Accessed on Feb 26, 2020. 2013.
53. European Medicines Agency. Science Medicines Health. Good Clinical Practice. Available from: https://www.ema.europa.eu/en/human-regulatory/research-development/compliance/good-clinical-practice#international-clinical-trials-section. Accessed on March 18 2020.
54. Chan AW, Tetzlaff JM, Altman DG, Laupacis A, Gotzsche PC, Krleza-Jeric K, et al. SPIRIT 2013 statement: defining standard protocol items for clinical trials. Ann Intern Med. 2013;158(3):200-7.
55. Corticosteroids. LiverTox: Clinical and Research Information on Drug-Induced Liver Injury. Bethesda (MD)2012.
56. ICH Steering Committee. International conference on harmonisation of technical requirements for registration of pharmaceuticals for human use. ICH Harmonised Tripartite. Guideline for Statistical Principles for Clinical Trials re.
57. O'Brien PC, Fleming TR. A multiple testing procedure for clinical trials. Biometrics. 1979;35(3):549-56.
58. Cao B, Wang Y, Wen D, Liu W, Wang J, Fan G, et al. A Trial of Lopinavir-Ritonavir in Adults Hospitalized with Severe Covid-19. N Engl J Med. 2020.
59. Kahan BC, Morris TP. Improper analysis of trials randomised using stratified blocks or minimisation. Stat Med. 2012;31(4):328-40.

### 18 Appendices

#### *18.1 Appendix 1: Trial organisation diagram*

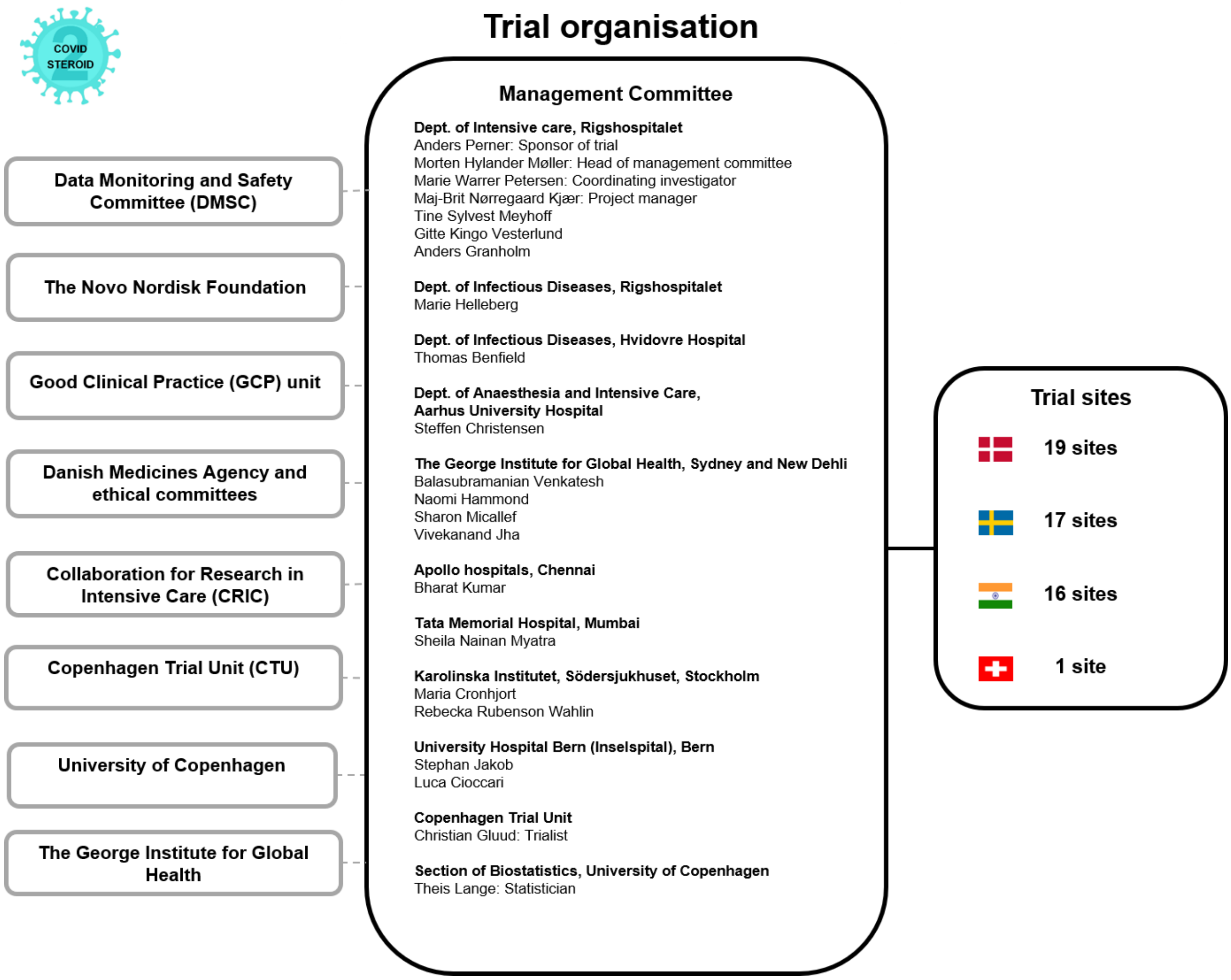

#### 18.2 *Appendix 2: Trial flow chart*

Please refer to the CONSORT Statement for more information (http://www.consort-statement.org/) (60). The flowchart will be modified to reflect the flow of participants in the trial. The flowchart (n= ) will be completed at the end of the trial.

**CONSORT 2010 Flow Diagram**

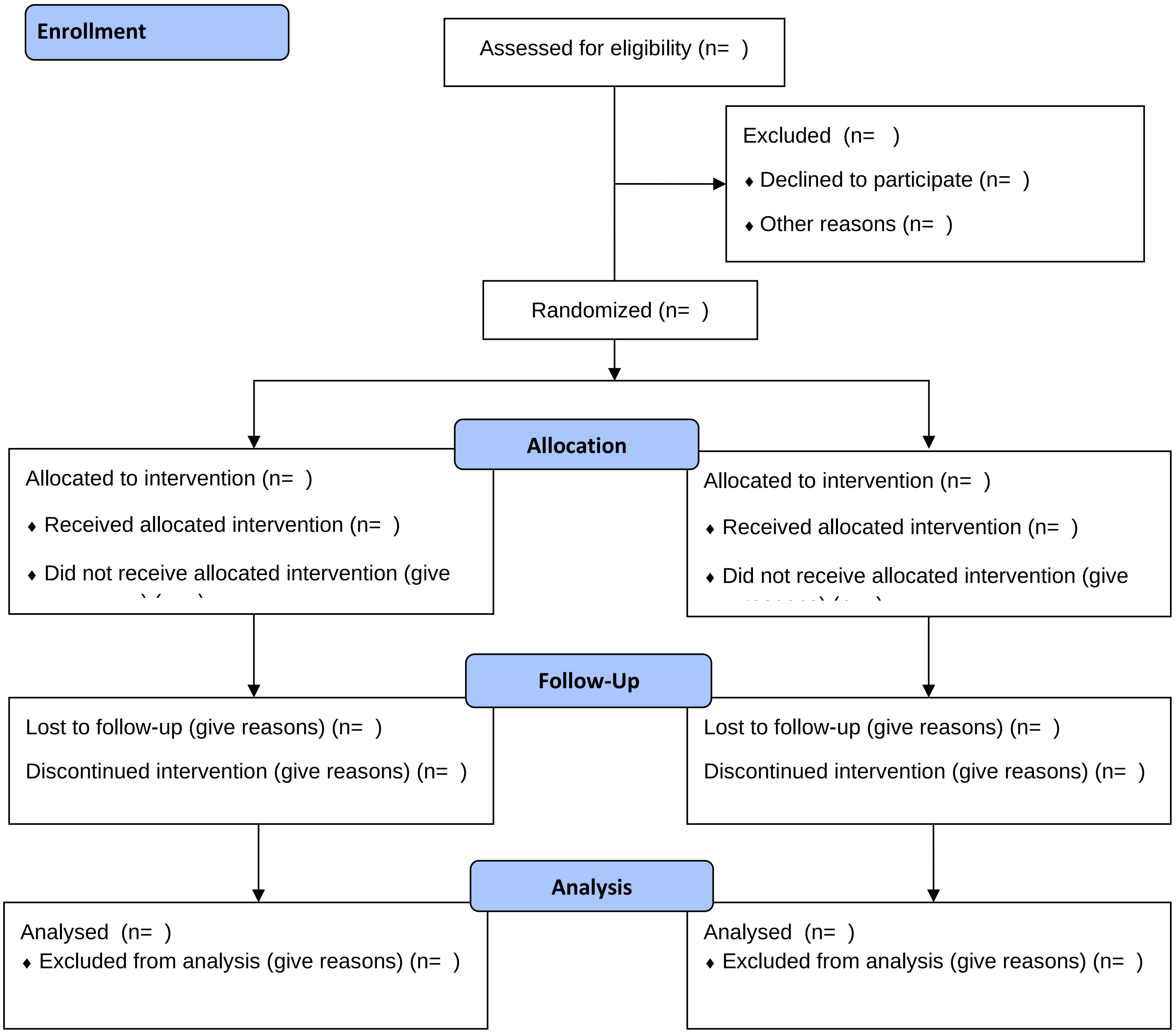

#### 18.3 Appendix 3: Trial definitions

**Definition of stratification variables**

Site: all participating trial sites (hospitals) will be assigned a number identifying the site.

Invasive mechanical ventilation: use of mechanical ventilation via a cuffed endotracheal tube at the time of randomisation.

Age: the age of the participant in whole years at the time of randomisation. Is the participant above 70 years old? (y/n). The participants will be stratified according to age ≥70 years versus <70 years.

**Participant identification**

National identification number (NIN): civil registration number (CPR number, 10 digits without dash) in or replacement CPR number if the participant does not have a CPR number in Denmark. Fictive NIN in other countries than Denmark generated from date of birth or year of birth and trial site ID.

**Definition of the inclusion criteria**

Age: defined under *Definition of stratification variables*

Confirmed SARS-CoV-2 requiring hospitalisation**:** We will include patients admitted to a trial site with SARS-CoV-2. We will accept any detections of SARS-CoV-2 approved by the national Health Authorities in the participating countries. Currently, detection of SARS-CoV-2 RNA from upper (i.e. pharyngeal swap) or lower airway secretions (i.e. tracheal secretion or bronchoalveolar lavage) is used.

Supplementary oxygen criterion at the time of randomisation:

- Invasive mechanical ventilation: Defined under *Definition of stratification variables* **OR**
- Non-invasive ventilation or continuous use of continuous positive airway pressure (CPAP) for hypoxia: Non-invasive ventilation includes positive pressure ventilation via a tight mask or helmet, continuous use of CPAP (mask, helmet or tracheostomy). This does not include intermittent use of CPAP.
- Oxygen supplementation with an oxygen flow ≥10 L/min irrespectively of system used (mask or nasal cannula) or the addition of atmospheric air

**Definition of the exclusion criteria**

Use of systemic corticosteroids in doses higher than 6 mg dexamethasone equivalents for other indications than COVID-19: systemic corticosteroids (IV, IM, oral or per GI tube; not including nebulised, inhaled or transdermal corticosteroids) in doses higher than 6 mg dexamethasone / 6 mg betamethasone / 200 mg cortisone / 160 mg hydrocortisone / 32 mg methylprednisolone / 40 mg prednisolone / 40 mg prednisone. Other indications include:

- Adrenal insufficiency (i.e. primary, secondary or tertiary)
- Anti-emetic treatment (i.e. post-operative or chemotherapy-induced nausea and vomiting)
- Immunosuppressive treatment (i.e. rheumatic diseases, allergic diseases, chronic obstructive pulmonary disease, haematological diseases, chronic kidney diseases, autoimmune hepatitis, inflammatory bowel disease, chronic neurological diseases)

Use of systemic corticosteroids for COVID-19 for 5 days or more:  Use of systemic corticosteroids for COVID-19 for 5 consecutive days or more up to the day of screening.

Invasive fungal infection: Any of the following:

- Suspected invasive fungal infection: presence of plasma markers in blood (e.g. candida mannan antigen and galactomannan antigen)
- Confirmed invasive fungal infection: positive culture from blood, peritoneal fluid or tissue

Active tuberculosis: Either microbiologically confirmed or diagnosed based on epidemiological, clinical and radiographic data.

Pregnancy: confirmed by positive urine human gonadotropin (hCG) or plasma-hCG.

Known hypersensitivity to dexamethasone: history of any hypersensitivity reaction to dexamethasone, including but not limited to urticaria, eczema, angioedema, bronchospasm and anaphylaxis.

Consent not obtainable: patients where the clinician or investigator is unable to obtain the necessary consent according to the national regulations, including patients with no relatives or patients who are hospitalised against their will.

**Definition of baseline variables**

Sex: the genotypic sex of the participant

Age at enrolment: the age of the participant in whole years at the time of randomisation. The age will be calculated from the date of birth and date of enrolment in the COVID STEROID 2 trial.

Date of admission to hospital: the date of admission to the first hospital the participant was admitted to during the current hospital admission

Department at which participant was included:

- Emergency department: accident/emergency/casualty/acute department at COVID STEROID 2 trial site
- Hospital ward: medical or surgical ward at COVID STEROID 2trial site, including dedicated COVID-19 hospital wards
- Intermediate care unit: area of the hospital with higher resources to monitor patients as defined by the site, but invasive mechanical cannot be given.
- Intensive care unit: area of the hospital where invasive mechanical can be given.
- Other: any location in the same or another hospital not covered in the other categories

Use of respiratory support at randomisation:

- Closed system (y/n): Use of invasive mechanical ventilation as defined under *Definition of stratification variables*or use of Non-invasive ventilation or continuous use of continuous positive airway pressure (CPAP) for hypoxia as defined under *Definition of inclusion criteria.* If yes, latest $FiO_2$ prior to randomisation
- Open system with an oxygen flow ≥10 L/min: If yes, the maximum supplemental oxygen flow on an open system at randomisation (+/- 1 h) will be registered.

Limitations of care (y/n): participant with limitation(s) in use of life support (i.e. invasive mechanical ventilation, circulatory support, renal replacement therapy) and/or cardio-pulmonary resuscitation at the time of randomisation.

Chronic use of systemic corticosteroids for other indications than COVID-19 (y/n): Systemic corticosteroids (IV, IM, oral or per GI tube; not including nebulised, inhaled or transdermal corticosteroids) for any other indications than COVID-19 at the time of randomisation.

Treatment during current hospital admission prior to randomisation:

Agents with potential anti-viral action used against COVID-19: any treatment that potentially inhibits viral replication, categorised as remdesivir, convalescent plasma, or other (e.g. umifenovir, interferon alfa, interferon beta, camostat).

Anti-bacterial agents: any antibiotic treatment commenced due to documented or suspected bacterial infection before microbiological results are available

Agents with potential anti-inflammatory action: any treatment with potential anti-inflammatory actions used against COVID-19 prior to screening, categorised as Janus kinase inhibitor, IL-6 inhibitors or other.

Co-morbidities: any chronic co-morbidity present in the past medical history prior to admission and defined as follows:

- History of ischemic heart disease or heart failure: previous myocardial infarction, invasive intervention for coronary artery disease, stable or unstable angina, NYHA class 3 or 4 or any measured LVEF <40%.
- Diabetes mellitus: Treatment at time of hospital admission with any anti-diabetic medications.
- Chronic pulmonary disease: Treatment at time of hospital admission with any relevant drug indicating chronic pulmonary disease.
- Immunosuppressive therapy within the last 3-months: use of systemic immunosuppressive drugs (e.g. tumor necrosis factor (TNF) inhibitors, calcineurin inhibitors, mTOR inhibitors, anti-thymocyte globulins, interleukin-2 inhibitors, mycophenolate, azathioprine, belimumab, corticosteroids), chemotherapy (e.g. alkylating agents, anti-metabolites, mitotic inhibitors, topoisomerase inhibitors, others) or radiotherapy within the last 3 months before randomisation.

Blood values, interventions and vital parameters:

- Participant weight: measured or estimated in kg
- $PaO_2$, $SaO_2$ and lactate prior to inclusion: will be assessed from the most recent arterial blood gas sample; alternatively, if arterial blood gas sample is not available, $SpO_2$ will be assessed from the most recent measure by pulse oximeter.
- Circulatory support: infusion of any vasopressor/inotrope agent for a minimum of 1 hour (i.e. norepinephrine, epinephrine, phenylephrine, vasopressin analogues, angiotensin, dopamine, dobutamine, milrinone or levosimendan) within the last 24 hours prior to randomisation.
- Renal replacement therapy: any form of acute or chronic intermittent or continuous renal replacement therapy (including days between intermittent dialysis) within the last 72 hours prior to randomisation.

**Definition of variables assessed in day forms (day 1-14)**

- Invasive mechanical ventilation (on this day): defined under *Definition of the inclusion criteria.*

- Circulatory support (for at least 1 hour on this day): defined under *Definition of the baseline variables.*
- Any form of renal replacement therapy (on this day): any form of renal replacement therapy (e.g. dialysis, hemofiltration or hemodiafiltration) at any rate on this day. Including days between intermittent renal replacement therapy.

- SAR on this day (y/n for everyone)

- New episodes of septic shock: we will define septic shock according to the Sepsis-3 criteria (61):
    - Suspected or confirmed superinfection
    - New infusion (or 50% increase) of vasopressor/inotrope agent (*Definition in the baseline variables*) to maintain a mean arterial blood pressure of 65 mmHg or above
    - Lactate of 2 mmol/L or above in any plasma sample performed on the same day
- Invasive fungal infection: defined under *Definition of exclusion criteria*
- Clinically important gastrointestinal (GI) bleeding: any GI bleeding **AND** use of at least 2 unit of red blood cells on the same day. GI bleed defined as hematemesis, coffee ground emesis, melena, haematochezia or bloody nasogastric aspirate on this day.
- Anaphylactic reaction to IV dexamethasone: anaphylactic reactions defined as urticarial skin reaction **AND** at least one of the following observed after randomisation
    - Worsened circulation (>20% decrease in blood pressure or >20% increase in vasopressor dose)
    - Increased airway resistance (>20% increase in the peak pressure on the ventilation)
    - Clinical stridor or bronchospasm
    - Subsequent treatment with bronchodilators

**Definition of variables assessed in day forms (from day 1 and up to 10 days)**
- Use of open-label systemic corticosteroids on this day: Use of any open-label systemic (IV, IM or oral/per GI tube) corticosteroids (i.e. hydrocortisone, methylprednisolone, dexamethasone, prednisolone or prednisone) in any dose

- Trial intervention: Did the participant receive trial medication on this day: yes, if the trial participant received the bolus of trial medication on this day; no, if the trial participant did not receive the bolus of trial medication on this day.

  - If no, please apply reason for violating the protocol: By error/lack of resources, other reason.

**Definitions of outcome measures**

Primary outcome

Days alive without life support (i.e. invasive mechanical ventilation, circulatory support or renal replacement therapy) at day 28: will be assessed from the use of life support including invasive mechanical ventilation, vasopressor/inotrope, and renal replacement therapy as defined in *Definition of inclusion criteria*, *Definition of baseline variables* and *Definition of variables assessed in day form.* Total number of days alive without all 3 life supporting interventions within 28 days after randomisation.

Secondary outcomes

- Number of participants with one or more serious adverse reactions (SARs) at day 28: at least one new episode of either septic shock, invasive fungal infection, clinically important GI bleeding or anaphylactic reaction to IV dexamethasone as defined under *Definition of variables assessed in day form.*
- All-cause mortality at day 28 after randomisation: death from any cause within 28 days post-randomisation.
- All-cause mortality at day 90 after randomisation: death from any cause within 90 days post-randomisation.
- Days alive without life support (i.e. invasive mechanical ventilation, circulatory support or renal replacement therapy) at day 90: will be assessed from the use of life support invasive mechanical ventilation including vasopressor/inotrope, and renal replacement therapy as defined in *Definition of inclusion criteria*, *Definition of baseline variables* and *Definition of variables assessed in day form.* Total number of days alive without all 3 life supporting interventions within 28 days after randomisation.
- Days alive and out of hospital at day 90: will be assessed from the discharge date from the index hospitalisation, the number of days readmitted to hospital (if any) and date of death, if relevant, within the 90-day period

- All-cause mortality at 180 days after randomisation: death from any cause within 180 days post-randomisation.
- Health-Related Quality of Life (HRQoL) at 180 days after randomisation: HRQoL at 180 days (+/- 2 weeks): EQ-5D-5L and EQ-VAS scores (https://euroqol.org/) obtained by survey by mail or phone as chosen by the participant. Non-survivors will be given the worst possible score. If the participant is incapable of answering the questionnaire (e.g. due to cognitive impairment or coma) we will ask proxies to assess HRQoL for the trial participant (proxy point of view) using the questionnaire aimed for proxies. Non-survivors will be given the worst possible score. EQ-5D-5L will be converted to an index value in combination with the EQ-VAS quantitative measure (0-100 points) quantifying self-rated health

**Definitions of other variables assessed during follow up**

- Discharged against medical advice to home/other hospital/other facility (y/n)
    - If yes: was the participant on any life support (i.e. invasive mechanical ventilation, circulatory support or renal replacement therapy) at the time of discharge (y/n)?
        - If no, did the participant receive supplementary oxygen at the time of discharge?
            - Yes, 0-9 L/min of supplementary oxygen
            - Yes, ≥10 L/min of supplementary oxygen
            - No

- Use of ECMO from randomisation to day 28: oxygen supplied through extracorporeal membrane on any day from randomisation to day 28.

**Definitions of subgroups**

Elderly patients: ≥70 years versus <70 years. Age is defined under *Definition of stratification variables.*

Invasive mechanical ventilation: invasive mechanical ventilation versus oxygen by other delivery systems. Invasive mechanical ventilation is defined under *Definition of stratification variables*; oxygen by other delivery system encompass both non-invasive ventilation, continuous use of CPAP and oxygen supplementation with an oxygen flow ≥10 L/min irrespectively of system used or the addition of atmospheric air as defined under *Definition of inclusion criteria.*

Shock: patients with shock versus without shock. Shock of any cause in patients requiring infusion of vasopressor/inotropic agent (norepinephrine, epinephrine, phenylephrine, vasopressin analogues, angiotensin, dopamine, dobutamine, milrinone or levosemindan) to maintain a mean arterial blood pressure of 65 mmHg or above AND with a lactate of 2 mmol/L or above in any plasma within 24 hours of randomisation.

Duration of corticosteroid use before enrolment in COVID STEROID 2 trial: patients who received any systemic corticosteroid for COVID-19 for 0 to 2 consecutive days compared to 3 to 4 consecutive days up to enrolment.

Limitations of care: patients with limitations of care compared to patients without limitations of care. Limitation of care is defined under *Definition of baseline variables*.

Chronic use of systemic corticosteroids for other indications than COVID-19: patients with versus without chronic use of systemic corticosteroids as defined under *Definition of baseline variables*.

#### 18.4 Appendix 4: Trial medication labels for dexamethasone and betamethasone

**COVID STEROID trial medication for clinical trial**
Dexamethasone 12 mg OR dexamethasone 6 mg

For injection

Patient name..........................................................................................................................

Identification number.............................................................................................................

Date and time of preparation of trial medication....................................................

Signature................................................................................................................................

Must be stored at ≤ 25 °C

Sponsor: Prof. Anders Perner, Dept. of Intensive Care, Rigshospitalet, Denmark. Tel. +45 3545 8333

**COVID STEROID trial medication for clinical trial**
Betamethasone 12 mg OR betamethasone 6 mg

For injection

Patient name..........................................................................................................................

Identification number.............................................................................................................

Date and time of preparation of trial medication....................................................

Signature................................................................................................................................

Must be stored at ≤ 25 °C

Sponsor: Prof. Anders Perner, Dept. of Intensive Care, Rigshospitalet, Denmark. Tel. +45 3545 8333

#### 18.5 Appendix 5: Charter for the independent data monitoring and safety committee

**Introduction**

The independent Data Monitoring and Safety Committee (IDMSC) will constitute its own plan of monitoring and meetings. However, this charter defines the minimum of obligations and primary responsibilities of the IDMSC, its relationship with other trial components, its membership, and the purpose and timing of its meetings, as perceived by the COVID STEROID 2 Management Committee. The charter also outlines the procedures for ensuring confidentiality and proper communication, the statistical monitoring guidelines to be implemented by the IDMSC, and an outline of the content of the open and closed reports which will be provided to the IDMSC.

**Primary responsibilities of the IDMSC**

The IDMSC are responsible for safeguarding the interests of trial participants, assessing the safety and efficacy of the interventions during the trial, and for monitoring the overall conduct of the trial. The IDMSC will provide recommendations about stopping or continuing the trial to the Management Committee of the COVID STEROID 2 trial. The IDMSC may also – if applicable - formulate recommendations related to the selection/recruitment/retention of participants, their management, adherence to protocol-specified regimens and retention of participants, and the procedures for data management and quality control.

The IDMSC will be advisory to the COVID STEROID 2 Management Committee. The Management Committee will be responsible for promptly reviewing the IDMSC recommendations, to decide whether to continue or terminate the trial, and to determine whether amendments to the protocol or changes in trial conduct are required.

The IDMSC may meet physically or by phone at their own discretion in order to evaluate the planned interim analyses of the COVID STEROID 2 trial. The interim analysis will be performed by an independent statistician selected by the members of the IDMSC, Susanne Rosthøj from the Department of Biostatistics, University of Copenhagen. The IDMSC may additionally meet whenever they decide or contact each other by telephone or e-mail to discuss the safety for trial participants. The IDMSC can, at any time during the trial, request information about the distribution of events, including outcome measures and serious adverse reactions (SARs) according to group allocation. Further, the IDMSC can request unmasking of the interventions, if deemed important (see section on ‘closed sessions’). The recommendations of the IDMSC regarding stopping, continuing or changing the design of the trial should be communicated without delay to the COVID STEROID 2 Management Committee. As fast as possible, and no later than 48 hours, the Management Committee has the responsibility to inform all trial sites and investigators, about the recommendation of the IDMSC and the Management Committee decision hereof.

**Members of the IDMSC**

The IDMSC is an independent multidisciplinary group consisting of a clinician, a trialist and a biostatistician that, collectively, has experience in the conduct, monitoring and analysis of randomised clinical trials.

<u>IDMSC Clinician</u>

Christian Hassager, Professor in cardiology, Copenhagen University Hospital, Denmark

<u>IDMSC Trialist</u>

Manu Shankar-Hari, Clinician Scientist, Reader and Consultant in Intensive Care Medicine, National Institute for Health Research and Kings College, London, United Kingdom

<u>IDMSC Biostatistician</u>

Susanne Rosthøj, Department of Biostatistics, University of Copenhagen

**Conflicts of interest**

The members of the IDMSC will fill-in and sign a conflicts of interest form. IDMSC membership is restricted to individuals free of conflict of interest. The source of these conflicts may be financial, scientific, or regulatory in nature. Thus, neither trial investigators nor individuals employed by the sponsor, or individuals who might have regulatory responsibilities for the trial products, are members of the IDMSC. Furthermore. the IDMSC members do not own stocks in the companies having products being evaluated by the COVID STEROID 2 trial.

The IDMSC members will disclose to fellow members any consulting agreements or financial interests they have with the sponsor of the trial, with the contract research organisation (CRO) for the trial (if any), or with other sponsors having products that are being evaluated or having products that are competitive with those being evaluated in the trial. The IDMSC will be responsible for deciding whether these consulting agreements or financial interests materially impact their objectivity.

The IDMSC members will be responsible for advising fellow members of any changes in these consulting agreements and financial interests that occur during the trial. Any IDMSC members who develop significant conflicts of interest during the trial should resign from the IDMSC.

IDMSC membership is to be for the duration of the clinical trial. If any members leave the IDMSC during the trial, the Management Committee will appoint the replacement(s).

**Formal interim analysis meetings**

One formal interim analysis meeting will be held to review data related to protocol adherence, treatment efficacy and participant safety. The 3 members of the IDMSC will meet when 28-day follow-up data of 500 participants (50% of sample size) have been obtained.

**Final analysis meeting**

The 3 members of the IDMSC will meet when 28-day follow-up data the full sample size (1,000 participants) have been obtained.

**Proper communication**

To enhance the integrity and credibility of the trial, procedures will be implemented to ensure that the IDMSC has sole access to evolving information from the clinical trial regarding comparative results of efficacy and safety data aggregated by treatment group. An exception will be made to permit access to an independent statistician who will be responsible for serving as a liaison between the database and the IDMSC.

At the same time, procedures will be implemented to ensure that proper communication is achieved between the IDMSC and the Management Committee. To provide a forum for exchange of information among various parties who share responsibility for the successful conduct of the trial, a format for open sessions and closed sessions will be implemented. The intent of this format is to enable the IDMSC to preserve confidentiality of the comparative efficacy results while at the same time providing opportunities for interaction between the IDMSC and others who have valuable insights into trial-related issues.

**Closed sessions**

Sessions involving only IDMSC membership who generates the closed reports (called closed sessions) will be held to allow discussion of confidential data from the clinical trial, including information about protocol adherence and the relative efficacy and safety of interventions. To ensure that the IDMSC will be fully informed in its primary mission of safeguarding the interest of participants, the IDMSC will be blinded in its assessment of safety and efficacy data. However, the IDMSC can request unblinding from the Management Committee.

Closed reports will include analysis of the primary outcome measure and rates of SARs. These closed reports will be prepared by the independent IDMSC biostatistician, with assistance from the trial data manager, in a manner that allow them to remain blinded. The closed reports should provide information that is accurate, with follow-up on the primary outcome that is complete as soon as possible and at latest within one month from the date of the IDMSC meeting.

**Open reports**

For each IDMSC meeting, open reports will be available to all who attend the IDMSC meeting. The reports will include data on recruitment and baseline characteristics, and pooled data on eligibility violations, completeness of follow-up, and compliance. The independent IDMSC statistician will prepare these open reports in co-operation with the trial data manager. The reports should be provided to IDMSC members approximately three days prior to the date of the meeting.

**Minutes of the IDMSC Meetings**

The IDMSC will prepare minutes of their meetings. The closed minutes will describe the proceedings from all sessions of the IDMSC meeting, including the listing of recommendations by the committee. Because it is possible that these minutes may contain unblinded information, it is important that they are not made available to anyone outside the IDMSC.

**Recommendations to the Management Committee**

The planned interim analyses will be conducted after participant no. 500 have been followed for 28 days.

After the interim analysis meetings, the IDMSC will make a recommendation to the Management Committee to continue, hold or terminate the trial.

The independent IDMSC will recommend pausing or stopping the trial if group-differences in the primary outcome measure, SARs or suspected unexpected serious adverse reactions (SUSARs) are observed at the interim analysis with statistical significance levels adjusted according to the O'Brien-Fleming alfa-spending function (57). If the recommendation is to stop the trial, the IDMSC will discuss and recommend on whether the final decision to stop the trial will be made after the analysis of all participants included at the time (including participants randomised after this interim analysis) or whether a moratorium shall take place (setting the trial at hold) in the further inclusion of participants during these extra analyses. If further analyses of the participants included after the interim analysis is recommended, the rules for finally recommending stopping of the trial should obey the O'Brien-Fleming stopping boundary (57). Furthermore, the IDMSC can recommend pausing or stopping the trial if continued conduct of the trial clearly compromises participant safety. However, stopping for futility will not be an option as an intervention effects less than those estimated in the power calculation for the primary outcome may be clinically relevant as well.

All recommendation will be based on safety and efficacy considerations and will be guided by statistical monitoring guidelines defined in this charter and the trial protocol.

The Management Committee is jointly responsible with the IDMSC for safeguarding the interests of participants and for the conduct of the trial. Recommendations to amend the protocol or change the conduct of the trial made by the IDMSC will be considered and accepted or rejected by the

Management Committee. The Management Committee will be responsible for deciding whether to continue, hold or stop the trial based on the IDMSC recommendations.

The IDMSC will be notified of all changes to the trial protocol or conduct. The IDMSC concurrence will be sought on all substantive recommendations or changes to the protocol or trial conduct prior to their implementation.

After completion of the interim analysis, the recommendations from the IDMSC and the conclusion reached by the Management Committee will be submitted to the Ethics Committee.

After completion of the full analysis of outcomes at day 28 (i.e. days alive without life support, mortality and SAR), the IDMSC will make a recommendation to the Management Committee to submit a primary report on 28-day outcomes or await the 90-day outcomes.

**Statistical monitoring guidelines**

The outcome parameters are defined in the statistical analysis plan in the COVID STEROID trial protocol. For the two intervention groups, the IDMSC will evaluate data on:

- Days alive without life support at day 28

- Mortality at day 28

- The number of participants with ≥1 SAR(s) and/or SUSAR(s) at day 28

The IDMSC will be provided a masked data set (as group 0 and 1) from the coordinating centre. The data set will include data on stratification variables and outcome measures according to the outcomes above in the two groups.

Based on evaluations of these outcomes, the IDMSC will decide if they want further data from the coordinating center and when to perform the next analysis of the data. For analyses, the data will be provided in one file as described below.

The IDMSC may also be asked to ensure that procedures are properly implemented to adjust trial sample size or duration of follow-up to restore power, if protocol specified event rates are inaccurate. If so, the algorithm for doing this should be clearly specified.

**Conditions for transfer of data from the Coordinating Centre to the IDMSC**

The IDMSC will be provided with a data file containing the data defined as follows:

Row 1 contains the names of the variables (to be defined below).

Row 2 to N (where N-1 is the number of participants having entered the trial) each contains the data of one participant.

Column 1 to p (where p is the number of variables to be defined below) each contains in row 1 the name of a variable and in the next N rows the values of this variable.

The values of the following variables should be included in the database for the all three interim analyses:

1. screening_id: a number that uniquely identifies the participant

2. rand_code: The randomisation code (group 0 or 1). The IDMSC is not to be informed on what intervention the groups received

3. days_alive_without_lifesup_d28_cum_indic (continuous scale)

4. day_28_indic: 28 day-mortality indicator (2 = censored, 1=dead, 0=alive at day 28)

5. SAR_indic: SAR indicator (1 = one or more SARs, 0 = no SAR)

#### 18.6 Appendix 6: Informed consent

Participants will be enrolled after consent by proxy is obtained according to Danish regulations. We will follow the normal procedures for collection of informed consent and any modifications to these as approved by the Ethics Committee due to restrictions in physical visits to the hospitals during the current SARS-CoV-2 epidemic. All consenting parties will be provided with written and oral information about the trial, so he/she is able to make an informed decision about participation in the trial.

All patients with COVID-19 and severe hypoxia will be temporarily incompetent because of the acute illness, low oxygen saturation and stress-response associated with lack of oxygen. Thus, participants will be enrolled after obtaining informed consent from a doctor (first trial guardian), who is independent of the trial, who has knowledge of the clinical condition and who is familiar with the trial protocol to such extent that he/she can judge for each patient, if it will be reasonable to enroll the patient in the trial.

As soon as possible after enrolment, consent will be obtained from the participant’s next of kin and a second trial guardian.

The second trial guardian is also a doctor who is independent of the trial, who has knowledge of the clinical condition and who is familiar with the trial protocol to such extent that he/she can judge for each patient, if it will be reasonable to enroll the patient in the trial.

To minimise the risk of transmission of SARS-CoV-2 between trial staff and the next of kin, we will inform and obtain informed consent from the next of kin by telephone. We will contact the next of kin by telephone and arrange a time and date for a telephone conversation with a member of the trial staff (e.g. doctor, research nurse, medical student etc) who is certified in obtaining informed consent. During this conversation, we will arrange how to send the written information to the next of kin (i.e. e-mail, post). We will encourage the next of kin to read the written information before the next conversation. We will also encourage the next of kin to bring a companion; in this case, the telephone conversation will be held with the telephone on speaker. After we have informed the next of kin about the trial, we will ask the next of kin to return the signed consent form by post.

Participants will be asked for informed consent as soon as possible after they regain consciousness. For participants, both oral and written information will be given preferably in person. The participant has the right to bring a companion.

If deemed necessary by the treating doctor, we will inform the participant orally before enrolment. In these instances, we will not include the patient, if he/she declines to participate. If the patient accepts to participate, we will re-inform the participant once he/she has regained full competence, i.e. when the participant receives less than 10 L/min of supplementary oxygen; is not mechanically ventilated; and is awake, alert and oriented as judged by treating clinician. First hereafter, we will collect the informed consent. For these participants, the procedure for obtaining informed consent will follow the same rules as stated above.

All consent forms will be signed by the consenting party and the member of trial staff who have provided trial information for the consenting party. We will emphasise that the consenting party has at least 24 hours to decide whether to give consent or not. Written information and the consent forms will be subjected to review and approval by the relevant ethic committees.

**Lack of informed consent from the participant's next of kin**

If information about the participant's next of kin is not available after inclusion, the investigator will seek information from e.g. the participant's general practitioner, the police, nursing homes etc. In these situations, it may take 1-2 weeks to conclude that no next of kin can be identified. If a next of kin is not identified and the participant remains incompetent, the trial intervention will be discontinued. All initiatives to identify the participant's next of kin will be documented in patient files, logs or similar.

**Lack of informed consent from the participant's next of kin and the participant deceases**

If the participant deceases before informed consent has been obtained (due to rapid progression of critical illness or because the participant's next of kin is not yet identified) and the participant has been correctly included in the trial, the collected data will be kept for analysis.

**Deviation from the standard informed consent**

According to the standard informed consent form from the National Ethics Committee regarding competent participants, the participant can choose not to receive information about the data collected during the trial. However, the purpose of this trial is not to generate new knowledge about the specific participant, so we find that this question is redundant, and have omitted the question from the consent form to spare the participant from making unnecessary decisions.

**Trial personnel**

Screening will be performed by the clinical staff. Collection of informed consent will be performed by the dedicated trial staff. If questions arise during informed consent, responsible trial staff can be reached through a 24-h hotline. All personnel with functions in the COVID STEROID 2 trial will be trained and approved according to GCP-guidelines before engaging in the trial.

#### 18.7 Appendix 7: Co-enrolment

Based upon an updated critical appraisal of the literature, the COVID STEROID 2 Management Committee endorses and encourages co-enrolment in the COVID STEROID 2 trial. The following issues have been considered.

**Ethical considerations**

Preventing eligible patients from co-enrolment in trials, which they would authentically value participating in, and whose material risks and benefits they understand, violates their autonomy - and thus contravenes a fundamental principle of research ethics (62).

Permitting co-enrolment is in accordance with existing recommendations for the conduct of trustworthy clinical practice guidelines, taking into account benefits and harms, quality of evidence, values and preferences (of patients or their relatives) and cost considerations, as outlined by the Institute of Medicine, the Guideline International Network, and according to The Grading of Recommendations Assessment, Development and Evaluation (GRADE) methodology (63-65).

Patient relatives have limited concerns about co-enrolment (66).

**General considerations**

Critically ill patients receive many different interventions in addition to the trial intervention because of acute and chronic illness. Consequently, the potential for interactions is a prerequisite in clinical trials in critically ill patients, and co-enrolment is thus little different from what occurs in single-enrolment trials (62).

In pragmatic trials, like the COVID STEROID 2 trial, other interventions will be given at random and are therefore difficult to control for. If interaction in fact is an issue, it may be better controlled for if patients are co-enrolled and randomised to more than one intervention.

Clinical research with a potential to inform and improve clinical practice is valuable and should be supported. More high-quality clinical research can be conducted in a timely fashion and more information can be generated to guide clinical practice, if co-enrolment is permitted (67).

**Scientific and statistical considerations**

Pragmatic clinical trials allowing inclusion of a broad range of trial participants and options for drug treatments and other therapies (co-enrolment) have higher external validity/generalizability than non-pragmatic trials with restrictions regarding trial participants and co-enrolment (68).

Non-pragmatic trials with restrictions regarding study participants and co-enrolment are exposed to drugs and other treatments in a less clinically relevant setting where interactions are largely uncontrolled and poorly evaluated. Co-enrolment in pragmatic trials facilitates evaluation of clinically relevant and patient-important interactions (62).

Co-enrolment into two or more trials does not invalidate the original randomization of the individual trials. Separate analysis of each individual trial, ignoring the issue of co-enrolment into the other trial, will retain the balance of patient characteristics expected by standard random assignment within each trial (62).

The National Institute of Health supports co-enrolment (68); so does the Canadian Critical Care Trials group (http://www.ccctg.ca/Home.aspx) and the Australian New Zealand Intensive Care Society's Clinical Trial Group (http://www.anzics.com.au/Pages/CTG/CTG-home.aspx). We have co-enrolment agreements with the two latter research groups.

Co-enrolment into two or more trials does not seem to affect the natural course of the disease of the other condition being studied (62). Co-enrolment does not appear to influence patient safety or trial results (69, 70). Empirically, co-enrolment has a small effect on study power (62).

In conclusion, we highly support and encourage co-enrolment because of overall benefit, including ethical, practical and scientific benefit, and no evidence of harm.

**Co-enrolment agreement form**

We will encourage engagement in research projects other than the COVID STEROID 2 trial. Please fill in the information of the trial to be evaluated as counterpart for co-enrolment with the COVID STEROID 2 trial and send it by e-mail to.

Once we have received the information below, we will contact the principal/coordinating investigator of the trial and facilitate exchange of protocols and other relevant documents between the Management Committees. You will find a list of titles already considered for co-enrolment by clicking http://www.cric.nu/covid-steroid-2-co-enrolment-list/

We have prepared the form for only one trial, but please feel free to copy as many forms as you need. The co-enrolment agreement form can be found by clicking http://www.cric.nu/covid-steroid-2-co-enrolment-form/

**Official full/short title of the project:**

**Contact information of principal/coordinating investigator of the trial:**

Name:

E-mail:

#### 18.8 Appendix 8: List of proposed sub-studies

A Bayesian secondary analysis of all outcomes recorded within 90 days of randomisation.

#### *18.9 Local trade names for dexamethasone/betamethasone used in the COVID STEROID 2 trial*

**Denmark**

Dexavit™, Vital Pharma Nordic, Denmark, ATC code: H02AB02.

**Sweden**

Betapred™, Alfasigma S.p.A., Italy, ATC code: H02AB01.

**Switzerland**

Mephameson™, Mepha Pharma AG, Switzerland, ATC code: H02AB02.

**India**

Daksone™, Daksh Pharmaceuticals Pvt. Ltd, India, ATC code: H02AB02.

Dacdac™, Wockhardt Limited (Merind), India, ATC code: H02AB02.

Decmax™, GLS Pharma Ltd., India, ATC code: H02AB02.

Demisone™, Cadila Pharmaceuticals Ltd. (Genvista), India, ATC code: H02AB02.

Dexona™, Zydus Cadila Healthcare Ltd. (Alidac), India, ATC code: H02AB02.

Dex-V™, Vensat Bio, India, ATC code: H02AB02.

Intradex™, Intra Labs India Pvt. Ltd, India, ATC code: H02AB02.

### Higher vs. Lower Doses of Dexamethasone in Patients with COVID-19 and Severe Hypoxia: the COVID STEROID 2 trial

**Management Committee**

Anders Perner, sponsor

Morten Hylander Møller

Marie Warrer Petersen

Maj-Brit Nørregaard Kjær

Tine Sylvest Meyhoff

Marie Helleberg

Anders Granholm

Gitte Kingo Vesterlund

Thomas Benfield

Steffen Christensen

Maria Cronhjort

Rebecka Rubenson Wahlin

Stephan Jakob

Luca Cioccari

Balasubramanian Venkatesh

Vivekanand Jha

Bharath Kumar Tirupakuzhi Vijayaraghavan

Sheila Nainan Myatra

Naomi Hammond

Oommen John

Abhinav Bassi

Sharon Micallef

Christian Gluud, trialist

Theis Lange, statistician

**Protocol version and date**

**1.9, 27.01.2021**

**Applicable protocol registration numbers**

ClinicalTrials.gov identifier NCT04509973

Ethics committee number H-20051056

EudraCT number 2020-003363-25

Danish Medicines Agency number 2020-07-16

1 ABSTRACT ... 6
2 ADMINISTRATIVE INFORMATION ... 9
2.1 Local and International Sponsors ... 9
2.2 Local investigators and clinical trial sites ... 11
2.3 Methodological trial sites ... 18
3 LIST OF ABBREVIATIONS ... 20
4 INTRODUCTION AND BACKGROUND ... 22
4.1 Severe acute respiratory syndrome coronavirus 2/Coronavirus Disease 19 ... 22
4.2 Corticosteroids in COVID-19 ... 22
4.3 Type and dose of corticosteroids for sepsis, ARDS and COVID-19 ... 23
4.4 Ethical justification and trial rationale ... 28
4.5 Trial conduct ... 29
5 TRIAL OBJECTIVES ... 30
6 TRIAL DESIGN ... 30
6.1 Randomisation ... 30
6.2 Blinding ... 30
6.3 Unblinding ... 32
6.4 Participant timeline ... 32
7 SELECTION OF PARTICIPANTS ... 33
7.1 Inclusion criteria ... 33
7.2 Exclusion criteria ... 33
7.3 Participant discontinuation and withdrawal ... 34
8 SELECTION OF TRIAL SITES AND PERSONNEL ... 35
8.1 Trial sites and setting ... 35
8.2 Trial personnel ... 35
8.3 Trial interventions ... 36
8.4 Experimental intervention ... 36
8.5 Control intervention ... 36
8.6 Co-interventions ... 36
8.7 Concomitant interventions ... 37
8.8 Monitoring of participants ... 37
8.9 Criteria for modification of interventions for a given trial participant ... 37
8.10 Assessment of participant compliance ... 38
8.11 Intervention accountability ... 38
9 OUTCOME MEASURES ... 39
9.1 Primary outcome ... 39
9.2 Secondary outcomes ... 39
10 SAFETY ... 40
10.1 Definitions ... 40
10.2 Risk and safety issues in the COVID STEROID 2 trial ... 41
10.3 Assessment of adverse events ... 41

### 19 Abstract

##### Background

Severe acute respiratory syndrome coronavirus-2 (SARS-CoV-2) is causing a pandemic of coronavirus disease 2019 (COVID-19) with many patients developing severe hypoxic respiratory failure. Many patients have died, and healthcare systems in several countries have been or will be overwhelmed because of a surge of patients needing hospitalisation and intensive care. The care in COVID-19 is primarily supportive, including respiratory and circulatory support.

Preliminary results from the Randomised Evaluation of COVid-19 thERapY (RECOVERY) trial have reported a reduction in 28-day mortality with low-dose dexamethasone (6 mg) once daily versus no intervention in hospitalised patients with COVID-19; an effect that may have been more pronounced in patients with increasing hypoxia. Yet, higher doses of dexamethasone may be beneficial in patients with non-COVID-19 acute respiratory distress syndrome. At present, it is unclear what dose of dexamethasone is most beneficial in patients with COVID-19 and severe hypoxia, and clinical equipoise exists.

##### Objectives

We aim to assess the effects of higher (12 mg) vs lower doses (6 mg) of intravenous dexamethasone on the number of days alive without life-support in adult patients with COVID-19 and severe hypoxia.

##### Design

International, parallel-group, centrally randomised, stratified, blinded, clinical trial.

##### Inclusion and exclusion criteria

We will screen all adult patients who have documented COVID-19 receiving at least 10 L/min of oxygen independent of delivery system OR mechanical ventilation. We will exclude patients who have an indication for systemic use of higher doses of corticosteroids (above 6 mg dexamethasone or equivalent) for other indications than COVID-19, who have received corticosteroids for COVID-

19 for 5 consecutive days or more, who have invasive fungal infection, who have active tuberculosis, who have known hypersensitivity to dexamethasone, who are pregnant, and those in whom informed consent cannot be obtained.

**Experimental intervention**

Dexamethasone 12 mg once daily for up to 10 days will be given as bolus injection.

**Control intervention**

Dexamethasone 6 mg once daily for up to 10 days will be given as bolus injection.

**Outcomes**

The primary outcome is days alive without life support (invasive mechanical ventilation, circulatory support, or renal replacement therapy) at day 28. Secondary outcomes are serious adverse reactions (new episode of septic shock, invasive fungal infection, clinically important gastrointestinal bleeding, or anaphylactic reaction to dexamethasone) at day 28; days alive without life support at day 90; days alive and out of hospital at day 90; all-cause mortality at day 28, day 90 and 180 days; and health-related quality of life at 180 days.

**Statistics**

The primary outcome will be compared using the Kryger Jensen and Lange test and reported as differences in means and medians along with 95% confidence intervals adjusted for the stratification variables (site, use of invasive mechanical ventilation, and age). Secondary binary outcomes will be analysed using binomial regression models with log links with results quantified as adjusted relative risks supplemented with adjusted risk differences, both with 95% confidence intervals.

**Trial size and testing strategy/design**

At maximum, we will randomise 1000 participants. The independent data monitoring and safety committee will conduct an interim analysis after 500 participants have been followed for 28 days. The alpha values for the interim analysis and the final analysis are 0.0054 and 0.0492, respectively as by the O'Brien-Fleming bounds, which preserves type I error at the usual 5%. In both analyses, the Kryger Jensen and Lange test will be employed to compare the groups on the primary outcome.

**Estimated timeline**

- August 2020, authority approvals and 1st patient randomised

- December 2020, interim analysis

- Mid 2021, last patient randomised and primary report on 28-day outcomes submitted.

- Late 2021, report on 90-day outcomes submitted

- Mid 2022, report on 180-day outcomes submitted

### 20 Administrative information

#### *20.1 Local and International Sponsors*

**International Sponsor/Central Coordinating Centre**

Dept. of Intensive Care

Rigshospitalet

Blegdamsvej 9
2100 Copenhagen Ø

+453545 7167

**International Sponsor Contact**

Anders Perner, senior staff specialist and professor in intensive care medicine

Dept. of Intensive Care

Rigshospitalet

Blegdamsvej 9

2100 Copenhagen Ø

+45 3545 8333

**Local sponsor /Coordinating Centre, Sweden**

Dept. of Clinical Science and Education

Södersjukhuset, Karolinska Institutet
Sjukhusbacken 10

11883 Stockholm

**Local Sponsor contact, Sweden**

Maria Cronhjort, senior staff specialist in intensive care medicine

Dept. of Anaesthesia and Intensive Care

Södersjukhuset, Karolinska Institutet
Sjukhusbacken 10

11883 Stockholm

**Local sponsor /Coordinating Centre, Switzerland**

Department of Intensive Care Medicine

Bern University Hospital (Inselspital)

Freiburgstrasse 15

3010 Bern

+41 31 632 53 00

**Local Sponsor contact, Switzerland**

Stephan Jakob, senior staff specialist and professor in intensive care medicine

Department of Intensive Care Medicine

Bern University Hospital (Inselspital)

Freiburgstrasse 15

3010 Bern

**Local sponsor /Coordinating Centre, India**

The George Institute for Global Health, New Delhi, India

Plot No. 57, Second Floor, Corporation Bank Building

Nagarjuna Circle, Punjagutta, Hyderabad - 500 082 | Telangana | India

T +91 40 3099 4444

F +91 40 3099 4400

**Local Sponsor contact, India**

Vivekanand Jha

The George Institute for Global Health, New Delhi, India

#### *20.2 Local investigators and clinical trial sites*

**Denmark**

Marie Warrer Petersen, Dept. of Intensive Care

Marie Helleberg, Dept. of Infectious Diseases

Vibeke Jørgensen, Dept. of Thoracic Anaesthesiology

Margit Smitt, Dept. of Neuroanaesthesiology

Rigshospitalet

Blegdamsvej 9

2100 Copenhagen Ø

Christian Aage Wamberg, Dept. of Anaesthesiology and Intensive Care

Bispebjerg Bakke 23

2400 Copenhagen NV

Klaus Tjelle, Dept. of Anaesthesia and Intensive Care

Thomas Benfield, Dept. of Infectious Diseases

Charlotte Suppli Ulrik, Dept. of Respiratory Medicine

Hvidovre Hospital, University of Copenhagen

2650 Hvidovre

Anne Sofie Andreasen, Dept. of Anaesthesia and Intensive Care

Herlev Hospital, University of Copenhagen

2730 Herlev

Thomas Mohr, Dept. of Intensive Care

Gentofte Hospital, University of Copenhagen

2900 Hellerup

Morten Bestle, Dept. of Anaesthesia and Intensive Care

North Zealand Hospital, University of Copenhagen
3400 Hillerød

Lone Poulsen, Dept. of Anaesthesia and Intensive Care

Zealand University Hospital, Køge

4600 Køge

Thomas Hildebrandt, Dept. of Anaesthesia and Intensive Care

Zealand University Hospital, Roskilde

4000 Roskilde

Klaus Vennick Marcussen, Dept. of Anaesthesia

Slagelse Hospital

4200 Slagelse

Christoffer G. Sølling, Dept. of Anaesthesia and Intensive Care

Viborg Hospital

8800 Viborg

Anne Brøchner, Dept. of Anaesthesia and Intensive Care

Kolding Hospital

6000 Kolding

Iben Strøm Darfelt, Dept. of Anaesthesia

Regional Hospital West Jutland, Herning

7400 Herning

Bodil S. Rasmussen, Dept. of Anaesthesia and Intensive Care

Aalborg University Hospital

9000 Aalborg

Steffen Christensen, Dept. of Anaesthesia and Intensive Care

Aarhus University Hospital

8200 Aarhus

Thomas Strøm, Dept. of Anaesthesia and Intensive Care

Odense University Hospital

5000 Odense

**Sweden**

Maria Cronhjort, Dept. of Anaesthesia and Intensive Care

Rebecka Rubenson Wahlin, Dept. of Anaesthesia and Intensive Care

Carl-Johan Treutiger, Dept. of Infectious diseases

Buster Mannheimer, Dept. Internal Medicine

Jacob Hollenberg, Dept of Cardiology

Södersjukhuset, 11883 Stockholm

118 83 Stockholm

Johan Mårtensson, Dept. of Anaesthesia and Intensive Care

Pontus Naucler, Dept. of Infectious diseases

Karolinska Universitetssjukhuset, Solna

171 76 Stockholm

Åke Norberg, Dept. of Anaesthesia and Intensive Care

Karolinska Universitetssjukhuset, Huddinge

141 86 Stockholm

Olof Wall, Dept. of Anaesthesia and Intensive Care

Sara Tehrani, Dept. Internal Medicine

Magnus Hedenstierna, Dept. of Infectious diseases

Danderyds Sjukhus

182 88 Stockholm

Andreas Wiklund, Dept. of Anaesthesia and Intensive Care

Capio St Görans Sjukhus

112 81 Stockholm

Michelle Chew, Dept. of Anaesthesia and Intensive Care

Universitetssjukhuset i Linköping

581 85 Linköping

Fredrik Schiöler, Dept. of Anaesthesia and Intensive Care

Vrinnevisjukhuset, Norrköping

603 79 Norrköping

Fredrik Sjövall, Dept. of Anaesthesia and Intensive Care

Anna Nilsson, Dept. of Infectious diseases

Skånes universitetssjukhus (SUS) Malmö

214 28 Malmö

Jonathan Oras, Dept. of Anaesthesia and Intensive Care

Magnus Gisslen, Dept. of Infectious diseases

Sahlgrenska universitetssjukhuset

413 45 Göteborg

**Switzerland**

Stephan Jakob, Department of Intensive Care Medicine

Luca Cioccari, Department of Intensive Care Medicine

Bern University Hospital (Inselspital)

3010 Bern

**India**

BK Tirupakuzhi Vijayaraghavan

Apollo Hospitals

600 006 Chennai

JV Divatia

Tata Memorial Hospital

400 012 Mumbai

Farhad N Kapadia

P.D. Hinduja National Hospital & Medical Research Centre

400 016 Mumbai

Pravin Amin

Bombay Hospital & Medical Research Centre

400 020 Mumbai

Sanjith Saseedharan

S L Raheja Fortis Hospital

400 016 Mumbai

Amit Shobhavat

K. J. Somaiya Super Specialty Hospital

400 022 Mumbai

Rajesh Chawla

Indraprastha Apollo Hospital

110 076 New Delhi

Omender Singh

Max Super Specialty Hospital, Saket

110017 New Delhi

Urvi Shukla

Symbiosis University Hospital and Research Centre

412 115 Pune

Binila Chacko

Christian Medical College Vellore
632 004 Vellore

Kedar Toraskar

Wockhardt hospitals

400011 Mumbai

Syed Moied Ahmed

Jawahar Lal Nehru Medical College, AMU

202002 Aligarh

Saif Mohd. Khan

Rajendra Institute of Medical Sciences

834009 Ranchi

Kapil Borawake

Vishwaraj Hospital

412201 Pune

Neeta Bose Nayak

Gotri General Hospital

390021 Vadodara

Zubair Umer Mohamed

Amrita Institute of Medical Sciences

682 041 Kochi

#### *20.3 Methodological trial sites*

**Coordinating centre**

Dept of Intensive Care

Rigshospitalet

Blegdamsvej 9
2100 Copenhagen Ø

+453545 7167

**Methods centre**

Copenhagen Trial Unit, Centre for Clinical Intervention Research
Rigshospitalet
Tagensvej 22

2200 Copenhagen N

**Statistical centre**

Section of Biostatistics
University of Copenhagen
Øster Farimagsgade 5
1014 Copenhagen K

**Independent Data Monitoring and Safety Committee (IDMSC)**

Christian Hassager, Professor in cardiology, Copenhagen University Hospital, Denmark

Manu Shankar-Hari, Clinician Scientist, Reader and Consultant in Intensive Care Medicine, National Institute for Health Research and Kings College, London, United Kingdom

Susanne Rosthøj, Statistician from University of Copenhagen

### 21 List of abbreviations

AE, Adverse Event

AR, Adverse Reaction

ARDS, Acute Respiratory Distress Syndrome

CI, Confidence Interval

CPAP, Continuous Positive Airway Pressure

CRIC, Collaboration for Research in Intensive Care

CTU, Copenhagen Trial Unit

IDMSC, Independent Data Monitoring and Safety Committee

eCRF, Electronic Case Report Form

EudraCT, European Union Drug Regulating Authorities Clinical Trial

GI, Gastrointestinal

GLM, Generalised Linear Model

GRADE, Grading of Recommendations Assessment, Development and Evaluation

HR, Hazard Ratio

HRQoL, Health Related Quality of Life

ICH-GCP, International Conference on Harmonisation on Good-Clinical -Practice

ICMJE, International Committee of Medical Journal Editors

ICU, Intensive Care Unit

IL-6, Interleukin 6

IQR, Interquartile Range

ITT, Intention-to-treat

IV, Intravenous

MD, Mean Difference

OR, Odds Ratio

QoL, Quality of Life

RCT, Randomised clinical trial

RR, Relative Risk

RRI, Relative Risk Increase

RRR, Relative Risk Reduction

SAR, Serious Adverse Reaction

SAE, Serious Adverse Event

SARS, Severe Acute Respiratory Syndrome

SD, Standard Deviation

SR, Systematic review

SSC, Surviving Sepsis Campaign

SUSAR, Suspected Unexpected Serious Adverse Reaction

WHO, World Health Organization

### 22 Introduction and background

#### *22.1 Severe acute respiratory syndrome coronavirus 2/Coronavirus Disease 19*

In December 2019, the Wuhan Municipal Health Committee in China identified an outbreak of viral pneumonia cases of unknown cause (1). A novel coronavirus was soon identified as the cause of the disease (1). This novel virus has been named severe acute respiratory syndrome coronavirus 2 (SARS-CoV-2) (2) and the disease caused by the virus has been designated coronavirus disease 2019 (COVID-19) (3). Since the initial outbreak in China in December 2019, SARS-CoV-2 has spread globally and COVID-19 has been declared a pandemic by the World Health Organization (WHO)(4). Currently, the number of reported patients with COVID-19 and associated deaths are, as of July 14, 2020, more than 13.100.000 and 573.000, respectively (5). There are currently large outbreaks in the US, Brazil, Russia, and India with many severely ill patients admitted to hospitals and intensive care units (ICUs).

SARS-CoV-2 causes respiratory tract infection (6). The symptoms vary from mild to severe pneumonia and from mild to severe acute respiratory distress syndrome (ARDS) (6). Current estimates suggest that up to 40% of hospitalised COVID-19 patients develop ARDS (6-10). Further, 20-35% of those patients admitted to the ICU may develop septic shock (6, 8, 9, 11, 12). Both conditions are associated with high morbidity and mortality (6, 13).

#### *22.2 Corticosteroids in COVID-19*

The current care in COVID-19 is primarily supportive including oxygen, mechanical ventilation, and general intensive care (14). Many patients are treated with various antiviral drugs or immunomodulatory agents, including corticosteroids (15). Until recently, clinical equipoise existed regarding the use of systemic corticosteroids for COVID-19. The Surviving Sepsis Campaign guidelines on the management of critically ill adults with COVID-19 recommended use of low-dose corticosteroids for shock reversal over no use (weak recommendation, low quality of evidence) and use of corticosteroids over no use for those with ARDS (weak recommendation, low quality of evidence) (11). In contrast, the WHO and the Infectious Diseases Society of America (IDSA) recommended against the use corticosteroids in COVID-19 (16, 17).

A preliminary report from the Randomised Evaluation of COVid-19 thERapY (RECOVERY) trial was released on June 22, 2020 (18). In the RECOVERY trial, 6,425 hospitalised patients with suspected or confirmed COVID-19 were randomised to open-label dexamethasone 6 mg daily for up to 10 days vs. usual care (18). The preprint of the preliminary results reported an overall relative reduction of 17% in 28-day mortality (age-adjusted rate ratio 0.83, 95% confidence interval (CI) 0.74 to 0.92) with indications of greatest benefit among those patients requiring invasive mechanical ventilation (rate ratio 0.65, 95% CI 0.51 to 0.82) (18). These results are supported by similar findings in a recently updated systematic review including patients with non-COVID-19 ARDS (risk ratio (RR) 0.72, 95% CI 0.55 to 0.93) (19).

International collaborative research initiatives have been formed with the aim of harmonising and coordinating data collection to enable prospective meta-analyses of the ongoing randomised trials of corticosteroids for COVID-19. The results of these meta-analyses are still not available. As of June 22, 2020, 16 trials assessing corticosteroids for COVID-19 were registered at ClinicalTrials.gov, many of which have already commenced enrolment (18, 20-34). Of these, the COVID STEROID trial is initiated by the same Sponsor and Management Committee of the COVID STEROID 2 trial (31). The COVID STEROID trial assesses low-dose hydrocortisone 200 mg daily vs. placebo in patients with COVID-19 and severe hypoxia. The trial was commenced on April 15, 2020, but paused after randomising 30 patients on June 16, 2020, due to the press release from the RECOVERY trial (35). The decision to continue or stop the COVID STEROID trial will be made after the peer-reviewed publication of the RECOVERY trial is available as well as results from an ongoing prospective meta-analysis of trials assessing corticosteroids for COVID-19.

#### *22.3 Type and dose of corticosteroids for sepsis, ARDS and COVID-19*

The choice of type, dose, and duration of corticosteroids for treatment of sepsis and ARDS is controversial. Various regimens have been used in different trials (Table 1). Generally, studies with short-course high-dose corticosteroids for sepsis did not show a reduction in mortality or showed increased mortality, whereas studies employing longer-course low-dose steroids showed shock reversal and potentially also a reduction in mortality (36). Clinical guidelines published in 2018 stated that the optimal corticosteroid dose and duration of treatment are still uncertain (37). A later dose-response meta-analysis suggested that long-course (7 days) low-dose (200–300 mg per day) hydrocortisone treatment with cumulative dose ≥1,000 mg was beneficial for the reduction of 28-day mortality in patients with sepsis (38).

Similarly, a meta-analysis of corticosteroids for ARDS was inconclusive regarding short-course high-dose treatment (>30 mg dexamethasone or equivalent per day), whereas early initiation of longer course low-dose corticosteroids (≤30 mg dexamethasone or equivalent per day) reduced the duration of mechanical ventilation and mortality (39).

In the RECOVERY trial, low-dose dexamethasone (6 mg) versus no intervention was shown to reduce 28-day mortality in hospitalised patients with suspected or confirmed COVID-19 (18). The findings from the RECOVERY trial have been implemented in a COVID-19 treatment guideline from the National Institutes of Health (NIH) and the updated guideline from IDSA in which dexamethasone 6 mg is recommended for COVID-19 patients receiving supplemental oxygen or mechanical ventilation (40, 41). Therefore, dexamethasone 6 mg is likely to be part of the standard care of COVID-19 patients receiving supplemental oxygen or mechanical ventilation in most hospitals as observed in our clinical practice survey (results below). The remaining ongoing trials of corticosteroids for COVID-19 assess different corticosteroids (i.e. dexamethasone, methylprednisolone, hydrocortisone, or prednisone) with varying daily doses used (median dexamethasone equivalent dose 15 mg, interquartile range (IQR) 10-16 mg) (20-34). However, the results of these trials have not yet been published (20-34), leaving the optimal dosing for COVID-19 uncertain.

In trials in non-COVID-19 ARDS, the doses used (median dexamethasone equivalent dose 12 mg, IQR 9-16 mg (42-48)) have been within the dosing regimens used in the COVID-19 trials. Of note, higher doses of dexamethasone has previously been used in a clinical trial in non-COVID ARDS suggesting benefit and no obvious harm (42).

In short-term use in healthy volunteers, dose-dependent activation of the corticosteroid receptor has been observed for increasing doses up to 60 mg of prednisone (equivalent to 12 mg of dexamethasone) suggesting that doses up to 12 mg of dexamethasone may offer additional anti-inflammatory effects (49). In that study, the adverse effects were independent of the dosing (49).

We, the COVID STEROID 2 trial investigators, have done a survey of clinical practice in early July 2020 at 26 potential COVID STEROID 2 trial sites after the preprint publication of the RECOVERY trial results. All sites had used corticosteroids for patients with COVID-19; at most sites (95%), all patients had received corticosteroids. Most sites used mainly dexamethasone, and the median steroid dose (in dexamethasone equivalents) used at sites in patients with COVID-19 was 9.6 mg (IQR 6.0 – 15.0 mg).

In a concomitant survey of clinical preferences done early July 2020 among doctors at potential COVID STEROID 2 trial sites after the preprint publication of the RECOVERY trial results, 86% of 250 responding doctors would always or most times use steroids in patients with COVID-19 and hypoxia; 56% would use 6 mg of dexamethasone or equivalent, and 36% would use a dose above 6 mg (unpublished results). As for preferences for an upcoming trial, most doctors (95%) would enrol their patients with severe COVID-19 into a trial of steroids, and most (55%) into one of 12 mg vs. 6 mg dexamethasone (unpublished results).

**Type and dose of corticosteroid in the COVID STEROID 2 trial**

For the COVID STEROID 2 trial, participants in the experimental intervention arm will receive intravenous dexamethasone 12 mg for up to 10 days or until discharge from the participating trial site without tapering. Dexamethasone has previously been used without tapering in a clinical trial assessing an even higher dose of dexamethasone (median 15 mg for 10 days) for non-COVID ARDS with potential benefit and without obvious harm (42).

Participants in the control intervention arm will receive intravenous dexamethasone 6 mg daily for up to 10 days or until discharge from the participating trial site without tapering, which is the exact protocol used in the RECOVERY trial (18). The RECOVERY trial investigators have not yet reported data on adverse events (18).

**Table 1. Estimates on the effects of corticosteroid vs. placebo/no treatment in critically ill patients with severe infection and/or severe respiratory failure:** Most data are from recently updated systematic reviews (SRs) of randomised clinical trials (RCTs), except those from viral acute respiratory distress syndrome (ARDS) and COVID-19.

| | Septic shock (50) | ARDS without COVID-19 (19) | Community acquired pneumonia (51) | Viral ARDS (11) | COVID-19(18) |
|---|---|---|---|---|---|
| Evidence base | SR of 22 RCTs, including 7297 participants | SR of 7 RCTs, including 851 participants | SR of 13 RCTs, including 2005 participants | SR of 10 observational studies on other corona viruses | Predefined subgroup of patients receiving invasive mechanical ventilation in 1 RCT, including 1007 participants in this subgroup |
| Corticosteroid used | Hydrocortisone 18 trials<br>Methylprednisolone 2 trials | Methylprednisolone 3 trials<br>Hydrocortisone 2 trials<br>Inhaled budesonide 2 trials<br>Dexamethasone 1 trial | Hydrocortisone 6 trials<br>Methylprednisolone or prednisolone 5 trials<br>Dexamethasone 1 trials<br>Prednisone 1 trial | Not reported | Dexamethasone |
| Daily dose (dexamethasone-equivalent) | 7.5-11.3 mg/day | 4-32 mg/day | 7.5-15 mg/day | Not reported | 6 mg |
| Mortality | RR 0.98 [0.89 to 1.08] | RR 0.72 [0.55 to 0.93] | RR 0.67 [0.45 to 1.01] | OR 0.83 [0.32 to 2.17] | Rate ratio 0.65 [0.51 to 0.82] |
| Admission to ICU | - | - | RR 0.69 [0.46 to 1.03] | | |
| Need for ventilation | - | - | RR 0.45 [0.26 to 0.79] | | |
| Days ventilated | MD −0.75 [−1.34 to -0.17] days | MD –4.8 [–7.0 to –2.6] days | - | | |
| Days in shock | MD −1.52 [−1.71 to -1.32] days | - | - | | |
| Days in ICU | MD −0.75 [−1.34 to −0.17] days | MD 0.1 [–3.0 to 3.2] days | - | | |
| Days in hospital | MD −0.87 [−2.17 to 0.44] days | MD –3.6 [–7.2 to –0.02] days | MD -1.22 [-2.08 to -0.35] days | | |
| Adverse events<br>Secondary infections<br>Hyperglycaemia<br>Gastrointestinal bleeding | <br>RR 1.05 [0.95 to 1.16]<br>RR 1.11 [1.07 to 1.16]<br>RR 1.09 [0.80 to 1.46] | <br>RR 0.82 [0.67 to 1.02]<br>RR 1.12 [1.01 to 1.24]<br>RR 0.71 [0.30 to 1.73] | <br>-<br>RR 1.49 [1.01 to 2.19]<br>RR 0.82 [0.33 to 1.62] | | |

| | | | |
|---|---|---|---|
| Encephalopathy<br>Neuromuscular weakness | RR 1.99 [0.37 to 10.84]<br>- | -<br>RR 0.85 [0.62 to 1.18] | RR 1.65 [0.88 to 3.08]<br>- |

ARDS: acute respiratory distress syndrome; SR: systematic review; RCTs: randomised clinical trials; mg: milligrams; RR: relative risk; OR: odds ratio; HR: hazard ratio; MD: mean difference; ICU: intensive care unit

#### *22.4 Ethical justification and trial rationale*

Patients with COVID-19 and severe hypoxia (the hallmark of ARDS) are at high risk of death (6, 7). Until recently, the care for these patients was exclusively supportive, including respiratory and circulatory support.

The RECOVERY trial reported a reduction in 28-day mortality with low-dose dexamethasone (6 mg) for hospitalised patients with suspected or confirmed COVID-19. Yet, higher doses of dexamethasone (median 15 mg) may be beneficial in non-COVID-19 ARDS (42), and higher doses were also used in the other trials of corticosteroids in COVID-19 (median dose 12 mg). Also, in a contemporary survey of the COVID STEROID 2 trial sites after the preprint publication of the RECOVERY trial had been published, 95% of sites used steroids in all patients, most often as dexamethasone in doses above 6 mg (median 9.6 mg (IQR 6.0 to 15.0)), a result supported by those of a concomitant survey of clinician's preferences (unpublished results). Taken together, it is unclear which dose of dexamethasone is most beneficial to COVID-19, and clinical equipoise exists among clinicians and researchers.

The present trial will be conducted to the highest of methodological standards with ongoing assessment of the known serious adverse reactions to corticosteroid, including a planned interim analysis. Any serious adverse reactions for single participants and the group of participants receiving higher vs. lower dose of dexamethasone will be assessed and handled. The control group will receive the exact same protocol as in the RECOVERY trial in addition to usual clinical care. We, the COVID STEROID 2 trial group, find the trial justifiable both medically and ethically.

The patients to be enrolled in the COVID STEROID 2 trial cannot consent due to the combination of severe infection and severe hypoxia. COVID-19 with severe hypoxia is a medical emergency that requires immediate interventions including life-supportive interventions. Therefore, we cannot delay enrolment and need to use the consent procedures for emergency research.

Informed consent will be obtained according to national law in the participating countries. In Denmark, we will use the consent procedures for temporarily incompetent patients for all patients enrolled in the COVID STEROID 2 trial (https://www.retsinformation.dk/Forms/R0710.aspx?id=192671). Here, patients will be enrolled after informed consent from a doctor (first trial guardian), who is independent of the trial, who has knowledge of the clinical condition and who is familiar with the trial protocol to such extent that he/she can judge for each patient, if it will be reasonable to enrol the patient in the trial. As soon as possible after enrolment, consent will be obtained from the patient's next of kin and another doctor (second trial guardian). The second trial guardian is also independent of the trial, has knowledge of the clinical condition, and is familiar with the trial protocol to such extent that he/she can judge for each patient, if it will be reasonable to enrol the patient in the trial. Participants, who regain competence, will be asked for informed consent as soon as possible (Appendix 6, 18.6). The process leading to informed consent will follow all applicable regulations. The consenting parties will be provided with written and oral information about the trial allowing them to make an informed decision about participation in the trial. Written information and the consent form will be subject to review and approval by the ethical committee system. The consenting party can at any time, without further explanation, withdraw consent.

#### *22.5 Trial conduct*

The COVID STEROID 2 trial will comply with the published trial protocol, the Helsinki Declaration in its latest version (52), the International Conference on Harmonization on Good-Clinical-Practice (GCP) guidelines (53), General Data Protection Regulation, and national laws (including Databeskyttelsesloven in Denmark). The Management Committee of the trial will oversee the conduct. We have written the protocol in accordance with the Standard Protocol Items: Recommendations for Interventional Trials (SPIRIT) 2013 Statement (54) and will register the trial in the ClinicalTrials.gov and European Union Drug Regulating Authorities Clinical Trials (EudraCT) registries before the enrolment of the first participant. No substantial deviation from the protocol will be implemented without prior review and approval of the regulatory authorities except where it may be necessary to eliminate an immediate hazard to the trial participants. In such case, the deviation will be reported to the authorities within 7 days.

Enrolment will start after the approval by the Ethics Committee, the Danish Medicines Agency and the Capital Region Knowledge Center for Data Compliance (legal department). We will publish the approved protocol online at the Collaboration of Research in Intensive Care's website at www.cric.nu and submit a manuscript with main points of the protocol including description of design, rationale and the detailed statistical analysis plan to a peer-reviewed medical journal.

### 23 Trial objectives

The objective of the *Higher vs. Lower Doses of Dexamethasone in Patients with COVID-19 and Severe Hypoxia – COVID STEROID 2 trial* is to assess the effects 12 mg vs. 6 mg of intravenous dexamethasone on the number of days alive without life-support and other patient-centered outcomes in adult patients with COVID-19 and severe hypoxia. We hypothesise that dexamethasone 12 mg will increase the number of days alive without life support as compared to dexamethasone 6 mg in patients with COVID-19 and severe hypoxia.

### 24 Trial design

The COVID STEROID 2 trial is an investigator-initiated, international, parallel-group, blinded, centrally randomised, stratified, clinical trial.

#### *24.1 Randomisation*

Patients with COVID-19 fulfilling all inclusion criteria and no exclusion criteria will be randomised. The 1:1 randomisation will be centralised and web-based according to the computer-generated allocation sequence list, stratification variable (trial site, the use of invasive mechanical ventilation (y/n), age below 70 years (y/n)), and varying block size at Copenhagen Trial Unit (CTU) to allow immediate and concealed allocation to one of the two intervention groups. The allocation sequence list will exclusively be known to the data manager at CTU and will be unknown the unblinded trial site staff preparing the trial medication (section 6.2), to the clinicians, to the investigators and statistician conducting the analysis. Each trial participant will be allocated a unique screening number.

#### *24.2 Blinding*

We will mask the allocation for the participants, the clinical staff, the trial site staff registering the outcome data, the trial Management Committee, and the trial statistician, who will conduct the analyses with the two intervention groups coded as e.g. 0 and 1. A dedicated team of trial site staff (medical-, pharmacy- or nurse students or pharmacists, research nurses or doctors) who are certified in medicine handling procedures will unblinded prepare the trial medication and perform daily data entry about the administration of the trial medications including any protocol violations. This unblinded team of trial site staff will not be involved in the care of trial participants, outcome assessment, or in the statistical analyses. They will be instructed not to reveal the allocation under any circumstances.

**Trial medication preparation**

We will use shelf-medications from the hospital department's pharmacy for both intervention and control medication. The local trade names used in the COVID STEROID 2 trial are presented in Appendix 9, 18.9.

To ensure blinding, the trial medications will be prepared by the unblinded trial site staff, and the participants and clinical staff will thus remain blinded to the treatment allocation. For each participant, the trial medication will be prepared once daily and administered as a bolus injection.

**Preparation of experimental intervention: dexamethasone 12 mg**

The experimental intervention is dexamethasone 12 mg daily given as bolus injection for up to 10 days. We will allow the use of betamethasone 12 mg at sites where dexamethasone is not available, as these are diastereomers and likely equipotent (55).

Dexamethasone phosphate is a clear colourless solution and comes in vial of 1 and 5 ml (4 mg per ml, which equals 3.33 mg of dexamethasone). For each participant allocated to the experimental intervention, 3.6 ml of dexamethasone phosphate will be drawn into one 5 ml syringe together with 1.4 ml isotonic saline (0.9%) to a total volume of 5 ml and a concentration of 2.88 mg/ml of dexamethasone phosphate, which equals a total of 12 mg of dexamethasone. The trial medication will be delivered daily to the clinical staff by the unblinded trial staff and administered as one bolus injection.

**Preparation of control intervention: dexamethasone 6 mg**

The control intervention is dexamethasone 6 mg daily given as bolus injection for up to 10 days. We will allow the use of betamethasone 6 mg at sites where dexamethasone is not available, as these are diastereomers and likely equipotent (55).

For each participant allocated to the control intervention, 1.8 ml of dexamethasone phosphate (4 mg per ml, which equals 3.33 mg of dexamethasone) will be mixed with 3.2 ml of isotonic saline (0.9%) to a total volume of 5 ml and a concentration of 1.44 mg/ml of dexamethasone phosphate, which equals a total of 6 mg of dexamethasone. The dexamethasone solution will be delivered daily to the clinical staff by the unblinded trial staff and administered as one bolus injection.

#### *24.3 Unblinding*

**Unblinding of the intervention for a participant**

The intervention may be unblinded if deemed necessary by the treating clinician or the investigator for treatment or safety reasons. The sponsor or his delegate will break the blind for a participant if there is clinical suspicion of an unexpected serious adverse reaction (SUSAR) and judge the 'expectedness' of this according to the product information. Any SUSAR will be reported to the authorities accordingly.

Unblinding of the intervention for a participant can be performed around the clock by contacting the sponsor or his delegate. The sponsor or his delegate will contact the unblinded trial site staff from whom the trial allocation is available, and the intervention will be discontinued. The primary investigator at the site will be informed about the participant's allocation.

**Unblinding of the entire trial**

The Management Committee may stop and unblind the trial if there are clear indications that one intervention is superior to the other based on the recommendations from the independent Data Monitoring and Safety Committee (IDMSC) or other relevant data.

The members of the IDMSC will remain blinded unless 1) they request otherwise or 2) the interim analysis has provided strong indications of one intervention being beneficial or harmful.

#### *24.4 Participant timeline*

We will strive to enrol participants as soon as they fulfil the inclusion criteria, and no later than within 5 days of initiation of standard care corticosteroids for COVID-19. The allocated intervention will be continued so that participants in total receives 10 days of corticosteroids or until discharge from the participating site or death (whichever occurs first). Thus, no participant will receive corticosteroid for COVID-19 for more than 10 consecutive days. We will follow the patients for 28 days after randomisation and identify survivors at days 90 and 180 in electronic patient records or in registries. At day 180, we will contact surviving participants or their next of kin for health-related quality of life (HRQoL) follow-up.

**End of trial**

The trial will end when the last patient enrolled has completed 180-day follow up (last-patient last-visit). We will report the end-of-trial no later than 90 days after the last-patient last-visit to the Danish Medicines Agency and Ethics Committee.

### 25 Selection of participants

All patients admitted to an active trial site will be considered for participation. Patients will be eligible if they comply with the inclusion and exclusion criteria (full definitions are presented in Appendix 3, 18.3).

#### *25.1 Inclusion criteria*

All the following criteria must be fulfilled:

- Aged 18 years or above **AND**
- Confirmed SARS-CoV-2 (COVID-19) requiring hospitalisation **AND**
- Use of one of the following:
    - Invasive mechanical ventilation **OR**
    - Non-invasive ventilation or continuous use of continuous positive airway pressure (CPAP) for hypoxia **OR**
    - Oxygen supplementation with an oxygen flow of at least 10 L/min independent of delivery system

#### *25.2 Exclusion criteria*

We will exclude patients who fulfil any of the following criteria:

- Use of systemic corticosteroids for other indications than COVID-19 in doses higher than 6 mg dexamethasone equivalents
- Use of systemic corticosteroids for COVID-19 for 5 days consecutive days or more

- Invasive fungal infection
- Active tuberculosis
- Fertile woman (<60 years of age) with positive urine human gonadotropin (hCG) or plasma-hCG
- Known hypersensitivity to dexamethasone
- Previously randomised into the COVID STEROID 2 trial
- Informed consent not obtainable

We will not exclude patients enrolled in other interventional trials unless the protocols of the two trials collide. We will establish co-enrolment agreements when possible with the sponsor/investigator to maintain an updated list of trials approved for co-enrolment (Appendix 7, 18.7).

#### *25.3 Participant discontinuation and withdrawal*

The procedure for handling withdrawal of consent from a participant will follow national regulations. In Denmark, the procedure will be as follows.

**Discontinuation and withdrawal at the choice of the participant or the proxy**

A participant, who no longer wishes to participate in the trial, can withdraw his/her consent at any time without need of further explanation, and without consequences for further treatment.

For incapacitated participants, consent can be withdrawn at any time by the person(s), who has given proxy-consent. To limit the amount of missing data, we will collect as much data as possible from each participant. Therefore, the investigator will ask the participant or the proxy if they allow continued data registration and follow-up at day 180.

**Discontinuation and withdrawal at the choice of the investigator**

A participant may have the intervention stopped by the clinician or investigator at any time, if:

- The participant experiences intolerable adverse reactions or events (including Serious Adverse Reactions (SARs) or Suspected Unexpected Serious Adverse Reactions (SUSARs)) suspected to be related to the trial intervention.
- The clinicians in conjunction with the coordinating investigator decide it to be in the interest of the participant

- Withdrawal from active therapy
- The participant is subject to compulsory hospitalisation.

In these participants, the collection of data and the follow-up will continue, and the participant will remain in the intention-to-treat population.

**Discharge**
The trial allocation will be stopped when patients are discharged or transferred to a non-participating hospital department. The patient will still be followed through the electronic health records, including registration of data for days alive without life support and day alive and out of hospital. Participants who are discharged or transferred to a department participating in the COVID STEROID 2 trial will continue the allocated intervention at the new trial site for a total treatment duration of 10 days from randomisation. If the participant is readmitted to a COVID STEROID 2 trial site from a non-participating hospital department within 10 days of randomisation, the allocation will also resume for a total treatment duration of up to 10 days from randomisation depending on the number of days with corticosteroid treatment before randomisation.

### 26 Selection of trial sites and personnel

#### *26.1 Trial sites and setting*

Trial sites will be hospitals in Denmark, Sweden, Switzerland and India. Trial sites are listed in the section *Administrative information* (p. 4). This section will be updated during the trial, and authorities will be notified.

#### *26.2 Trial personnel*

All clinical staff caring for patients will be eligible to care for and give the interventions to the trial participants. The primary trial personnel are constituted of a dedicated team of medical-, pharmacy- or nurse students or research nurses or doctors who will be trained and certified in all trial-related procedures. The screening will be done by the clinical doctors. When a candidate patient is identified, the clinical team will alert the trial staff, who will seek consent and thereafter screen the patient in the eCRF.

Medical students will be eligible to screen and enrol patients in the eCRF, obtain informed consent, prepare trial medication and perform data entry. Nurse and pharmacy students and pharmacists will be eligible to obtain informed consent, prepare trial medication and perform data entry; nurse- and pharmacy students and pharmacists can only screen and enrol of patients in the eCRF if a named doctor or medical student checks and signs the inclusion notes. All participating trial sites will receive written and oral instructions about the trial procedures. A 24-hour hotline will be available for trial-related questions.

#### 26.3 Trial interventions

The intervention period is up to 10 days from randomisation or until hospital discharge or death, whichever comes first. The intervention period will be adjusted for each participant so that the number of consecutive days with the use of corticosteroid for COVID-19 before randomisation is subtracted from the 10-day intervention period (e.g. a participant who has received corticosteroid for COVID-19 for 3 consecutive days prior to randomisation will receive 7 days of the trial intervention).

#### 26.4 Experimental intervention

Intravenous bolus injection of dexamethasone 12 mg. We will allow the use of betamethasone 12 mg at sites, where dexamethasone is not available.

#### 26.5 Control intervention

Intravenous bolus injection of dexamethasone 6 mg. We will allow the use of betamethasone 6 mg at sites, where dexamethasone is not available.

#### 26.6 Co-interventions

All participants in the trial will be given co-interventions at discretion of the treating clinicians. We will recommend against the use of additional corticosteroids (systemically or as inhalation) and other anti-inflammatory agents (e.g. IL-6 inhibitors) in all trial participants.

Based upon an updated critical appraisal of the literature, the Management Committee endorses and encourages co-enrolment in the COVID STEROID 2 trial (Appendix 7, 18.7). Co-enrolment agreements will be established when possible with the sponsor/investigator to maintain an updated list of trials approved for co-enrolment (Appendix 7, 18.7).

#### 26.7 Concomitant interventions

All other interventions will be allowed as per the clinical team including those affecting CYP3A4, because it is not clinical practice at the trial sites to change the use or dosing of dexamethasone or betamethasone with concomitant use of CYP3A4 inhibitors or inducers.

#### 26.8 Monitoring of participants

The participant will be monitored closely due to the severity of their illness. The level of monitoring will be as per the clinical standard of the trial sites including continuous monitoring of oxygen saturation and pulse when severe hypoxia is present; 1-2 hourly measurements of blood pressure and respiratory rate when severe hypoxia is present; and 8-hourly measurement of body temperature; daily measurement of blood values including C-reactive protein (CRP), leukocyte count, hemoglobin, creatinine, urea and electrolytes, pH, atrial blood gases, lactate, and blood glucose. Additional measurements will be done on clinical indications including microbiological cultures, markers of candida infections and electrocardiograms (ECGs).
These data will not be registered in the COVID STEROID 2 trial eCRF but will be available in the participant's health care records for the Sponsor and/or the authorities if needed.

#### 26.9 Criteria for modification of interventions for a given trial participant

The clinical team may at any time violate the protocol if they find it to be in the best interest of the participant. We will have a COVID STEROID 2 trial hotline to enable discussion around-the-clock between the clinicians caring for trial participants and the COVID STEROID 2 trial team regarding protocol related issues. Protocol violations will be registered and reported.

#### 26.10 Assessment of participant compliance

We will monitor protocol compliance at the trial site through the electronic case report form (eCRF) and alert trial sites in the case of clear violations (central monitoring). In addition, the trial will be externally monitored according to the GCP Directive and the monitoring plan (section 13).

#### 26.11 Intervention accountability

Both the trial intervention and control medications are routinely used for in-hospital treatment of patients and we will use shelf-medication from the department's pharmacy. The trial medication will only be handled by the trained trial staff and the clinical staff who are trained and certified for the caring for patients. The methods used for trial medication preparations are described in 6.2.

**Trial medications**

The list of local brands used in Denmark, Sweden, Switzerland and India are presented in Appendix 18.9.

<u>Experimental intervention</u>

Active medication: dexamethasone phosphate, solution for injection, 4 mg/ml, ATC code: H02AB02

At sites where dexamethasone is not available: betamethasone sodium phosphate, solution for injection, 5.3 mg/ml, ATC code: H02AB01

Solution: isotonic saline, solution for intravenous injection, 9mg/ml, ATC code: B05BB01. Each ml contains 9 mg of saline in sterile water. Content of electrolytes/l: 154mmol chloride, 154 mmol natrium. Osmolarity 308mmol/l.

<u>Control intervention</u>

Active medication: dexamethasone phosphate, solution for injection, 4 mg/ml ATC code: H02AB02

At sites where dexamethasone is not available: betamethasone sodium phosphate, solution for injection, 5.3 mg/ml, ATC code: H02AB01

Solution: isotonic saline, solution for intravenous injection, 9mg/ml, ATC code: B05BB01. Each ml contains 9 mg of saline in sterile water. Content of electrolytes/l: 154mmol chloride, 154 mmol natrium. Osmolarity 308mmol/l.

**Labelling**

When the trial drug is prepared, it will be labelled with a COVID STEROID 2-trial sticker, making clinical personnel aware that the syringe contains trial medication. The sticker will hold information about the participant's data, the trial medicines, the date and time of preparation, the expire date and time, the signature of the trial staff preparing the medications and a telephone number for the COVID STEROID 2-trial 24-h hotline (the labels are presented in Appendix 4, 18.4). To ensure blinding of the clinicians, the sticker will not hold information about the BATCH / LOT numbers of the trial medications. The BATCH / LOT numbers will instead be noted in a trial medication log. This log will only be available to the unblinded research staff.

### 27 Outcome measures

#### *27.1 Primary outcome*

Days alive without life support (i.e. invasive mechanical ventilation, circulatory support or renal replacement therapy (including days in between intermittent renal replacement therapy)) from randomisation to day 28.

#### *27.2 Secondary outcomes*

- Number of participants with one or more serious adverse reactions (SARs) at day 28 defined as new episodes of septic shock, invasive fungal infection, clinically important gastrointestinal (GI) bleeding or anaphylactic reaction to IV dexamethasone
- All-cause mortality at day 28
- All-cause mortality at day 90
- Days alive without life support at day 90
- Days alive and out of hospital at day 90
- All-cause mortality at day 180

- HRQoL at day 180 using EQ-5D-5L and EQ-VAS

### 28 Safety

#### *28.1 Definitions*

In the COVID STEROID 2 trial, we will use the definitions below (56):

**Adverse event (AE)**

Any undesirable medical event occurring to a participant during a clinical trial, which does not necessarily have a causal relationship with the intervention.

**Adverse reaction (AR)**

Any undesirable and unintended medical response related to the intervention occurring to a participant during a clinical trial.

**Serious adverse event (SAE)**

Any adverse event that results in death, is life-threatening, requires hospitalisation or prolongation of existing hospitalisation, results in persistent or significant disability or incapacity, or is a congenital anomaly or birth defect.

**Serious adverse reaction (SAR)**

Any adverse reaction that results in death, is life-threatening, requires hospitalisation or prolongation of existing hospitalisation, results in persistent or significant disability or incapacity, or is a congenital anomaly or birth defect. The SARs are identified in the Danish Summary of Products Characteristics (SmPC) for dexamethasone.

**Suspected unexpected serious adverse reaction (SUSAR)**

Any suspected adverse reaction which is both serious and unexpected (the nature or severity of which is not consistent with SmPC for dexamethasone).

#### *28.2 Risk and safety issues in the COVID STEROID 2 trial*

The trial participants will be hospitalised patients for whom adverse events and reactions are documented routinely in the patient health record (i.e. notes, charges and laboratory reports). We will record the occurrence of SARs in the 28 days following randomisation for all participants and report them as an outcome measure.

For all participants, we will register daily the presence or absence of potential SARs according to intravenous dexamethasone in the Danish SmPC, which are serious and relevant to short course use in critically ill patients, i.e. new episodes of septic shock, invasive fungal infections, clinically important GI bleeding and anaphylaxis.

#### *28.3 Assessment of adverse events*

**Timing**

In all participants, we will assess the occurrence of SARs in the 28 days following randomisation (the maximum intervention period is 10 days; 28 days allow for at least another 18 days of assessment after the intervention, which is clinically relevant in short course use in critically ill patients.

**Classification of an event**

We will make no inferences about a causal relationship between the intervention and the SARs but register the occurrence in the two groups and report them in the final report according to the definition given above.

The investigators will classify SAEs (as per the definition above occurring within 28 days from randomisation) and report them to the sponsor. If such a SAE is deemed both unexpected and related to the intervention by the investigator, it will be considered a SUSAR and reported as such. If the sponsor does not adjudicate the SAE as related to the intervention, this will also be noted in the SUSAR report.

**Reporting**

Any SAE adjudicated to be unexpected or related to the trial intervention by the investigator, will be reported within 24 hours to the sponsor or his delegate. If deemed a SUSAR by the sponsor, he will report it to the Danish Medicine Agency, the Ethics Committee and all trial sites within 7 days. No later than 8 days after the reporting, the Sponsor will inform the Danish Medicines Agency of relevant information on the Sponsor's and the investigator's follow-up action to the life-threatening or fatal SUSAR. Any other SUSARs will be reported to the Danish Medicines Agency no later than 15 days from the time when the Sponsor is informed.

In Appendix 10 (section 18.10), SAEs seen frequently in critically ill patients with COVID-19 and/or critically ill patients in general are listed. The listed SAEs do not have to be reported to the sponsor within 24 hours of occurrence if adjudicated not to be related to the intervention and expected in the patent population. All SAEs not listed in Appendix 10 will be reported to sponsor within 24 hours of occurrence.

Once a year, the sponsor will submit a list of all SARs that have occurred at all sites during the trial period and a report on safety of the trial subjects to the Danish Medicines Agency and National Ethics Committee.

The sponsor will notify the Danish Medicines Agency when the trial has been completed (no later than 90 days thereafter) and if earlier than planned, the reasons for stopping the trial.

In addition, we will report all SARs defined in 9.2 as outcome measures and all SUSARs in the final trial report and the results of the trial will be reported on EudraCT within 12 months of 'last-patient-last-visit'.

### 29 Procedures, assessments and data collection

#### *29.1 Screening*

All patients admitted to a participating trial site with confirmed COVID-19 and severe hypoxia (as defined in section 7.1) will be eligible for screening. The screening will be done by the clinical doctors. When a candidate patient is identified, the clinical team will alert the trial staff, who will seek consent and thereafter screen the patient in the eCRF.

For all fertile women under 60 years of age, screening for hCG in urine or plasma will be done before enrolment in the trial. If a hCG-test has already been done under the current admission, we will use the test result of this for screening for pregnancy.

#### *29.2 Procedures of informed consent*

Participants will be enrolled after consent by proxy is obtained according to national regulations. We will follow the normal procedures for collection of informed consent and any modifications to these as approved by the Ethics Committee due to restrictions in physical visits to the hospitals during the current SARS-CoV-2 epidemic. The procedure for informed consent in Denmark is described in Appendix 6 (18.6).

#### *29.3 Data collection*

The screening of participants will be done by the clinical team as described in 10.1. The clinicians will pass on information about eligible participants to the COVID STEROID 2 trial site staff who will hereafter obtain informed consent from the first trial guardian (the Danish procedure).

After informed consent is obtained, the data below (10.4) will be obtained by the trial site staff from the participant's hospital files, national/regional/hospital registers (source data as defined per site and region) and interview with participant or next of kin and entered into the web-based eCRF (the server hosting the database is located at CTU, Rigshospitalet, Region Hovedstaden). For participants transferred from a trial site to a non-trial site, data related to the outcomes will be collected from either hospital files (if accessible) or investigator contact to the non-trial site or health care registers.

#### *29.4 Variables*

All variables are defined in Appendix 3 (18.3).

**Screening variables**

Inclusion and exclusion criteria (7.2 and 7.3)

Number of consecutive days of systemic use of corticosteroids for COVID-19 up to the day of screening

**Baseline variables**

- Sex
- Age at enrolment (date of birth)
- Date of admission to hospital

- Number of days with symptoms before hospital admission
- Department at which the participant was included:

  - Emergency department
  - Hospital ward
  - Intermediate care unit
  - Intensive care unit

- Use of respiratory support at randomisation:

  - Closed system (y/n): Invasive mechanical ventilation or non-invasive ventilation or continuous use of CPAP
    - If yes, latest FiO2 prior to randomisation
    - If yes, no. of days of mechanical ventilation prior to randomisation
  - Open system with an oxygen flow ≥10 L/min (y/n)
    - If yes, maximum supplemental oxygen flow on an open system at randomisation (+/- 1 h)

- Limitations of care (i.e. not for invasive mechanical ventilation, circulatory support, renal replacement therapy, cardio-pulmonary resuscitation) at the time of randomisation (y/n)
- Chronic use of systemic corticosteroids for other indications than COVID-19 (y/n)
- Treatment for COVID-19 during current hospital admission prior to randomisation:
  - Agents with potential anti-viral action:
    - Remdesivir
    - Convalescent plasma
    - Other
  - Anti-bacterial agent (y/n)
  - Agents with potential anti-inflammatory action:
    - Janus kinase inhibitor(y/n)
    - IL-6 inhibitors (y/n)
    - Other
- Chronic co-morbidities:
  - History of ischaemic heart disease or heart failure (y/n)
  - Diabetes Mellitus (y/n)
  - Chronic pulmonary disease (y/n)
  - Immunosuppressive therapy within the last 3-months (y/n)
- Blood values, interventions and vital parameters:
  - Participant weight
  - $PaO_2$ and $SaO_2$ in the most recent arterial blood gas sample prior to inclusion **OR** $SpO_2$ from pulse oximeter if arterial blood gas sample is not available
  - Circulatory support (infusion of vasopressor/inotropes) within the last 24 hours prior to randomisation (y/n)
  - Renal replacement therapy within the last 72 hours prior to randomisation (y/n)
  - Highest plasma lactate within the last 24 hours prior to randomisation

**Daily during admission for the first 14 days after randomisation (day forms)**

- Invasive mechanical ventilation (y/n)
- Circulatory support (continuous infusion of vasopressor/inotropes for a minimum of 1 hour) on this day (y/n)
- Any form of renal replacement therapy on this day including days between intermittent renal replacement therapy (y/n)
- SAR(s) on this day (y/n for each)

- New episodes of septic shock
- Invasive fungal infection
- Clinically important GI bleeding
- Anaphylactic reaction to IV dexamethasone

**Daily registration of major protocol violations up to 10 days (from day 1 and up to 10 days)**

- Use of open-label systemic corticosteroids on this day (y/n)

-Trial intervention (y/n): did the participant receive the trial medication on this day? (yes/no)

- If no, apply reasons: by error/lack of resources, other reason

**Discharge form**

- Died in hospital
- Discharged from hospital
- Discharged to another ward participating in the COVID STEROID 2 trial
- Discharged to another ward not participating in the COVID STEROID 2 trial

**Follow-up 28 days after randomisation**

- Death (y/n, if yes: date of death)
- Number of days on invasive mechanical ventilation from day 15-28
- Number of days with circulatory support (infusion of vasopressor/inotropes for a minimum of 1 hour) from day 15-28
- Number of days on renal replacement therapy from day 15-28
- The occurrence of SAR(s) from day 15-28:
    - New episodes of septic shock (y/n, if yes: apply date(s))
    - Invasive fungal infection (y/n, if yes: apply date(s))
    - Clinically important GI bleeding (y/n, if yes: apply date(s))
    - Anaphylactic reaction to IV dexamethasone (y/n, if yes: apply date(s))
- Use of extracorporeal membrane oxygenation (ECMO) from randomisation to day 28 (y/n)
- Discharged against medical advice to home/other hospital/other facility (y/n)
    - If yes, apply medical condition at the time of discharge: on life support (i.e. invasive mechanical ventilation, circulatory support or renal replacement therapy), receiving supplementary oxygen (<10 L/min or ≥10 L/min) or no supportive therapy

**Follow-up 90 days after randomisation**

- Death (y/n, if yes date of death)
- Number of days on invasive mechanical ventilation from day 29-90
- Number of days with circulatory support (continuous infusion of vasopressor/inotropes for a minimum of 1 hour) from day 29-90
- Number of days on renal replacement therapy from day 29-90, including days between intermittent renal replacement therapy
- Date of discharge from hospital
- Additional hospital admissions (y/n, if yes: date of re-admission(s) and discharge(s))

**Follow-up 180 days after randomisation**

- Death (y/n, if yes date of death)
- HRQoL
  - EQ-5D-5L
  - EQ-VAS

### 30 Statistical plan and data analysis

The analyses will be done according to the principles stipulated in ICH-GCP guidelines (56). The protocol and detailed statistical analysis plan will be published online at www.cric.nu and in a peer-reviewed journal before the conduct of the planned interim analysis.

The primary outcome will be compared using the Kryger Jensen and Lange test and reported as differences in means and medians along with 95% confidence intervals adjusted for the stratification variables (site, use of invasive mechanical ventilation, and age). Secondary binary outcomes will be analysed using binomial regression models with log links adjusted for the stratification variables (site, use of invasive mechanical ventilation, and age) with results quantified as adjusted relative risks supplemented with adjusted risk differences, both with 95% confidence intervals.

#### 30.1 Sample size and power

**Sample size estimation and testing strategy**

At maximum, we will randomise 1,000 participants. A blinded statistician will conduct an interim analysis after the first 500 participants have been followed for 28 days. The alpha values for the interim analysis and the final analysis are 0.0054 and 0.0492, respectively as by the O'Brien-Fleming bounds, which preserves type I error at the usual 5% (57). In both analyses, a Kryger Jensen and Lange test will be employed to compare the groups on the primary outcome. The trial will be stopped early if the alpha cut-off is crossed at the interim analysis. The trial has 85% power to detect a 15% relative reduction in 28-day mortality combined with a 10% reduction in time on life support among the survivors.

The mortality outcomes will be tested in a hierarchical procedure along with the primary outcome (first primary outcome, then 28 days mortality, and finally 90 days mortality) reusing the alpha if the previous test was significant. If the primary outcome is insignificant at trial conclusion, ordinary 5% level test will be employed for all additional outcomes, but the results interpreted as exploratory.

**Power estimations for secondary outcomes**

We expect to have 80% statistical power to detect the following effects for the secondary outcomes based on the trial design described above. Power is reported at the 5% level even though the two mortality outcomes are also part of the primary outcome's hierarchical testing procedure.

- A 21% relative risk reduction for the mortality at day 28 (control event rate 30%)

- A 18% relative risk reduction for the mortality at day 90 (control event rate 40%)

- A 32% relative risk reduction for the number of participants with one or more SARs (control event rate 15%)
- A 15% relative risk reduction for the mortality at day 180 (control event rate 50%)

The estimates of control event rates for mortality at day 28 originate in data of previous coronavirus studies (6, 58); the estimates of the control event rates for mortality at day 90 and the number of participants with SARs are based on best clinical estimate. We expect the following secondary outcomes to be highly skewed (non-normally distributed): Days alive out of hospital at day 90 and

HRQoL at 180 days. The power estimations for these are, therefore, somewhat uncertain why we refrain from making these estimates.

#### *30.2 Statistical methods*

The analyses will be done in the intention-to-treat (ITT) population defined as all randomised participants for whom there is consent for the use of data.

The primary outcome will be compared using a Kryger Jensen and Lange test adjusted for the stratification variables (site, invasive mechanical ventilation, and age). Differences will be quantified as differences in means and medians along with 95% confidence intervals. For the binary outcomes (including mortality outcomes), we will use generalised linear models with log links and binomial error distributions adjusted for the stratification variables (site, invasive mechanical ventilation, and age) as the primary analysis (59). Differences in binary outcomes will be quantified using adjusted relative risks and secondarily adjusted risk differences along with 95% confidence intervals. This will be supplemented with Fisher's exact tests. Days alive without life support at day 90 and days alive out of hospital at day 90 will be analysed similarly to the primary outcome.

We will challenge the primary result in analyses adjusted for important baseline risk factors (age, co-morbidities, and use of life-support), in subgroups (Table 2) and in the per-protocol population being the ITT population except those having one or more major protocol violations as defined above (11.4 Variables). If there are more than 5% missing data for outcomes and/or covariates for an analysis, we will multiply impute the missing data for that analysis.

All statistical tests will be 2-tailed. Several outcome measures (including SARs and days alive without the use of life support at day 28 and 90) are composite; we will also report each component of these outcomes as recommended (56) in a supplement to the main report.

*Table 2. Heterogeneity of the intervention effects on the primary outcome will be analysed in the following subgroups based on baseline characteristics. As statistical test, we will use test of interaction in the adjusted analysis described above (p = 0.01).*

| Subgroup | Definition | Expected direction of the interaction |
|---|---|---|
| Elderly patients | Patients ≥70 years versus <70 years of age | Larger beneficial effect of higher dose dexamethasone in the younger population |
| Invasive mechanical ventilation | Patients who receive invasive mechanical ventilation versus oxygen by other delivery systems | Larger beneficial effect of higher dose dexamethasone in patients who receive invasive mechanical ventilation |
| Shock | Patients with shock versus without shock | Larger beneficial effect of higher dose dexamethasone in patients with shock |
| Duration of corticosteroid use before enrolment in COVID STEROID 2 trial | Patients who received corticosteroids for COVID-19 for 0 to 2 days versus 3 to 4 days before enrolment | Larger beneficial effect of higher dose dexamethasone in patients with short duration (≤2 days) of corticosteroid use before enrolment |
| Limitations of care | Patients with limitations of care (i.e. not for invasive mechanical ventilation, circulatory support, renal replacement therapy, cardio-pulmonary resuscitation) versus without limitations of care | Larger beneficial effect of higher dose dexamethasone in patients without limitations of care |
| Chronic use of systemic corticosteroids for other indications than COVID-19 | Patients with versus without chronic use of systemic corticosteroids | Larger beneficial effect of higher dose dexamethasone in patients without chronic use of systemic corticosteroids |

**Effect measures**

We will present the effects on the primary outcome as raw mean differences as well as median differences. For binary outcomes, we will report results as raw and adjusted relative risks and absolute risk differences, computed using generalized linear models (GLMs) with appropriate link functions (log links) and binomial error-distribution. Results will be presented with 95% confidence intervals (CI) for the analyses of the primary outcome (P-value 0.05) and 99% CIs for those of the secondary outcomes (P-value 0.01) due to the multiplicity of these. Significance of results will be based on the test described under testing strategy.

**Interim analysis**

We will conduct one interim-analyses after 500 participants have been followed for 28 days.

The IDMSC will analyse the primary outcome and the occurrence of SARs as described in the charter (Appendix 5, 18.5). The IDMSC will submit their recommendations to the Management Committee, which make the final decision regarding the continuing, pausing or stopping of the trial as described in the IDMSC charter.

After completion of the interim analysis, the recommendations from the IDMSC and the conclusion reached by the Management Committee will be submitted to the Ethics Committee.

**Early stopping criteria**

We will employ O'Brien-Fleming bounds which imply a significant cut-off of 0.0054 at the interim analysis. The Kryger Jensen and Lange test will be employed to compare the groups for the primary outcome. The trial will be stopped early if the alpha cut-off is crossed at the interim analysis.

**Final analysis**

Before unblinding the interventions groups, we will submit the statistical report of primary and secondary outcomes at day 28 (i.e. days alive without life support, mortality and SAR) to the IDMSC. The IDMSC will be asked to submit their recommendations to the Management Committee on whether to submit a primary report on 28-day outcomes or await the analyses of 90-day outcomes.

### 31 Quality control and quality assurance

The sponsor and his delegates will be responsible for organising the trial sites including education of the local investigators, the trial site staff and clinical staff before the initiation of the trial. This education will be continuously documented in the site master file.

After initiation, trial site investigators will be responsible for all trial-related procedures at their site, including education of staff in trial-related procedures, recruitment and follow-up of participants and entry of data. Clinical staff at the trial sites will be responsible for screening of eligible patients and the treatment of trial participants.

#### *31.1 Monitoring*

The trial will be externally monitored according to the GCP Directive and the monitoring and data verification plan including the documentation of informed consent of trial participants. The monitoring and data verification plan will be developed together with the GCP unit of Copenhagen University Hospital and adhered to by the staff monitoring all trial sites.

After the consent is obtained, Sponsor and his delegates will have access to the participants hospital files for quality control and monitoring. Sponsor will allow direct access to source data for GCP monitoring or control visits by the Danish national authorities overseeing drug trials. In addition, we will use central monitoring of site through the eCRF, including adherence to the protocol.

#### *31.2 Drug traceability measures*

The registration of the batch numbers and the expiry dates of the dexamethasone and saline used, and the identity of the clinician administering the dexamethasone and saline will be registered as per standard practice at the sites. These data will not be registered in the trial documents but can be obtained by the Sponsor or the authorities if needed. We believe that this is a safe procedure because both the dexamethasone and saline used in the COVID STEROID 2 trial has been in clinical use for many years and the safety of single doses cannot be questioned. The same procedure was approved by the Danish Medicines Agency in the CLASSIC (EudraCT no. 2018-000404-42) and COVID STEROID trials (EudraCT no. 2020-001395-15).

### 32 Legal and organisational aspects

#### *32.1 Finance*

**Trial funding**

The trial is funded by grants from the Novo Nordisk Foundation (DKK 5.000.000,-) and Rigshospitalet (DKK 1.875.000,-). The funding organisation has not been or will not be involved in the design, conduct, analyses, or reporting of the trial nor will it have ownership of the data. The Sponsor and trial staff have no financial affiliations to the Novo Nordisk Foundation.

**Compensation**

Dependent on the workload and preferences, the trial sites will receive case money or funds to the salary for the dedicated team of trial staff.

**Insurance**

In Denmark, the trial participants are covered by the Danish Law 'Lov om Patientskadeerstatning'; in Sweden, by the 'Läkemedelsförsäkringen'; and in Switzerland, by the participating hospital's insurance. In India, insurance will be covered by the local sponsor (The George Institute for Global Health, India).

### 33 Plan for publication, authorship and dissemination

All trial results, whether positive, negative or neutral, will be published preferably in a peer-reviewed medical journal. Furthermore, the results will be published at the Collaboration for Research in Intensive Care (CRIC) home page (www.cric.nu). We will adhere to the Consolidated Standards of Reporting Trials (CONSORT) statement (60), including the accountability of all patients screened (Appendix 2, 18.2).

Before unblinding the intervention groups, the Management Committee will write two abstracts based on the statistical report with the group allocation masked, one assuming the experimental intervention group is X and the control intervention group is Y, and one assuming the opposite. Then, the allocation code will be unmasked.

Authorship will be granted according to the guidelines from the International Committee for Medical Journal Editors (ICMJE; http://www.icmje.org/recommendations/browse/roles-and-responsibilities/defining-the-role-of-authors-and-contributors.html). The listing of authors will be as follows on the primary publication: MW Petersen will be first author, SN Myatra the second, and BKT Vijayaraghavan the third author. The next authors will be the site investigators according to the number of included participants per site, and then the other members of the Management Committee. A. Perner will be the last and corresponding author.

The Management Committee may grant additional authorships depending on personal input as per the Vancouver definitions. Investigators on sites may be granted authorship on sub-study publications if they contribute significantly as per the Vancouver definitions.

The IDMSC and investigators not qualifying for authorship will be acknowledged with their names under 'the COVID STEROID 2 trial investigators' in an *appendix* to the final manuscript.

The funding sources will be acknowledged, but they will have no influence on the data handling or analyses, the writing of the manuscript or the decision to publish.

#### *33.1 Sub-studies*

Sub-studies will be encouraged if they do not hamper the completion of the main protocol and can be conducted after approval of the specific protocol by the Management Committee and the authorities. Thus, specific protocols for any sub-studies will be submitted to and approved by the relevant authorities and ethic committees before the commencement of such studies. In Appendix 8 (18.8), any proposed sub-studies are listed.

#### *33.2 Intellectual property rights*

The COVID STEROID 2 trial group owns the trial data.

##### 33.3 Organisational framework

The COVID STEROID 2 trial will be conducted and managed by the Sponsor, Management Committee, (Appendix 1, 18.1), the dedicated trial site team, the investigators, and the Research Unit at Department of Intensive Care, Rigshospitalet.

### 34 Estimated trial timeline

- August 2020, authority approvals and 1st participant randomised

- December 2020, interim analysis

- Mid 2021, last participant randomised and primary report on 28-day outcomes submitted.

- Late 2021, report on 90-day outcomes submitted

- Mid 2022, report on 180-day outcomes submitted

18. Horby P, Lim WS, Emberson J, Mafham M, Bell J, Linsell L, et al. Effect of Dexamethasone in Hospitalized Patients with COVID-19: Preliminary Report. medRxiv. 2020:2020.06.22.20137273.
19. Ye Z, Wang Y, Colunga-Lozano LE, Prasad M, Tangamornsuksan W, Rochwerg B, et al. Efficacy and safety of corticosteroids in COVID-19 based on evidence for COVID-19, other coronavirus infections, influenza, community-acquired pneumonia and acute respiratory distress syndrome: a systematic review and meta-analysis. CMAJ. 2020.
20. Randomized, Embedded, Multifactorial Adaptive Platform Trial for Community-Acquired Pneumonia (REMAP-CAP) (NCT02735707). Available from: https://clinicaltrials.gov/ct2/show/NCT02735707?term=remap+cap&draw=2&rank=1.
21. Targeted Steroids for ARDS Due to COVID-19 Pneumonia: A Pilot Randomized Clinical Trial (NCT04360876). Available form: https://clinicaltrials.gov/ct2/show/NCT04360876?term=COVID+STEROID&draw=2&rank=3.
22. Steroid Dosing by bioMARker Guided Titration in Critically Ill Patients With Pneumonia (SMART) (NCT03852537). Available from: https://clinicaltrials.gov/ct2/show/NCT03852537?term=COVID+STEROID&draw=2&rank=4.
23. Glucocorticoid Therapy for COVID-19 Critically Ill Patients With Severe Acute Respiratory Failure (NCT04244591). Available from: https://clinicaltrials.gov/ct2/show/NCT04244591?term=COVID+STEROID&draw=2&rank=5.
24. Methylprednisolone in the Treatment of Patients With Signs of Severe Acute Respiratory Syndrome in Covid-19 (MetCOVID) (NCT04343729). Available from: https://clinicaltrials.gov/ct2/show/NCT04343729?term=COVID+methylprednisolone&draw=2&rank=3.
25. Efficacy of Dexamethasone Treatment for Patients With ARDS Caused by COVID-19 (DEXA-COVID19) (NCT04325061). Available from: https://clinicaltrials.gov/ct2/show/NCT04325061?term=COVID+and+dexa&draw=2&rank=1.
26. Community-Acquired Pneumonia : Evaluation of Corticosteroids (CAPE_COD) (NCT02517489). Available from: https://clinicaltrials.gov/ct2/show/NCT02517489?term=COVID+hydrocortisone&draw=2&rank=5.
27. Efficacy and Safety of Corticosteroids in COVID-19 (NCT04273321). Available from: https://clinicaltrials.gov/ct2/show/NCT04273321.
28. Dexamethasone Treatment for Severe Acute Respiratory Distress Syndrome Induced by COVID-19 (DHYSCO) (NCT04347980). Available from: https://clinicaltrials.gov/ct2/show/NCT04347980.
29. Dexamethasone for COVID-19 Related ARDS: a Multicenter, Randomized Clinical Trial (NCT04395105). Available from: https://clinicaltrials.gov/ct2/show/NCT04395105.
30. Dexamethasone and Oxygen Support Strategies in ICU Patients With Covid-19 Pneumonia (COVIDICUS) (NCT04344730). Available from: https://clinicaltrials.gov/ct2/show/NCT04344730.
31. Low-dose Hydrocortisone for Patients with COVID-19 and Severe Hypoxia (NCT04348305). Available from: https://clinicaltrials.gov/ct2/show/NCT04348305?term=COVID+STEROID&draw=2&rank=1. .
32. COVID-19-associated ARDS Treated With Dexamethasone: Alliance Covid-19 Brasil III (CoDEX) (NCT04327401). Available from: https://clinicaltrials.gov/ct2/show/NCT04327401.
33. Efficacy and Safety of Corticosteroids in Oxygen-dependent Patients With COVID-19 Pneumonia (CORTICOVIDHUGO) (NCT04359511). Available from: https://clinicaltrials.gov/ct2/show/NCT04359511.
34. Corticosteroids During Covid-19 Viral Pneumonia Related to SARS-Cov-2 Infection (CORTI-Covid) (NCT04344288). Available from: https://clinicaltrials.gov/ct2/show/NCT04344288

52. World Medical Association. WMA Declaration of Helsinki - Ethical Principles for Medical Research involving Human Subjects. Available from: https://www.wma.net/policies-post/wma-declaration-of-helsinki-ethical-principles-for-medical-research-involving-human-subjects/20132013/. Accessed on Feb 26, 2020. 2013.
53. European Medicines Agency. Science Medicines Health. Good Clinical Practice. Available from: https://www.ema.europa.eu/en/human-regulatory/research-development/compliance/good-clinical-practice#international-clinical-trials-section. Accessed on March 18 2020.
54. Chan AW, Tetzlaff JM, Altman DG, Laupacis A, Gotzsche PC, Krleza-Jeric K, et al. SPIRIT 2013 statement: defining standard protocol items for clinical trials. Ann Intern Med. 2013;158(3):200-7.
55. Corticosteroids. LiverTox: Clinical and Research Information on Drug-Induced Liver Injury. Bethesda (MD)2012.
56. ICH Steering Committee. International conference on harmonisation of technical requirements for registration of pharmaceuticals for human use. ICH Harmonised Tripartite. Guideline for Statistical Principles for Clinical Trials re.
57. O'Brien PC, Fleming TR. A multiple testing procedure for clinical trials. Biometrics. 1979;35(3):549-56.
58. Cao B, Wang Y, Wen D, Liu W, Wang J, Fan G, et al. A Trial of Lopinavir-Ritonavir in Adults Hospitalized with Severe Covid-19. N Engl J Med. 2020.
59. Kahan BC, Morris TP. Improper analysis of trials randomised using stratified blocks or minimisation. Stat Med. 2012;31(4):328-40.
60. Schulz KF, Altman DG, Moher D, Group C. CONSORT 2010 statement: updated guidelines for reporting parallel group randomised trials. BMJ. 2010;340:c332.
61. Singer M, Deutschman CS, Seymour CW, Shankar-Hari M, Annane D, Bauer M, et al. The Third International Consensus Definitions for Sepsis and Septic Shock (Sepsis-3). JAMA. 2016;315(8):801-10.
62. Myles PS, Williamson E, Oakley J, Forbes A. Ethical and scientific considerations for patient enrollment into concurrent clinical trials. Trials. 2014;15:470.
63. Laine C, Taichman DB, Mulrow C. Trustworthy clinical guidelines. Ann Intern Med. 2011;154(11):774-5.
64. Qaseem A, Forland F, Macbeth F, Ollenschlager G, Phillips S, van der Wees P, et al. Guidelines International Network: toward international standards for clinical practice guidelines. Ann Intern Med. 2012;156(7):525-31.
65. Guyatt GH, Oxman AD, Vist GE, Kunz R, Falck-Ytter Y, Alonso-Coello P, et al. GRADE: an emerging consensus on rating quality of evidence and strength of recommendations. BMJ. 2008;336(7650):924-6.
66. Burgess E, Singhal N, Amin H, McMillan DD, Devrome H. Consent for clinical research in the neonatal intensive care unit: a retrospective survey and a prospective study. Arch Dis Child Fetal Neonatal Ed. 2003;88(4):F280-5; discussion F5-6.
67. Faden RR, Beauchamp TL, Kass NE. Informed consent, comparative effectiveness, and learning health care. N Engl J Med. 2014;370(8):766-8.
68. Randolph AG. The unique challenges of enrolling patients into multiple clinical trials. Crit Care Med. 2009;37(1 Suppl):S107-11.
69. Cook D, McDonald E, Smith O, Zytaruk N, Heels-Ansdell D, Watpool I, et al. Co-enrollment of critically ill patients into multiple studies: patterns, predictors and consequences. Crit Care. 2013;17(1):R1.
70. Cook DJ, Ferguson ND, Hand L, Austin P, Zhou Q, Adhikari NK, et al. Coenrollment in a randomized trial of high-frequency oscillation: prevalence, patterns, predictors, and outcomes*. Crit Care Med. 2015;43(2):328-38.
71. Favas TT, Dev P, Chaurasia RN, Chakravarty K, Mishra R, Joshi D, et al. Neurological manifestations of COVID-19: a systematic review and meta-analysis of proportions. Neurol Sci. 2020;41(12):3437-70.

### 36 Appendices

#### 36.1 Appendix 1: Trial organisation diagram

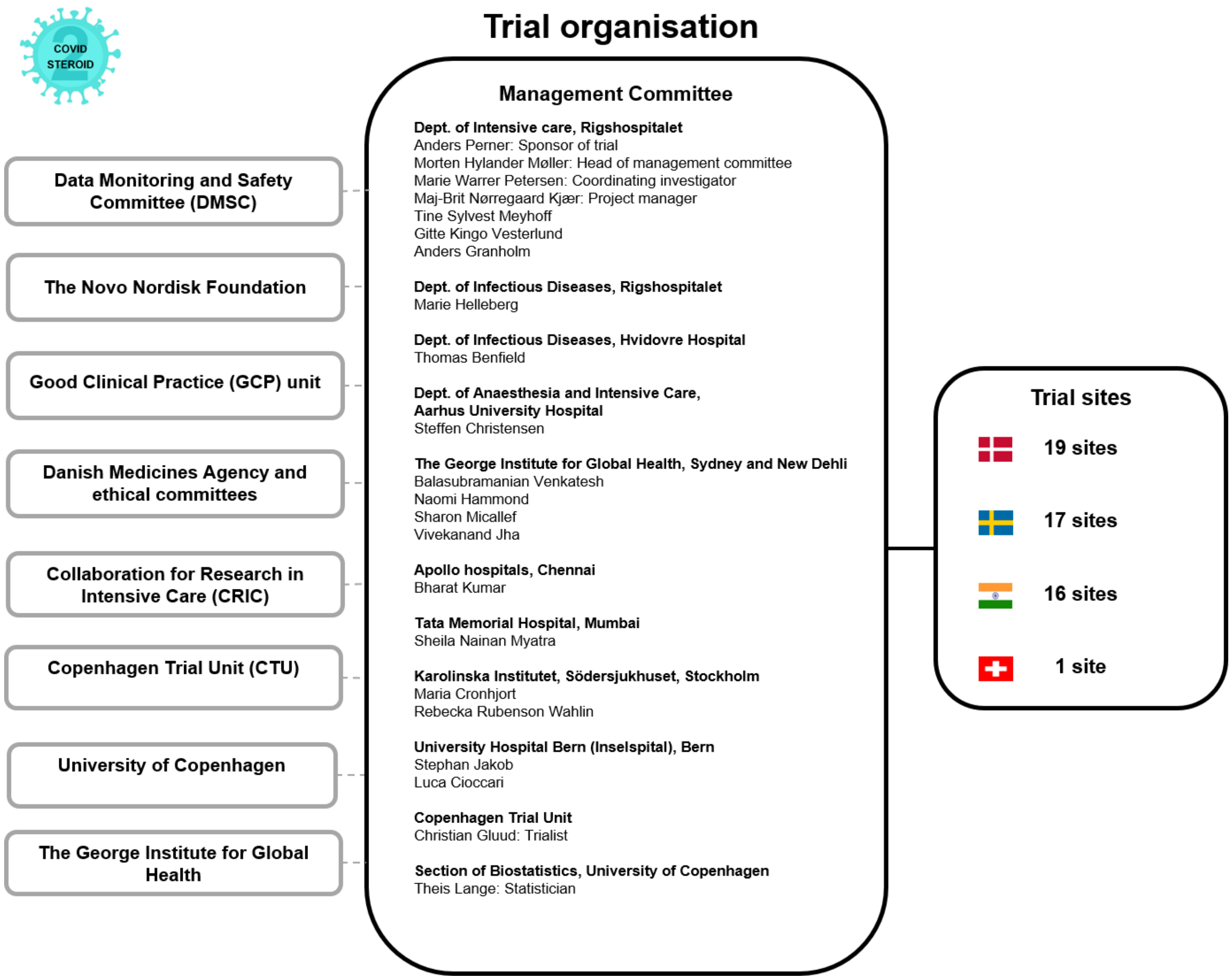

#### 36.2 *Appendix 2: Trial flow chart*

Please refer to the CONSORT Statement for more information (http://www.consort-statement.org/) (60). The flowchart will be modified to reflect the flow of participants in the trial. The flowchart (n= ) will be completed at the end of the trial.

**CONSORT 2010 Flow Diagram**

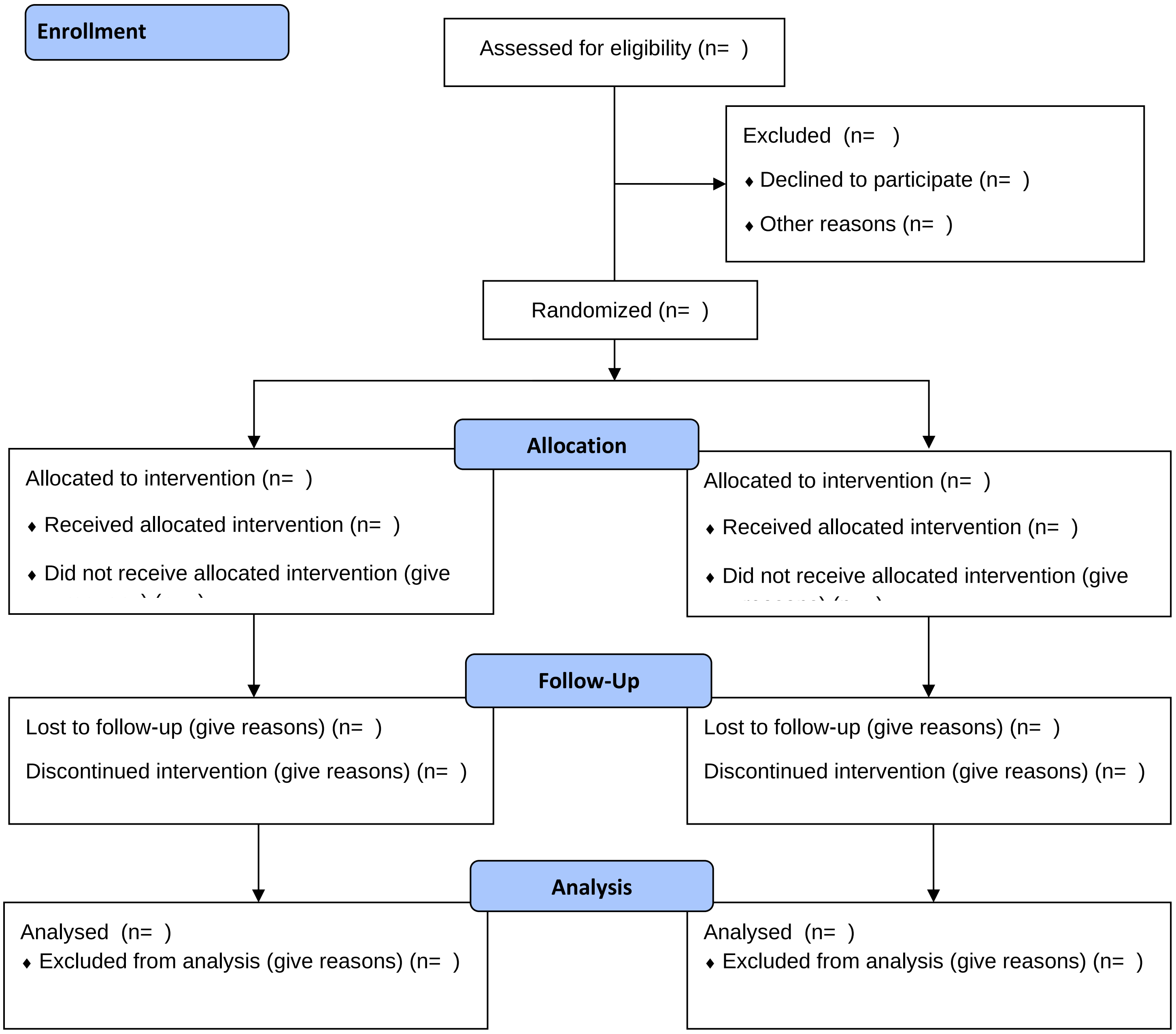

#### 36.3 *Appendix 3: Trial definitions*

**Definition of stratification variables**

Site: all participating trial sites (hospitals) will be assigned a number identifying the site.

Invasive mechanical ventilation: use of mechanical ventilation via a cuffed endotracheal tube at the time of randomisation.

Age: the age of the participant in whole years at the time of randomisation. Is the participant above 70 years old? (y/n). The participants will be stratified according to age ≥70 years versus <70 years.

**Participant identification**

National identification number (NIN): civil registration number (CPR number, 10 digits without dash) in or replacement CPR number if the participant does not have a CPR number in Denmark. Fictive NIN in other countries than Denmark generated from date of birth or year of birth and trial site ID.

**Definition of the inclusion criteria**

Age: defined under *Definition of stratification variables*

Confirmed SARS-CoV-2 requiring hospitalisation**:** We will include patients admitted to a trial site with SARS-CoV-2. We will accept any detections of SARS-CoV-2 approved by the national Health Authorities in the participating countries. Currently, detection of SARS-CoV-2 RNA from upper (i.e. pharyngeal swap) or lower airway secretions (i.e. tracheal secretion or bronchoalveolar lavage) is used.

Supplementary oxygen criterion at the time of randomisation:

- Invasive mechanical ventilation: Defined under *Definition of stratification variables* **OR**
- Non-invasive ventilation or continuous use of continuous positive airway pressure (CPAP) for hypoxia: Non-invasive ventilation includes positive pressure ventilation via a tight mask or helmet, continuous use of CPAP (mask, helmet or tracheostomy). This does not include intermittent use of CPAP.
- Oxygen supplementation with an oxygen flow ≥10 L/min irrespectively of system used (mask or nasal cannula) or the addition of atmospheric air

**Definition of the exclusion criteria**

Use of systemic corticosteroids in doses higher than 6 mg dexamethasone equivalents for other indications than COVID-19: systemic corticosteroids (IV, IM, oral or per GI tube; not including nebulised, inhaled or transdermal corticosteroids) in doses higher than 6 mg dexamethasone / 6 mg betamethasone / 200 mg cortisone / 160 mg hydrocortisone / 32 mg methylprednisolone / 40 mg prednisolone / 40 mg prednisone. Other indications include:

- Adrenal insufficiency (i.e. primary, secondary or tertiary)
- Anti-emetic treatment (i.e. post-operative or chemotherapy-induced nausea and vomiting)
- Immunosuppressive treatment (i.e. rheumatic diseases, allergic diseases, chronic obstructive pulmonary disease, haematological diseases, chronic kidney diseases, autoimmune hepatitis, inflammatory bowel disease, chronic neurological diseases)

Use of systemic corticosteroids for COVID-19 for 5 days or more: Use of systemic corticosteroids for COVID-19 for 5 consecutive days or more up to the day of screening.

Invasive fungal infection: Any of the following:

- Suspected invasive fungal infection: presence of plasma markers in blood (e.g. candida mannan antigen and galactomannan antigen)
- Confirmed invasive fungal infection: positive culture from blood, peritoneal fluid or tissue

Active tuberculosis: Either microbiologically confirmed or diagnosed based on epidemiological, clinical and radiographic data.

Pregnancy: confirmed by positive urine human gonadotropin (hCG) or plasma-hCG.

Known hypersensitivity to dexamethasone: history of any hypersensitivity reaction to dexamethasone, including but not limited to urticaria, eczema, angioedema, bronchospasm and anaphylaxis.

Consent not obtainable: patients where the clinician or investigator is unable to obtain the necessary consent according to the national regulations, including patients with no relatives or patients who are hospitalised against their will.

**Definition of baseline variables**

Sex: the genotypic sex of the participant

Age at enrolment: the age of the participant in whole years at the time of randomisation. The age will be calculated from the date of birth and date of enrolment in the COVID STEROID 2 trial.

Date of admission to hospital: the date of admission to the first hospital the participant was admitted to during the current hospital admission

Department at which participant was included:

- Emergency department: accident/emergency/casualty/acute department at COVID STEROID 2 trial site
- Hospital ward: medical or surgical ward at COVID STEROID 2trial site, including dedicated COVID-19 hospital wards
- Intermediate care unit: area of the hospital with higher resources to monitor patients as defined by the site, but invasive mechanical cannot be given.
- Intensive care unit: area of the hospital where invasive mechanical can be given.
- Other: any location in the same or another hospital not covered in the other categories

Use of respiratory support at randomisation:

- Closed system (y/n): Use of invasive mechanical ventilation as defined under *Definition of stratification variables*or use of Non-invasive ventilation or continuous use of continuous positive airway pressure (CPAP) for hypoxia as defined under *Definition of inclusion criteria.* If yes, latest $FiO_2$ prior to randomisation
- Open system with an oxygen flow ≥10 L/min: If yes, the maximum supplemental oxygen flow on an open system at randomisation (+/- 1 h) will be registered.

Limitations of care (y/n): participant with limitation(s) in use of life support (i.e. invasive mechanical ventilation, circulatory support, renal replacement therapy) and/or cardio-pulmonary resuscitation at the time of randomisation.

Chronic use of systemic corticosteroids for other indications than COVID-19 (y/n): Systemic corticosteroids (IV, IM, oral or per GI tube; not including nebulised, inhaled or transdermal corticosteroids) for any other indications than COVID-19 at the time of randomisation.

Treatment during current hospital admission prior to randomisation:

Agents with potential anti-viral action used against COVID-19: any treatment that potentially inhibits viral replication, categorised as remdesivir, convalescent plasma, or other (e.g. umifenovir, interferon alfa, interferon beta, camostat).

Anti-bacterial agents: any antibiotic treatment commenced due to documented or suspected bacterial infection before microbiological results are available
Agents with potential anti-inflammatory action: any treatment with potential anti-inflammatory actions used against COVID-19 prior to screening, categorised as Janus kinase inhibitor, IL-6 inhibitors or other.
Co-morbidities: any chronic co-morbidity present in the past medical history prior to admission and defined as follows:

- History of ischemic heart disease or heart failure: previous myocardial infarction, invasive intervention for coronary artery disease, stable or unstable angina, NYHA class 3 or 4 or any measured LVEF <40%.
- Diabetes mellitus: Treatment at time of hospital admission with any anti-diabetic medications.
- Chronic pulmonary disease: Treatment at time of hospital admission with any relevant drug indicating chronic pulmonary disease.
- Immunosuppressive therapy within the last 3-months: use of systemic immunosuppressive drugs (e.g. tumor necrosis factor (TNF) inhibitors, calcineurin inhibitors, mTOR inhibitors, anti-thymocyte globulins, interleukin-2 inhibitors, mycophenolate, azathioprine, belimumab, corticosteroids), chemotherapy (e.g. alkylating agents, anti-metabolites, mitotic inhibitors, topoisomerase inhibitors, others) or radiotherapy within the last 3 months before randomisation.

Blood values, interventions and vital parameters:

- Participant weight: measured or estimated in kg
- $PaO_2$, $SaO_2$ and lactate prior to inclusion: will be assessed from the most recent arterial blood gas sample; alternatively, if arterial blood gas sample is not available, $SpO_2$ will be assessed from the most recent measure by pulse oximeter.
- Circulatory support: infusion of any vasopressor/inotrope agent for a minimum of 1 hour (i.e. norepinephrine, epinephrine, phenylephrine, vasopressin analogues, angiotensin, dopamine, dobutamine, milrinone or levosimendan) within the last 24 hours prior to randomisation.
- Renal replacement therapy: any form of acute or chronic intermittent or continuous renal replacement therapy (including days between intermittent dialysis) within the last 72 hours prior to randomisation.

**Definition of variables assessed in day forms (day 1-14)**

- Invasive mechanical ventilation (on this day): defined under *Definition of the inclusion criteria.*

- Circulatory support (for at least 1 hour on this day): defined under *Definition of the baseline variables.*
- Any form of renal replacement therapy (on this day): any form of renal replacement therapy (e.g. dialysis, hemofiltration or hemodiafiltration) at any rate on this day. Including days between intermittent renal replacement therapy.

- SAR on this day (y/n for everyone)

- New episodes of septic shock: we will define septic shock according to the Sepsis-3 criteria (61):
    - Suspected or confirmed superinfection
    - New infusion (or 50% increase) of vasopressor/inotrope agent (*Definition in the baseline variables*) to maintain a mean arterial blood pressure of 65 mmHg or above
    - Lactate of 2 mmol/L or above in any plasma sample performed on the same day
- Invasive fungal infection: defined under *Definition of exclusion criteria*
- Clinically important gastrointestinal (GI) bleeding: any GI bleeding **AND** use of at least 2 unit of red blood cells on the same day. GI bleed defined as hematemesis, coffee ground emesis, melena, haematochezia or bloody nasogastric aspirate on this day.
- Anaphylactic reaction to IV dexamethasone: anaphylactic reactions defined as urticarial skin reaction **AND** at least one of the following observed after randomisation
    - Worsened circulation (>20% decrease in blood pressure or >20% increase in vasopressor dose)
    - Increased airway resistance (>20% increase in the peak pressure on the ventilation)
    - Clinical stridor or bronchospasm
    - Subsequent treatment with bronchodilators

**Definition of variables assessed in day forms (from day 1 and up to 10 days)**
- Use of open-label systemic corticosteroids on this day: Use of any open-label systemic (IV, IM or oral/per GI tube) corticosteroids (i.e. hydrocortisone, methylprednisolone, dexamethasone, prednisolone or prednisone) in any dose

- Trial intervention: Did the participant receive trial medication on this day: yes, if the trial participant received the bolus of trial medication on this day; no, if the trial participant did not receive the bolus of trial medication on this day.

  - If no, please apply reason for violating the protocol: By error/lack of resources, other reason.

**Definitions of outcome measures**

Primary outcome

Days alive without life support (i.e. invasive mechanical ventilation, circulatory support or renal replacement therapy) at day 28: will be assessed from the use of life support including invasive mechanical ventilation, vasopressor/inotrope, and renal replacement therapy as defined in *Definition of inclusion criteria*, *Definition of baseline variables* and *Definition of variables assessed in day form.* Total number of days alive without all 3 life supporting interventions within 28 days after randomisation.

Secondary outcomes

- Number of participants with one or more serious adverse reactions (SARs) at day 28: at least one new episode of either septic shock, invasive fungal infection, clinically important GI bleeding or anaphylactic reaction to IV dexamethasone as defined under *Definition of variables assessed in day form.*
- All-cause mortality at day 28 after randomisation: death from any cause within 28 days post-randomisation.
- All-cause mortality at day 90 after randomisation: death from any cause within 90 days post-randomisation.
- Days alive without life support (i.e. invasive mechanical ventilation, circulatory support or renal replacement therapy) at day 90: will be assessed from the use of life support invasive mechanical ventilation including vasopressor/inotrope, and renal replacement therapy as defined in *Definition of inclusion criteria*, *Definition of baseline variables* and *Definition of variables assessed in day form.* Total number of days alive without all 3 life supporting interventions within 28 days after randomisation.
- Days alive and out of hospital at day 90: will be assessed from the discharge date from the index hospitalisation, the number of days readmitted to hospital (if any) and date of death, if relevant, within the 90-day period

- All-cause mortality at 180 days after randomisation: death from any cause within 180 days post-randomisation.
- Health-Related Quality of Life (HRQoL) at 180 days after randomisation: HRQoL at 180 days (+/- 2 weeks): EQ-5D-5L and EQ-VAS scores (https://euroqol.org/) obtained by survey by mail or phone as chosen by the participant. Non-survivors will be given the worst possible score. If the participant is incapable of answering the questionnaire (e.g. due to cognitive impairment or coma) we will ask proxies to assess HRQoL for the trial participant (proxy point of view) using the questionnaire aimed for proxies. Non-survivors will be given the worst possible score. EQ-5D-5L will be converted to an index value in combination with the EQ-VAS quantitative measure (0-100 points) quantifying self-rated health

**Definitions of other variables assessed during follow up**

- Discharged against medical advice to home/other hospital/other facility (y/n)
  - If yes: was the participant on any life support (i.e. invasive mechanical ventilation, circulatory support or renal replacement therapy) at the time of discharge (y/n)?
    - If no, did the participant receive supplementary oxygen at the time of discharge?
      - Yes, 0-9 L/min of supplementary oxygen
      - Yes, ≥10 L/min of supplementary oxygen
      - No

- Use of ECMO from randomisation to day 28: oxygen supplied through extracorporeal membrane on any day from randomisation to day 28.

**Definitions of subgroups**

Elderly patients: ≥70 years versus <70 years. Age is defined under *Definition of stratification variables.*

Invasive mechanical ventilation: invasive mechanical ventilation versus oxygen by other delivery systems. Invasive mechanical ventilation is defined under *Definition of stratification variables*; oxygen by other delivery system encompass both non-invasive ventilation, continuous use of CPAP and oxygen supplementation with an oxygen flow ≥10 L/min irrespectively of system used or the addition of atmospheric air as defined under *Definition of inclusion criteria.*

Shock: patients with shock versus without shock. Shock of any cause in patients requiring infusion of vasopressor/inotropic agent (norepinephrine, epinephrine, phenylephrine, vasopressin analogues, angiotensin, dopamine, dobutamine, milrinone or levosemindan) to maintain a mean arterial blood pressure of 65 mmHg or above AND with a lactate of 2 mmol/L or above in any plasma within 24 hours of randomisation.

Duration of corticosteroid use before enrolment in COVID STEROID 2 trial: patients who received any systemic corticosteroid for COVID-19 for 0 to 2 consecutive days compared to 3 to 4 consecutive days up to enrolment.

Limitations of care: patients with limitations of care compared to patients without limitations of care. Limitation of care is defined under *Definition of baseline variables*.

Chronic use of systemic corticosteroids for other indications than COVID-19: patients with versus without chronic use of systemic corticosteroids as defined under *Definition of baseline variables*.

##### 36.4 Appendix 4: Trial medication labels for dexamethasone and betamethasone

**COVID STEROID trial medication for clinical trial**
Dexamethasone 12 mg OR dexamethasone 6 mg

For injection

Patient name.......................................................................................................................

Identification number.........................................................................................................

Date and time of preparation of trial medication....................................................

Signature.............................................................................................................................

Must be stored at ≤ 25 °C

Sponsor: Prof. Anders Perner, Dept. of Intensive Care, Rigshospitalet, Denmark. Tel. +45 3545 8333

**COVID STEROID trial medication for clinical trial**
Betamethasone 12 mg OR betamethasone 6 mg

For injection

Patient name.......................................................................................................................

Identification number.........................................................................................................

Date and time of preparation of trial medication....................................................

Signature.............................................................................................................................

Must be stored at ≤ 25 °C

Sponsor: Prof. Anders Perner, Dept. of Intensive Care, Rigshospitalet, Denmark. Tel. +45 3545 8333

#### 36.5 Appendix 5: Charter for the independent data monitoring and safety committee

**Introduction**

The independent Data Monitoring and Safety Committee (IDMSC) will constitute its own plan of monitoring and meetings. However, this charter defines the minimum of obligations and primary responsibilities of the IDMSC, its relationship with other trial components, its membership, and the purpose and timing of its meetings, as perceived by the COVID STEROID 2 Management Committee. The charter also outlines the procedures for ensuring confidentiality and proper communication, the statistical monitoring guidelines to be implemented by the IDMSC, and an outline of the content of the open and closed reports which will be provided to the IDMSC.

**Primary responsibilities of the IDMSC**

The IDMSC are responsible for safeguarding the interests of trial participants, assessing the safety and efficacy of the interventions during the trial, and for monitoring the overall conduct of the trial. The IDMSC will provide recommendations about stopping or continuing the trial to the Management Committee of the COVID STEROID 2 trial. The IDMSC may also – if applicable - formulate recommendations related to the selection/recruitment/retention of participants, their management, adherence to protocol-specified regimens and retention of participants, and the procedures for data management and quality control.

The IDMSC will be advisory to the COVID STEROID 2 Management Committee. The Management Committee will be responsible for promptly reviewing the IDMSC recommendations, to decide whether to continue or terminate the trial, and to determine whether amendments to the protocol or changes in trial conduct are required.

The IDMSC may meet physically or by phone at their own discretion in order to evaluate the planned interim analyses of the COVID STEROID 2 trial. The interim analysis will be performed by an independent statistician selected by the members of the IDMSC, Susanne Rosthøj from the Department of Biostatistics, University of Copenhagen. The IDMSC may additionally meet whenever they decide or contact each other by telephone or e-mail to discuss the safety for trial participants. The IDMSC can, at any time during the trial, request information about the distribution of events, including outcome measures and serious adverse reactions (SARs) according to group allocation. Further, the IDMSC can request unmasking of the interventions, if deemed important (see section on ‘closed sessions’). The recommendations of the IDMSC regarding stopping, continuing or changing the design of the trial should be communicated without delay to the COVID STEROID 2 Management Committee. As fast as possible, and no later than 48 hours, the Management Committee has the responsibility to inform all trial sites and investigators, about the recommendation of the IDMSC and the Management Committee decision hereof.

**Members of the IDMSC**

The IDMSC is an independent multidisciplinary group consisting of a clinician, a trialist and a biostatistician that, collectively, has experience in the conduct, monitoring and analysis of randomised clinical trials.

IDMSC Clinician

Christian Hassager, Professor in cardiology, Copenhagen University Hospital, Denmark

IDMSC Trialist

Manu Shankar-Hari, Clinician Scientist, Reader and Consultant in Intensive Care Medicine, National Institute for Health Research and Kings College, London, United Kingdom

IDMSC Biostatistician

Susanne Rosthøj, Department of Biostatistics, University of Copenhagen

**Conflicts of interest**

The members of the IDMSC will fill-in and sign a conflicts of interest form. IDMSC membership is restricted to individuals free of conflict of interest. The source of these conflicts may be financial, scientific, or regulatory in nature. Thus, neither trial investigators nor individuals employed by the sponsor, or individuals who might have regulatory responsibilities for the trial products, are members of the IDMSC. Furthermore. the IDMSC members do not own stocks in the companies having products being evaluated by the COVID STEROID 2 trial.

The IDMSC members will disclose to fellow members any consulting agreements or financial interests they have with the sponsor of the trial, with the contract research organisation (CRO) for the trial (if any), or with other sponsors having products that are being evaluated or having products that are competitive with those being evaluated in the trial. The IDMSC will be responsible for deciding whether these consulting agreements or financial interests materially impact their objectivity.

The IDMSC members will be responsible for advising fellow members of any changes in these consulting agreements and financial interests that occur during the trial. Any IDMSC members who develop significant conflicts of interest during the trial should resign from the IDMSC.

IDMSC membership is to be for the duration of the clinical trial. If any members leave the IDMSC during the trial, the Management Committee will appoint the replacement(s).

**Formal interim analysis meetings**

One formal interim analysis meeting will be held to review data related to protocol adherence, treatment efficacy and participant safety. The 3 members of the IDMSC will meet when 28-day follow-up data of 500 participants (50% of sample size) have been obtained.

**Final analysis meeting**

The 3 members of the IDMSC will meet when 28-day follow-up data the full sample size (1,000 participants) have been obtained.

**Proper communication**

To enhance the integrity and credibility of the trial, procedures will be implemented to ensure that the IDMSC has sole access to evolving information from the clinical trial regarding comparative results of efficacy and safety data aggregated by treatment group. An exception will be made to permit access to an independent statistician who will be responsible for serving as a liaison between the database and the IDMSC.

At the same time, procedures will be implemented to ensure that proper communication is achieved between the IDMSC and the Management Committee. To provide a forum for exchange of information among various parties who share responsibility for the successful conduct of the trial, a format for open sessions and closed sessions will be implemented. The intent of this format is to enable the IDMSC to preserve confidentiality of the comparative efficacy results while at the same time providing opportunities for interaction between the IDMSC and others who have valuable insights into trial-related issues.

**Closed sessions**

Sessions involving only IDMSC membership who generates the closed reports (called closed sessions) will be held to allow discussion of confidential data from the clinical trial, including information about protocol adherence and the relative efficacy and safety of interventions. To ensure that the IDMSC will be fully informed in its primary mission of safeguarding the interest of participants, the IDMSC will be blinded in its assessment of safety and efficacy data. However, the IDMSC can request unblinding from the Management Committee.

Closed reports will include analysis of the primary outcome measure and rates of SARs. These closed reports will be prepared by the independent IDMSC biostatistician, with assistance from the trial data manager, in a manner that allow them to remain blinded. The closed reports should provide information that is accurate, with follow-up on the primary outcome that is complete as soon as possible and at latest within one month from the date of the IDMSC meeting.

**Open reports**

For each IDMSC meeting, open reports will be available to all who attend the IDMSC meeting. The reports will include data on recruitment and baseline characteristics, and pooled data on eligibility violations, completeness of follow-up, and compliance. The independent IDMSC statistician will prepare these open reports in co-operation with the trial data manager. The reports should be provided to IDMSC members approximately three days prior to the date of the meeting.

**Minutes of the IDMSC Meetings**

The IDMSC will prepare minutes of their meetings. The closed minutes will describe the proceedings from all sessions of the IDMSC meeting, including the listing of recommendations by the committee. Because it is possible that these minutes may contain unblinded information, it is important that they are not made available to anyone outside the IDMSC.

**Recommendations to the Management Committee**

The planned interim analyses will be conducted after participant no. 500 have been followed for 28 days.

After the interim analysis meetings, the IDMSC will make a recommendation to the Management Committee to continue, hold or terminate the trial.

The independent IDMSC will recommend pausing or stopping the trial if group-differences in the primary outcome measure, SARs or suspected unexpected serious adverse reactions (SUSARs) are observed at the interim analysis with statistical significance levels adjusted according to the O'Brien-Fleming alfa-spending function (57). If the recommendation is to stop the trial, the IDMSC will discuss and recommend on whether the final decision to stop the trial will be made after the analysis of all participants included at the time (including participants randomised after this interim analysis) or whether a moratorium shall take place (setting the trial at hold) in the further inclusion of participants during these extra analyses. If further analyses of the participants included after the interim analysis is recommended, the rules for finally recommending stopping of the trial should obey the O'Brien-Fleming stopping boundary (57). Furthermore, the IDMSC can recommend pausing or stopping the trial if continued conduct of the trial clearly compromises participant safety. However, stopping for futility will not be an option as an intervention effects less than those estimated in the power calculation for the primary outcome may be clinically relevant as well.

All recommendation will be based on safety and efficacy considerations and will be guided by statistical monitoring guidelines defined in this charter and the trial protocol.

The Management Committee is jointly responsible with the IDMSC for safeguarding the interests of participants and for the conduct of the trial. Recommendations to amend the protocol or change the conduct of the trial made by the IDMSC will be considered and accepted or rejected by the Management Committee. The Management Committee will be responsible for deciding whether to continue, hold or stop the trial based on the IDMSC recommendations.

The IDMSC will be notified of all changes to the trial protocol or conduct. The IDMSC concurrence will be sought on all substantive recommendations or changes to the protocol or trial conduct prior to their implementation.

After completion of the interim analysis, the recommendations from the IDMSC and the conclusion reached by the Management Committee will be submitted to the Ethics Committee.

After completion of the full analysis of outcomes at day 28 (i.e. days alive without life support, mortality and SAR), the IDMSC will make a recommendation to the Management Committee to submit a primary report on 28-day outcomes or await the 90-day outcomes.

**Statistical monitoring guidelines**

The outcome parameters are defined in the statistical analysis plan in the COVID STEROID trial protocol. For the two intervention groups, the IDMSC will evaluate data on:

- Days alive without life support at day 28
- Mortality at day 28
- The number of participants with ≥1 SAR(s) and/or SUSAR(s) at day 28

The IDMSC will be provided a masked data set (as group 0 and 1) from the coordinating centre. The data set will include data on stratification variables and outcome measures according to the outcomes above in the two groups.

Based on evaluations of these outcomes, the IDMSC will decide if they want further data from the coordinating center and when to perform the next analysis of the data. For analyses, the data will be provided in one file as described below.

The IDMSC may also be asked to ensure that procedures are properly implemented to adjust trial sample size or duration of follow-up to restore power, if protocol specified event rates are inaccurate. If so, the algorithm for doing this should be clearly specified.

**Conditions for transfer of data from the Coordinating Centre to the IDMSC**

The IDMSC will be provided with a data file containing the data defined as follows:

Row 1 contains the names of the variables (to be defined below).

Row 2 to N (where N-1 is the number of participants having entered the trial) each contains the data of one participant.

Column 1 to p (where p is the number of variables to be defined below) each contains in row 1 the name of a variable and in the next N rows the values of this variable.

The values of the following variables should be included in the database for the all three interim analyses:

1. screening_id: a number that uniquely identifies the participant

2. rand_code: The randomisation code (group 0 or 1). The IDMSC is not to be informed on what intervention the groups received

3. days_alive_without_lifesup_d28_cum_indic (continuous scale)

4. day_28_indic: 28 day-mortality indicator (2 = censored, 1=dead, 0=alive at day 28)

5. SAR_indic: SAR indicator (1 = one or more SARs, 0 = no SAR)

#### 36.6 Appendix 6: Informed consent

Participants will be enrolled after consent by proxy is obtained according to Danish regulations. We will follow the normal procedures for collection of informed consent and any modifications to these as approved by the Ethics Committee due to restrictions in physical visits to the hospitals during the current SARS-CoV-2 epidemic. All consenting parties will be provided with written and oral information about the trial, so he/she is able to make an informed decision about participation in the trial.

All patients with COVID-19 and severe hypoxia will be temporarily incompetent because of the acute illness, low oxygen saturation and stress-response associated with lack of oxygen. Thus, participants will be enrolled after obtaining informed consent from a doctor (first trial guardian), who is independent of the trial, who has knowledge of the clinical condition and who is familiar with the trial protocol to such extent that he/she can judge for each patient, if it will be reasonable to enroll the patient in the trial.

As soon as possible after enrolment, consent will be obtained from the participant’s next of kin and a second trial guardian.

The second trial guardian is also a doctor who is independent of the trial, who has knowledge of the clinical condition and who is familiar with the trial protocol to such extent that he/she can judge for each patient, if it will be reasonable to enroll the patient in the trial.

To minimise the risk of transmission of SARS-CoV-2 between trial staff and the next of kin, we will inform and obtain informed consent from the next of kin by telephone. We will contact the next of kin by telephone and arrange a time and date for a telephone conversation with a member of the trial staff (e.g. doctor, research nurse, medical student etc) who is certified in obtaining informed consent. During this conversation, we will arrange how to send the written information to the next of kin (i.e. e-mail, post). We will encourage the next of kin to read the written information before the next conversation. We will also encourage the next of kin to bring a companion; in this case, the telephone conversation will be held with the telephone on speaker. After we have informed the next of kin about the trial, we will ask the next of kin to return the signed consent form by post.

Participants will be asked for informed consent as soon as possible after they regain consciousness. For participants, both oral and written information will be given preferably in person. The participant has the right to bring a companion.

If deemed necessary by the treating doctor, we will inform the participant orally before enrolment. In these instances, we will not include the patient, if he/she declines to participate. If the patient accepts to participate, we will re-inform the participant once he/she has regained full competence, i.e. when the participant receives less than 10 L/min of supplementary oxygen; is not mechanically ventilated; and is awake, alert and oriented as judged by treating clinician. First hereafter, we will collect the informed consent. For these participants, the procedure for obtaining informed consent will follow the same rules as stated above.

All consent forms will be signed by the consenting party and the member of trial staff who have provided trial information for the consenting party. We will emphasise that the consenting party has at least 24 hours to decide whether to give consent or not. Written information and the consent forms will be subjected to review and approval by the relevant ethic committees.

**Lack of informed consent from the participant's next of kin**

If information about the participant's next of kin is not available after inclusion, the investigator will seek information from e.g. the participant's general practitioner, the police, nursing homes etc. In these situations, it may take 1-2 weeks to conclude that no next of kin can be identified. If a next of kin is not identified and the participant remains incompetent, the trial intervention will be discontinued. All initiatives to identify the participant's next of kin will be documented in patient files, logs or similar.

**Lack of informed consent from the participant's next of kin and the participant deceases**

If the participant deceases before informed consent has been obtained (due to rapid progression of critical illness or because the participant's next of kin is not yet identified) and the participant has been correctly included in the trial, the collected data will be kept for analysis.

**Deviation from the standard informed consent**

According to the standard informed consent form from the National Ethics Committee regarding competent participants, the participant can choose not to receive information about the data collected during the trial. However, the purpose of this trial is not to generate new knowledge about the specific participant, so we find that this question is redundant, and have omitted the question from the consent form to spare the participant from making unnecessary decisions.

**Trial personnel**

Screening will be performed by the clinical staff. Collection of informed consent will be performed by the dedicated trial staff. If questions arise during informed consent, responsible trial staff can be reached through a 24-h hotline. All personnel with functions in the COVID STEROID 2 trial will be trained and approved according to GCP-guidelines before engaging in the trial.

#### 36.7 Appendix 7: Co-enrolment

Based upon an updated critical appraisal of the literature, the COVID STEROID 2 Management Committee endorses and encourages co-enrolment in the COVID STEROID 2 trial. The following issues have been considered.

**Ethical considerations**

Preventing eligible patients from co-enrolment in trials, which they would authentically value participating in, and whose material risks and benefits they understand, violates their autonomy - and thus contravenes a fundamental principle of research ethics (62).

Permitting co-enrolment is in accordance with existing recommendations for the conduct of trustworthy clinical practice guidelines, taking into account benefits and harms, quality of evidence, values and preferences (of patients or their relatives) and cost considerations, as outlined by the Institute of Medicine, the Guideline International Network, and according to The Grading of Recommendations Assessment, Development and Evaluation (GRADE) methodology (63-65).

Patient relatives have limited concerns about co-enrolment (66).

**General considerations**

Critically ill patients receive many different interventions in addition to the trial intervention because of acute and chronic illness. Consequently, the potential for interactions is a prerequisite in clinical trials in critically ill patients, and co-enrolment is thus little different from what occurs in single-enrolment trials (62).

In pragmatic trials, like the COVID STEROID 2 trial, other interventions will be given at random and are therefore difficult to control for. If interaction in fact is an issue, it may be better controlled for if patients are co-enrolled and randomised to more than one intervention.

Clinical research with a potential to inform and improve clinical practice is valuable and should be supported. More high-quality clinical research can be conducted in a timely fashion and more information can be generated to guide clinical practice, if co-enrolment is permitted (67).

**Scientific and statistical considerations**

Pragmatic clinical trials allowing inclusion of a broad range of trial participants and options for drug treatments and other therapies (co-enrolment) have higher external validity/generalizability than non-pragmatic trials with restrictions regarding trial participants and co-enrolment (68).

Non-pragmatic trials with restrictions regarding study participants and co-enrolment are exposed to drugs and other treatments in a less clinically relevant setting where interactions are largely uncontrolled and poorly evaluated. Co-enrolment in pragmatic trials facilitates evaluation of clinically relevant and patient-important interactions (62).

Co-enrolment into two or more trials does not invalidate the original randomization of the individual trials. Separate analysis of each individual trial, ignoring the issue of co-enrolment into the other trial, will retain the balance of patient characteristics expected by standard random assignment within each trial (62).

The National Institute of Health supports co-enrolment (68); so does the Canadian Critical Care Trials group (http://www.ccctg.ca/Home.aspx) and the Australian New Zealand Intensive Care Society's Clinical Trial Group (http://www.anzics.com.au/Pages/CTG/CTG-home.aspx). We have co-enrolment agreements with the two latter research groups.

Co-enrolment into two or more trials does not seem to affect the natural course of the disease of the other condition being studied (62). Co-enrolment does not appear to influence patient safety or trial results (69, 70). Empirically, co-enrolment has a small effect on study power (62).

In conclusion, we highly support and encourage co-enrolment because of overall benefit, including ethical, practical and scientific benefit, and no evidence of harm.

**Co-enrolment agreement form**

We will encourage engagement in research projects other than the COVID STEROID 2 trial. Please fill in the information of the trial to be evaluated as counterpart for co-enrolment with the COVID STEROID 2 trial and send it by e-mail to.

Once we have received the information below, we will contact the principal/coordinating investigator of the trial and facilitate exchange of protocols and other relevant documents between the Management Committees. You will find a list of titles already considered for co-enrolment by clicking http://www.cric.nu/covid-steroid-2-co-enrolment-list/

We have prepared the form for only one trial, but please feel free to copy as many forms as you need. The co-enrolment agreement form can be found by clicking http://www.cric.nu/covid-steroid-2-co-enrolment-form/

**Official full/short title of the project:**

**Contact information of principal/coordinating investigator of the trial:**

Name:

E-mail:

#### *36.8 Appendix 8: List of proposed sub-studies*

A Bayesian secondary analysis of all outcomes recorded within 90 days of randomisation.

#### *36.9 Local trade names for dexamethasone/betamethasone used in the COVID STEROID 2 trial*

**Denmark**

Dexavit™, Vital Pharma Nordic, Denmark, ATC code: H02AB02.

**Sweden**

Betapred™, Alfasigma S.p.A., Italy, ATC code: H02AB01.

**Switzerland**

Mephameson™, Mepha Pharma AG, Switzerland, ATC code: H02AB02.

**India**

Daksone™, Daksh Pharmaceuticals Pvt. Ltd, India, ATC code: H02AB02.

Dacdac™, Wockhardt Limited (Merind), India, ATC code: H02AB02.

Decmax™, GLS Pharma Ltd., India, ATC code: H02AB02.

Demisone™, Cadila Pharmaceuticals Ltd. (Genvista), India, ATC code: H02AB02.

Dexona™, Zydus Cadila Healthcare Ltd. (Alidac), India, ATC code: H02AB02.

Dex-V™, Vensat Bio, India, ATC code: H02AB02.

Intradex™, Intra Labs India Pvt. Ltd, India, ATC code: H02AB02.

#### *36.10 List of serious adverse events that do not have to be reported to the sponsor within 24 hours of occurrence*

Serious adverse events seen frequently in critically ill patients with COVID-19 and/or critically ill patients in general are listed in the table below. The listed serious adverse events do not have to be reported to the sponsor within 24 hours of occurrence if adjudicated not to be related to the intervention and expected in the patent population.

| **Organ system** | **Serious adverse events** |
|---|---|
| Nervous system | Stroke (71)<br>Psychosis (72)<br>Delirium (73)<br>Seizures (72)<br>Coma (72) |
| Respiratory system | Pneumothorax (74)<br>Pneumonia (75)<br>Acute respiratory failure (74) |
| Circulatory system | Severe heart failure, including cardiogenic shock (76)<br>Cardiac arrest (76)<br>Acute coronary syndrome (76)<br>Sepsis (75, 77)<br>Pulmonary embolus (78) |
| Digestive system | Pancreatitis (79)<br>Acute hepatic injury (80)<br>Ileus (81)<br>Bowel ischemia (81) |
| Urinary system | Acute kidney injury (74, 82)<br>Urinary tract infection (75) |
| Skeletal and muscular system | Soft tissue infection (75)<br>Osteomyelitis (75) |
| Skin and soft tissue | Decubitus (83) |
| Endocrine system | Diabetic ketoacidosis and/or coma (84) |
| Blood and lymphatic system | Clinically important thrombosis (78) |

| | |
|---|---|
| | Clinically important bleeding* (85) |

*Associated with transfusion of at least 2 units of red blood cells. If bleeding from the gastrointestinal tract, report as serious adverse reaction in the eCRF within 24 hours of occurrence.

#### Summary of changes to the original protocol

**Protocol version 1.7, 17 August 2020**

Original approved protocol entitled 'Higher vs. Lower Doses of Dexamethasone in Patients with COVID-19 and Severe Hypoxia: the COVID STEROID 2 trial',

**Protocol version 1.8, 07 December 2020 (amendment 1)**

<u>Section 2.2. Local investigators and clinical trial sites</u>

Description: New trial site in Denmark

**Protocol version 1.8, 09 January 2020 (other changes)**

<u>Section 12.2. Statistical methods, Section 18.3. Appendix 3: Trial definitions.</u>

Description: Removal of subgroup analysis of patients with compared to without shock. Addition of subgroup analyses of patients who receive IL-6 inhibitors at baseline compared to patients who do not receive IL-6 inhibitors at baseline and patients enrolled in India compared to patients enrolled in Denmark, Sweden and Switzerland.

**Protocol version 1.9, 27 January 2021 (amendment 2)**

<u>Section 2.2. Local investigators and clinical trial sites</u>

Description: New primary investigator at trial site in Denmark

<u>Section 10.3. Assessment of adverse events, Section 18.10. Appendix 10. List of serious adverse events that do not have to be reported to the sponsor within 24 hours of occurrence</u>

Description: New procedure for reporting serious adverse events (SAEs). SAEs seen frequently in critically ill patients with COVID-19 and/or critically ill patients in general do not have to be reported to the sponsor within 24 hours of occurrence if adjudicated not to be related to the intervention and expected in the patent population. The procedure was approved by relevant Danish authorities.

### Statistical analysis plan as of 25 January 2021

#### Dexamethasone 12 mg versus 6 mg for patients with COVID-19 and severe hypoxia: An international, randomized, blinded trial

The protocol and statistical analysis plan were published before conducting the interim analysis.[1] Analysis of data for the primary publication will be conducted according to this statistical analysis plan except for the changes outlined below.

**Coordinating investigator**

Marie Warrer Munch, Department of Intensive Care, Copenhagen University Hospital, Copenhagen, Denmark.

**Trial statistician**

Theis Lange, Section of Biostatistics, University of Copenhagen, Copenhagen, Denmark.

**Trial sponsor**

Anders Perner, Department of Intensive Care, Copenhagen University Hospital, Copenhagen, Denmark.

**Applicable protocol registration numbers**

ClinicalTrials.gov identifier NCT04509973

Ethics committee number H-20051056

EudraCT number 2020-003363-25

Danish Medicines Agency number 2020-07-16

Swiss National Clinical Trials Portal: SNCTP000004116

CTRI number: 2020/10/028731

### 1 METHODS

#### 1.1 TRIAL DESIGN

The COVID STEROID 2 trial is an investigator-initiated, international, parallel group, blinded, centrally randomised and stratified clinical trial. We plan to enrol 1,000 hospitalised adult patients with COVID-19 and severe hypoxia from sites in Denmark, Sweden, Switzerland and India. All trial results will be reported in accordance with the Consolidated Standards of Reporting Trials (CONSORT) statement.[2]

#### 1.2 TRIAL CONDUCT

We have prepared the COVID STEROID 2 protocol in accordance with the Standard Protocol Items: Recommendations for Interventional Trials (SPIRIT) Guidelines.[3] The trial will be conducted in accordance with the trial protocol, the Helsinki Declaration (latest version),[4] the International Conference on Harmonization (ICH) Good Clinical Practice (GCP) guidelines (latest version)[5] and all applicable laws in the participating countries.

#### 1.3 RANDOMISATION

Participants will be randomised in a 1:1 ratio to 12 mg vs. 6 mg of dexamethasone using a central web-based randomisation system administered by the Copenhagen Trial Unit (CTU). The randomisation will be performed using computer-generated allocation sequence lists stratified by trial site, invasive mechanical ventilation (yes/no) and age below 70 years (yes/no) with varying block sizes.

#### 1.4 ALLOCATION CONCEALMENT

The allocation sequence will be unknown to the trial staff preparing the trial medication, the clinicians, the investigators and the statistician conducting the analyses. The group allocations will remain masked (coded as 0 and 1) until two versions of the abstract for the trial report have been written.

#### 1.5 BLINDING

The allocation will be masked for all participants, clinical staff, trial staff reporting outcome data, the COVID STEROID 2 Management Committee, and the trial statistician.

#### 1.6 INCLUSION CRITERIA

We will screen for enrolment of adults with COVID-19 and severe hypoxia fulfilling the following three inclusion criteria:

- Aged 18 years or above **AND**
- Confirmed SARS-CoV-2 requiring hospitalisation **AND**
- Use of one of the following:
    - Invasive mechanical ventilation **OR**
    - Non-invasive ventilation or continuous use of continuous positive airway pressure (CPAP) for hypoxia **OR**
    - Oxygen supplementation with an oxygen flow of at least 10 L/min independent of delivery system.

A detailed description of the inclusion criteria is provided in the protocol.

#### 1.7 EXCLUSION CRITERIA

Any patient fulfilling one or more of the following exclusion criteria at the time of screening will be excluded from the trial:

- Use of systemic corticosteroids for other indications than COVID-19 in doses higher than 6 mg dexamethasone equivalents.
- Use of systemic corticosteroids for COVID-19 for 5 consecutive days or more
- Invasive fungal infection.
- Active tuberculosis.
- Fertile woman (<60 years of age) with positive urine human gonadotropin (hCG) or plasma-hCG.
- Known hypersensitivity to dexamethasone.
- Previously randomised in the COVID STEROID 2 trial.
- Informed consent not obtainable.

We will allow co-enrolment with other clinical trials unless the interventions or protocols of the trials collide. A detailed description of the exclusion criteria is provided in the protocol.

#### 1.8 OUTCOMES

Detailed definitions of all outcome measures are provided in the protocol.

##### 1.8.1 Primary outcome

Days alive without life support (i.e. invasive mechanical ventilation, circulatory support or renal replacement therapy (including days in between intermittent renal replacement therapy)) from randomisation to day 28.

##### 1.8.2 Secondary outcomes

- Number of participants with one or more serious adverse reactions (SARs) to dexamethasone from randomisation to day 28 defined as new episodes of septic shock, invasive fungal infection, clinically important gastrointestinal (GI) bleeding or anaphylactic reaction to IV dexamethasone.
- All-cause mortality at day 28.
- All-cause mortality at day 90.
- Days alive without life support at day 90.
- Days alive and out of hospital at day 90.
- All-cause mortality at day 180.
- HRQoL at day 180 using EQ-5D-5L and EQ-VAS.[6]

#### 1.9 REGISTERED VARIABLES

Detailed definitions of the registered variables are provided in protocol. Data will be entered in an online electronic case report form (OpenClinica).

##### 1.9.1 Baseline variables

1. Sex.
2. Age at enrolment (date of birth).
3. Date of admission to hospital.
4. Number of days with symptoms of COVID-19 before hospital admission.
5. Type of department at which the participant was included (i.e. emergency department, hospital ward, intermediate care unit, intensive care unit).
6. Use of respiratory support at randomisation:
    - Closed system: invasive mechanical ventilation or non-invasive ventilation or continuous use of CPAP (including latest fraction of inspired oxygen ($FiO_2$) and number of days on closed system ventilation prior to randomisation).

- Open system ventilation with an oxygen flow ≥10 L/min (including maximum supplemental oxygen flow at randomisation +/- 1 hour).

7. Limitations of care (i.e. limitations for invasive mechanical ventilation, circulatory support, renal replacement therapy, or cardio-pulmonary resuscitation) at the time of randomisation.
8. Chronic use of systemic corticosteroids for other indications than COVID-19.
9. Treatment for COVID-19 during current hospital admission prior to randomisation:
   - Agents with potential anti-viral action (i.e. remdesivir, convalescent plasma, other).
   - Agents with potential anti-inflammatory action (i.e. Janus kinase inhibitor, interleukin-6 inhibitors, other).
10. Treatment with systemic antibacterial agents in the 24 hours prior to randomisation.
11. Chronic co-morbidities:
    - History of ischaemic heart disease or heart failure.
    - Treatment at the time of hospital admission with any anti-diabetic drug indicating diabetes mellitus.
    - Treatment at the time of hospital admission with any drug indicating chronic pulmonary disease.
    - Use of immunosuppressive therapy within the last 3-months.
12. Laboratory values, interventions and vital parameters:
    - Participant weight (kilograms).
    - Arterial partial pressure of oxygen ($PaO_2$).
    - Saturation of oxygen ($SaO_2$) from arterial blood gas sample (preferred) or pulse oximeter.
    - Use of circulatory support within the last 24 hours prior to randomisation.
    - Use of any form of renal replacement therapy within the last 72 hours prior to randomisation.
    - Highest plasma lactate within the last 24 hours prior to randomisation.

##### 1.9.2 Variables registered daily during admission for the first 14 days after randomisation (day forms)

1. Use of invasive mechanical ventilation.
2. Use of circulatory support (continuous infusion of vasopressor/inotrope for a minimum of 1 hour).

3. Use of any form of renal replacement therapy (including days between intermittent renal replacement therapy).
4. SAR(s)
    - New episodes of septic shock.
    - Invasive fungal infection.
    - Clinically important GI bleeding.
    - Anaphylactic reaction to IV dexamethasone (only recorded during the intervention period)

##### 1.9.3 Protocol violations during the intervention period

Protocol violations will be recorded during the intervention period (for up to 10 days).

1. Use of open-label systemic corticosteroids.
2. Trial medication not administered as per protocol (1 bolus injection of either 12 mg or 6 mg dexamethasone according to the allocation on each day during the intervention period).

##### 1.9.4 Follow-up 28 days after randomisation

1. Vital status (if dead, date of death).
2. Number of days on invasive mechanical ventilation from day 15-28.
3. Number of days with circulatory support (continuous infusion of vasopressor/inotrope for a minimum of 1 hour) from day 15-28.
5. Number of days on renal replacement therapy (including days between intermittent renal replacement therapy) from day 15-28.
4. The occurrence of SAR(s) (section 2.10.2) from day 15-28 (if yes, apply date(s)).
5. Use of extracorporeal membrane oxygenation from randomisation to day 28 (y/n)
6. Discharged against medical advice to home/other hospital/other facility, including degree of life support (i.e. mechanical ventilation, circulatory support, renal replacement therapy) or supplementary oxygen at the time of discharge.

We will not register the occurrence of suspected unexpected serious adverse reactions (SUSARs) in dayforms or at 28-day follow-up as these rarely occur. Instead, SUSARs will be reported directly and without delay by the site investigator to the sponsor.

##### 1.9.5 Follow-up 90 days after randomisation

1. Vital status (if dead, date of death).
2. Number of days on invasive mechanical ventilation from day 29-90.

3. Number of days with circulatory support (continuous infusion of vasopressor/inotrope for a minimum of 1 hour) from day 29-90.
4. Number of days on renal replacement therapy (including days between intermittent renal replacement therapy) from day 29-90.
5. Date of discharge from hospital.
6. Additional hospital admissions (date(s) of re-admission(s) and discharge(s)).

###### 1.9.6 Follow-up 180 days after randomisation

1. Vital status (if dead, date of death).
2. HRQoL assessed by EQ-5D-5L.[6]
3. HRQoL assessed by EQ-VAS.[6]

#### 1.10 GENERAL ANALYTIC PRINCIPLES

We will conduct the primary analyses of both primary and secondary outcomes in the intention-to-treat (ITT) population (i.e. all randomised participants for whom consent has been given to use data). For the primary outcome, this analysis will be supplemented with a sensitivity analysis in the per protocol (PP) population (i.e. the ITT population except those having one or more major protocol violations as defined in section 2.10.3). All statistical tests will be two-tailed and reported with confidence intervals (CIs).

Significance level is set to 5% including the interim analysis. In practice, this means that a p-value below 0.0492 will be considered significant if the trial is not stopped at the interim analysis and accordingly 95.08% confidence intervals will be employed. For all secondary outcomes, we will employ 99% CIs for (p-value threshold for significance 0.01) due to the multiplicity of these. If not stopped early, the mortality outcomes will be tested in a hierarchical procedure along with the primary outcome (first primary outcome, then 28 days mortality, and finally 90 days mortality) reusing the alpha if the previous test was significant. If the primary outcome is insignificant at trial conclusion, 0.01 significance thresholds will be employed for all additional outcomes, but the results interpreted with caution.

#### 1.11 MISSING DATA

We will perform complete case analyses if less than 5% of patients have missing data for variables included in the primary or secondary outcome analyses. If 5% or more of patients have missing data for the outcome/covariates in any analysis, we will use multiple imputation with chained equations for that analysis. If multiple imputation is used, we will use the predictive mean matching and logistic regression methods for numerical and categorical variables, respectively, with 25 datasets imputed separately in each treatment group.[7,8] We will include all stratification variables, all variables used in the applicable analyses, important baseline prognostic variables (age, all co-morbidities listed above, use of all three life support measures at baseline), and all outcomes available at the time of analysis in the imputation models. If multiple imputation is used, these results will be reported as the primary and supplemented with complete case analyses and best-worst/worst-best analyses (as previously described[9]).

#### 1.12 STATISTICAL ANALYSES

##### 1.12.1 Primary outcome

Primary analysis of the primary outcome

1. Kryger Jensen and Lange test adjusted for stratification variables (site, invasive mechanical ventilation, and age below 70 years) in the ITT population.[10]

Sensitivity analysis of the primary outcome

2. Kryger Jensen and Lange test adjusted for stratification variables (site, invasive mechanical ventilation, and age below 70 years) and additional important prognostic baseline risk factors, i.e. all co-morbidities listed above, and use of circulatory support or renal replacement therapy in the ITT population.[10]
3. Kryger Jensen and Lange test adjusted for stratification variables (site, invasive mechanical ventilation, and age below 70 years) in the PP population.[10]
4. If 5% or more of patients have missing data and multiple imputation is used: best-worst/worst-best case analyses and complete case analysis.[8]

Subgroup analyses of the primary outcome

5. Test of interaction between the intervention and the pre-planned subgroups (section 2.13.4) by Kryger Jensen and Lange test adjusted for stratification variables (site, invasive mechanical ventilation, and age below 70 years) in the ITT population.

The Kryger Jensen and Lange test is a joint test for no treatment effect on an outcome which can have probability point mass in a single value (i.e. zero days alive without life support within 28 days).[10] The test builds on combining two regressions; we can therefore adjust as per usual analysis despite the expectation that the outcome will be highly skewed.[10]

Results from all analyses of the primary outcome will be reported as adjusted mean differences and median differences with confidence intervals (see preceding section for details on significance level). Secondarily, we will report the unadjusted (crude) mean differences and median differences.

As the primary outcome is composite, we will report results from the analysis of each component in a supplement to the main report.

##### 1.12.2 Secondary outcomes

<u>Binary outcomes</u>

We will conduct the following analyses for all binary outcomes (i.e. number of participants with one or more SARs at day 28; all-cause mortality at day 28, 90 and 180):

*Primary analysis*

1. Generalised linear models with log links and binomial error distributions adjusted for the stratification variables in the ITT population.[11]

*Secondary analysis*

2. Fisher's exact test in the ITT population.
3. Kaplan-Meier survival curve for the crude data on all-cause mortality at day 28, 90 and 180.

Results will be reported as adjusted relative risks and secondarily adjusted risk differences with corresponding confidence intervals (see section 2.11 for details on significance level). Secondarily, we will report the unadjusted (crude) relative risks and absolute risk differences.

We will conduct the following analyses for all continuous outcomes (i.e. days alive without life support at day 90; days alive and out of hospital at day 90; HRQoL at day 180 using EQ-5D-5L and EQ-VAS):

1. Kryger Jensen and Lange test adjusted for the stratification variables in the ITT population.[10]

The results will be reported as adjusted mean differences and median differences with confidence intervals (see preceding section for details on significance level) and unadjusted (crude) mean differences and median differences.

For composite outcomes (i.e. number of participants with one or more SARs at day 28; days alive without life support at day 90), we will report results from the analysis of each component in a supplement to the main report.

##### 1.12.3 Power estimations

Sample size and power estimations for the primary outcome

At maximum, we will randomise 1,000 participants. A blinded statistician will conduct an interim analysis after the first 500 participants have been followed for 28 days. The alpha values for the interim analysis and the final analysis are 0.0054 and 0.0492, respectively as by the O'Brien-Fleming bounds, which preserves type I error at the usual 5%. The trial has 85% power to detect a 15% relative reduction in 28-day mortality combined with a 10% reduction in time on life support among the survivors.

Power estimations for the secondary outcomes

We have 80% statistical power to detect the following effects for the secondary outcomes:

- A 21% relative risk reduction for the mortality at day 28 (control event rate 30%)
- A 18% relative risk reduction for the mortality at day 90 (control event rate 40%)
- A 32% relative risk reduction for the number of participants with one or more SARs (control event rate 15%)
- A 15% relative risk reduction for the mortality at day 180 (control event rate 50%)

The estimates of control event rates for mortality at day 28 originate in data of previous COVID-19 studies;[12,13] the estimates of the control event rates for mortality at day 90 and the number of participants with SARs are based on our best clinical estimate. We expect the outcomes ‘days alive out of hospital at day 90’ and ‘HRQoL at 180 days’ to be highly skewed (non-normally distributed). The power estimations for these outcomes would be uncertain, and we therefore refrain from making these estimates.

##### 1.12.4 Pre-planned subgroup analyses

We will assess the heterogeneity of the intervention effects on the primary outcome in the following subgroups based on baseline characteristics:

- Patients ≥70 years compared to <70 years of age: hypothesised larger beneficial effect of higher dose dexamethasone in patients <70 years of age.
- Patients who receive invasive mechanical ventilation compared to oxygen by other delivery systems: hypothesised larger beneficial effect of higher dose dexamethasone in patients who receive oxygen by other delivery systems.
- Patients who received corticosteroids for COVID-19 for 0 to 2 days compared to 3 to 4 days before enrolment: hypothesised larger beneficial effect of higher dose dexamethasone in patients with short duration (≤2 days) of corticosteroid use before enrolment.
- Patients who receive IL-6 inhibitors at baseline compared to patients who do not receive IL-6 inhibitors at baseline: hypothesised larger beneficial effect of higher dose dexamethasone in patients who receive IL-6 inhibitors at baseline.
- Patients with limitations of care (i.e. not for invasive mechanical ventilation, circulatory support, renal replacement therapy, cardio-pulmonary resuscitation) compared to patients without limitations of care: hypothesised larger beneficial effect of higher dose dexamethasone in patients without limitations of care.
- Patients with compared to without chronic use of systemic corticosteroids: hypothesised larger beneficial effect of higher dose dexamethasone in patients without chronic use of systemic corticosteroids.
- Patients enrolled in India compared to patients enrolled in Denmark, Sweden and Switzerland: hypothesised different effect of higher dose dexamethasone in patients enrolled in India as compared to patients enrolled in Denmark, Sweden and Switzerland.

Detailed definitions of the subgroups are available in the protocol.

#### 1.13 Independent Data Monitoring and Safety Committee

We have formed an Independent Data Monitoring and Safety Committee (IDMSC) consisting of a multidisciplinary group of a clinician, a trialist and a biostatistician that, collectively, have experience in the conduct, monitoring and analysis of randomised clinical trials. The charter for the IDMSC is provided in the protocol.

#### 1.14 Interim analysis

The independent statistician of the IDMSC will conduct one blinded interim analysis after 500 participants (50%) have been followed for 28 days. The alpha value for the interim analysis is 0.0054 as by the O'Brien-Fleming bounds, which preserves type I error at the usual 5%.[14] The trial will be stopped early if the alpha cut-off is crossed at the interim analysis.

The IDMSC will be provided with the following outcome data with the two groups masked (e.g. interventions coded as 0 and 1):

- Days alive without life support (i.e. invasive mechanical ventilation, circulatory support or renal replacement therapy (including days in between intermittent renal replacement therapy)) from randomisation to day 28.
- Number of participants with one or more SARs or SUSARs from randomisation to day 28.

The data set will also include data on the stratification (i.e. site, invasive mechanical ventilation, and age below 70 years) and baseline variables.

Statistical analyses conducted by the IDMSC

1. Days alive without life support at day 28: Kryger Jensen and Lange test adjusted for stratification variables (site, invasive mechanical ventilation, and age below 70 years) in the ITT population.[10]
2. Number of participants with one or more SARs or SUSARs from randomisation to day 28: generalised linear model with log links and binomial error distributions adjusted for the stratification variables (site, invasive mechanical ventilation, and age below 70 years) in the ITT population.[11]

The IDMSC can request additional data from the coordinating centre or unblinding of the intervention groups during the whole course of the trial.The IDMSC will submit their recommendations to the trial Management Committee, which makes the final decision to continue, pause or stop the trial.

#### 1.15 MONITORING DURING THE STUDY

The trial will be externally monitored according to the GCP Directive. A monitoring and data verification plan has been developed together with the GCP unit at Rigshospitalet, University of Copenhagen.

The trial will also be centrally monitored by the Sponsor or his delegates through the electronic case report form (eCRF), including monitoring of protocol adherence.

#### 1.16 CLOSE OUT

We will ensure that a plan for long-term storage of data and source documentation has been made at each site upon completion of the trial.

#### 1.17 DATA SHARING STATEMENT

We will make the final de-identified data set available for sharing in accordance with the recent International Committee of Medical Journal Editors (ICMJE) recommendations[15] and data sharing agreements adhering to the laws of the participating countries. All trial-related documents are available from www.cric.nu/covid-steroid-2.

#### Summary of changes to the statistical analysis plan made after 25 January 2021

Section 1.12.1 Primary outcome

We performed a *post hoc* analysis of bootstrapped adjusted mean difference because the data were markedly skewed (a high proportion (41.4%) of day 28 counts).

Before the lock of the trial database, we decided to do a post hoc sensitivity analysis assigning patients who died within 28 days the worst possible outcome (i.e. 0 days alive without life support) because this has previously been used in similar trials.[2,3]

Section 1.12.2 Secondary outcomes

It was stated that a generalized linear model with log and identity links should be used to compute risk ratios and risk differences, respectively, with confidence intervals for the analyses of the secondary outcomes. However, when applying the model to the trial data, convergence of the model was not obtained for the adjusted analyses. Accordingly, a two-step procedure was employed instead where a logistic regression model adjusted for stratification variables and g-computation to convert the output from the logistic regression to either a risk ratio or a risk difference. By construction, these quantities are adjusted for any chance differences in baseline values of stratification variables. For the unadjusted analyses of the secondary outcomes, the planned analysis was conducted without convergence problems.
