## Supplementary material for "Dexamethasone 12 mg versus 6 mg for patients with COVID-19 and severe hypoxia: an international, randomized, blinded trial": Suppl. 2

**eFigure 1.** Masking of Trial Medication

**eFigure 2.** Number of Enrolments per Site

**eFigure 3.** Number of Enrolments per Month

**eFigure 4.** Distributions of the Primary Outcome in the Post Hoc Analyses of the Primary Outcome Assigning Patients Who Had Died at Day 28 the Worst Possible Outcome (i.e., 0 Days Alive Without Life Support)

**eTables 1a and 1b.** Randomizations in Error and Wrongly Entered Stratification Variables

**eTable 2.** Trial Medication Administration and Protocol Violations

**eTable 3.** Results of the Analysis of the Primary Outcome in the Per Protocol Population

**eTable 4.** Best-Worst/Worst-Best Case Analyses of the Primary Outcome

**eTable 5.** Post Hoc Analyses of the Primary Outcome Assigning Patients Who had Died at Day 28 the Worst Possible Outcome (i.e., 0 Days Alive Without Life Support)

**eTable 6.** Analyses of the Single Components of Composite Outcomes

**eTable 7.** Results of the Unadjusted Analyses of Secondary Outcomes

**eTable 8.** All Serious Adverse Reactions and Serious Adverse Events

**eTable 9.** Use of Extracorporeal Membrane Oxygenation within 28 Days of Randomization

**eTable 10.** Discharge Against Medical Advice within 28 Days of Randomization

#### Contents

List of eFigures ... 3
List of eTables ... 4
eMethods ... 5
- The COVID STEROID 2 Trial Group ... 5
- Acknowledgements ... 11
- Inclusion Criteria ... 12
- Exclusion Criteria ... 13
- Baseline Characteristics ... 14
- Outcome definitions ... 17
- Criteria for Discontinuation of Trial Medication and Withdrawal ... 20
- Trial Populations ... 21
- Handling of Missing Data ... 22
- Abstract Written before Breaking the Randomization Code ... 24

eFigures ... 25
eTables ... 30
eReferences ... 40

Christian Hassager (Clinician), Manu Shankar-Hari (Trialist), and Susanne Rosthøj (Statistician).

**Site investigators and research staff**

The primary investigator at each site is marked with *.

Denmark

Department of Intensive Care, Copenhagen University Hospital – Rigshospitalet: Marie Warrer Munch*, Anders Perner, Maj-Brit Nørregaard Kjær, Morten Hylander Møller, Anders Granholm, Gitte Kingo Vesterlund, Tine Sylvest Meyhoff, Lene Russell, Kis Rønn Uhre, Ann Louise Syrach Lindgaard, Jette Degn, Mik Wetterslev, Praleene Sivapalan, Carl Thomas Anthon, Vibe Sommer Mikkelsen, Louise Colette la Porte, Marie Qvist Jensen, Jens Wolfgang Leistner, Camilla Meno Kristensen, Trine Bak Jonassen, Esben Christensen Clapp, Thomas Steen Jensen, Carl Johan Steensen Hjortsø, Matias Metcalf-Clausen, Reem Zaabalawi, Emilie Rose Bak, Liv Sanggaard Halstad, Suhayb Abdi, Emil Gleipner-Andersen, Emma Victoria Hatley, Tobias Saxtorph Aksnes.

Department of Infectious Diseases, Copenhagen University Hospital – Rigshospitalet: Marie Warrer Munch*, Marie Helleberg, Marie Qvist Jensen, Jens Wolfgang Leistner, Camilla Meno Kristensen, Trine Bak Jonassen, Esben Christensen Clapp, Thomas Steen Jensen, Carl Johan Steensen Hjortsø, Matias Metcalf-Clausen, Reem Zaabalawi, Emilie Rose Bak, Liv Sanggaard Halstad, Suhayb Abdi, Emil Gleipner-Andersen, Emma Victoria Hatley, Tobias Saxtorph Aksnes.

Department of Thoracic Anaesthesiology, Copenhagen University Hospital – Rigshospitalet: Marie Warrer Munch*, Vibeke Lind Jørgensen, Marie Qvist Jensen, Jens Wolfgang Leistner, Camilla Meno Kristensen, Trine Bak Jonassen, Esben Christensen Clapp, Thomas Steen Jensen, Carl Johan Steensen Hjortsø, Matias Metcalf-Clausen, Reem Zaabalawi, Emilie Rose Bak, Liv Sanggaard Halstad, Suhayb Abdi, Emil Gleipner-Andersen, Emma Victoria Hatley, Tobias Saxtorph Aksnes.

Department of Neuroanaesthesiology, Copenhagen University Hospital – Rigshospitalet: Marie Warrer Munch*, Margit Smitt, Marie Qvist Jensen, Jens Wolfgang Leistner, Camilla Meno Kristensen, Trine Bak Jonassen, Esben Christensen Clapp, Thomas Steen Jensen, Carl Johan Steensen Hjortsø, Matias Metcalf-Clausen, Reem Zaabalawi, Emilie Rose Bak, Liv Sanggaard Halstad, Suhayb Abdi, Emil Gleipner-Andersen, Emma Victoria Hatley, Tobias Saxtorph Aksnes.

Copenhagen Trial Unit, Centre for Clinical Intervention Research, The Capital Region, Copenhagen University Hospital – Rigshospitalet: Christian Gluud*, Janus Engstrøm.

Department of Infectious Diseases, Copenhagen University Hospital – Amager and Hvidovre: Thomas Benfield*, Charlotte Kastberg Levin, Gitte Kronborg, Rikke Krogh-Madsen, Marie Qvist Jensen, Jens Wolfgang Leistner, Camilla Meno Kristensen, Trine Bak Jonassen, Esben Christensen Clapp, Thomas Steen Jensen, Carl Johan Steensen Hjortsø, Matias Metcalf-Clausen, Reem Zaabalawi, Emilie Rose Bak, Liv Sanggaard Halstad, Suhayb Abdi, Emil Gleipner-Andersen, Emma Victoria Hatley, Tobias Saxtorph Aksnes.

Department of Intensive Care, Copenhagen University Hospital – Amager and Hvidovre: Thomas Benfield*, Klaus Tjelle Kristiansen, Ronni Thermann Reitz Plovsing, Marie Qvist Jensen, Jens Wolfgang Leistner, Camilla Meno Kristensen, Trine Bak Jonassen, Esben Christensen Clapp, Thomas Steen Jensen, Carl Johan Steensen Hjortsø, Matias Metcalf-Clausen, Reem Zaabalawi, Emilie Rose Bak, Liv Sanggaard Halstad, Suhayb Abdi, Emil Gleipner-Andersen, Emma Victoria Hatley, Tobias Saxtorph Aksnes.

Department of Respiratory Medicine, Copenhagen University Hospital – Amager and Hvidovre: Thomas Benfield*, Charlotte Suppli Ulrik, Marie Qvist Jensen, Jens Wolfgang Leistner, Camilla Meno Kristensen, Trine Bak Jonassen, Esben Christensen Clapp, Thomas Steen Jensen, Carl Johan Steensen Hjortsø, Matias Metcalf-Clausen, Reem Zaabalawi, Emilie Rose Bak, Liv Sanggaard Halstad, Suhayb Abdi, Emil Gleipner-Andersen, Emma Victoria Hatley, Tobias Saxtorph Aksnes.

Department of Anaesthesia and Intensive Care, Herlev Hospital: Anne Sofie Andreasen*, Helene Brix, Marie Qvist Jensen, Jens Wolfgang Leistner, Camilla Meno Kristensen, Trine Bak Jonassen,

Esben Christensen Clapp, Thomas Steen Jensen, Carl Johan Steensen Hjortsø, Matias Metcalf-Clausen, Reem Zaabalawi, Emilie Rose Bak, Liv Sanggaard Halstad, Suhayb Abdi, Emil Gleipner-Andersen, Emma Victoria Hatley, Tobias Saxtorph Aksnes.

Department of Anaesthesia and Intensive Care, Copenhagen University Hospital, North Zealand: Morten H. Bestle*, Martin Schønemann-Lund, Lone Valbjørn, Sanne Lauritzen, Ellen Bjerre Koch, Valdemar Oskar Ingemann Sørensen, Hans Eric Sebastian Seitz-Rasmussen, Frederik Heiberg Bestle.

Department of Anaesthesiology, Zealand University Hospital, Køge: Lone Musaeus Poulsen*, Nina Christine Andersen-Ranberg, Camilla Bekker Mortensen, Cecilie Bauer Derby, Kirstine Nanna la Cour, Sarah Weihe, Alison Holten Pind, Emma Ritsmer Stormholt.

Department of Anesthesia and Intensive Care, Slagelse Hospital, Slagelse: Anders Møller*, Klaus Vennick Marcussen*, Yifei Cao.

Dept. of Anaesthesia and Intensive Care, Odense University Hospital: Thomas Strøm*, Jens Michelsen, Eva Lærkner, Christine Gilberg, Michella Poulsen, Hanne Tanghus Olsen, Charlotte Rosenkilde.

Department of Anaesthesia and Intensive Care, Kolding Hospital: Anne Craveiro Brøchner*, Trine Haberlandt.

Department of Regional Health Research, Faculty of Health Sciences, University of Southern Denmark: Christian Gluud*.

Dept. of Anaesthesiology and Intensive Care, Bispebjerg and Frederiksberg Hospital: Christian Aage Wamberg*, Lars Peter Kloster Andersen, Diana Bertelsen Jensen, Mia Charlotte Krogh Hansen.

Department of Anaesthesia, Herning Hospital: Iben Strøm Darfelt*, Nilanjan Dey, Lene Friholdt Mahler.

Department of Anaesthesia and Intensive Care, Aalborg University Hospital: Bodil Steen Rasmussen*, Olav Lilleholt Schjørring, Søren Rosborg Aagaard, Stine Rom Vestergaard, Anne-Marie Gellert Bunzel, Rine Moulvad Siegumfeldt, Thomas Lass Klitgaard, Kirsten Uldal Krejberg.

Department of Infectious Diseases, Aalborg University Hospital: Bodil Steen Rasmussen*, Henrik Nielsen.

India

Department of Critical Care Medicine, Apollo Main Hospital, Chennai: Bharath Kumar Tirupakuzhi Vijayaraghavan*, Ajay Padmanaban*, Evangeline Elvira, Hilda Nirmala Kumari, Jayanthi Swaminathan, Arun Chander K, Ganesh Rajan N, Sona Thomas, Ann Thomas, Vishnu Odankadummal, Anu M Antony.

Department of Anaesthesia, Critical Care and Pain, Tata Memorial Hospital, Mumbai: Sheila Nainan Myatra*, Jigeeshu V. Divatia*, Atul P Kulkarni, Nishanth Baliga, Nirmalyo Lodh, Swapna C Vijayakumaran, Tarun Sahu, Kushal Kalvit, Vijaya P Patil, Amol T Kothekar, Shilpushp J Bhosale, Manoj Gorade, Anjana M Shrivastava.

Department of Intensive Care. P.D. Hinduja National Hospital & Medical Research Centre: Farhad N. Kapadia*. Jare Jagannath Uddhavrao, Ritika Sharma, Jyotsna Mali, Niranjan H R, Celine Lobo, Vaishali Neve, Deepika Maurya, Mahesh Salunke, Santiswaroop Pattanaik, Akhilesh Agrawal, Ashit Hegde, Rishi Kumar Badgurjar.

Department of Critical Care Medicine, Bombay Hospital Institute of Medical Sciences: Pravin R Amin*, Binit N Jhaveri, Digamber B Sarje, Ananda HJ, Bitan Sen, Ambily CM, Emilin P Jose.

Intensive Care Unit, S.L. Raheja Fortis Hospital, Mumbai: Sanjith Saseedharan*, Vaijayanti Kadam, Annapurna Chiluka, Rohit Kuril, Elizabeth Mathew, Suvarna Shirsekar.

Department of Respiratory Medicine, Indraprastha Apollo Hospital, New Delhi: Rajesh Chawla*, Sudha Kansal, Rinku Dahiya, Vijay Kumar Thakur, Rajni Tewatia, Savita Rawat, Harsh Tyagi.

Intensive Care Unit, Symbiosis University Hospital and Research Centre, Pune: Urvi Shukla*, Meenakshi Bhakare, Chinmayee Bhise, Amit Girme, Lini T. Kunjumon, Merlin Jose, Nikunj Sharma, Amol Bali.

Department of Critical Care Medicine, Sir HN Reliance Foundation Hospital: Mehul Shah*, Mayur Patel, Nirankar Bhutaka, Edwin Pathrose, Gopal Goyal, Riya Baby, Roshini Fernandez.

Department of Critical Care Medicine, Rajendra Institute of Medical Sciences: Mohd Saif Khan*, Pradip K Bhattacharya, Srishti Kindo, Aaditya A, Kritika Raj, Aastha Poddar, Aftab Ansari, Mustaque Alam.

Department of Internal Medicine, Vishwaraj Hospital, Pune: Kapil Borawake*, Vijay Khandagale, Satish Sarode, Sanesh Garde, Sushant Shinde ,Namdev Jagtap ,Chagan Khartode, Amit Palange, Aishwarya Pawar, Mayuri Mulay, Simrah Pirzade, Mohini Jagtap, Balbhim Rathod.

Department of Anaesthesiology, Gujarat Medical Education & Research Society (GMERS) Medical College and General Hospital, Gotri: Neeta Bose*, Dhara Tanna, Kinjal Chaudhary.

Department of Medicine, Gujarat Medical Education & Research Society (GMERS) Medical College and General Hospital, Gotri: Neeta Bose *, Shubhangi Deshpande, Rikin Raj.

Department of Critical Care, Sanjeevan Hospital: Subhal Dixit*, Sourab Ambapkar, Shobhana Ambapkar, Mukund Penurkar, Amit Sambare, Ravindra Joshi, Bhushan Kinolkar, Tarannum Aslam Shaikh.

Sweden

Department of Clinical Science and Education, Södersjukhuset, Karolinska Institutet: Rebecka Rubenson Wahlin*, Maria Cronhjort, Jacob Hollenberg, Anders Hedman, Anton Berggren, Felix Alarcón, Gabriel Yamin, Adam Heymowski.

Department of Anaesthesia and Intensive Care, Södersjukhuset: Rebecka Rubenson Wahlin*, Maria Cronhjort, Anton Berggren, Felix Alarcón, Gabriel Yamin, Adam Heymowski.

Department of Cardiology, Medical Intensive Care Unit, Södersjukhuset: Rebecka Rubenson Wahlin*, Jacob Hollenberg, Anders Hedman, Anton Berggren, Felix Alarcón, Gabriel Yamin, Adam Heymowski.

Department of Anaesthesia and Intensive Care, Universitetssjukhuset i Linköping: Michelle Chew*, Helen Didriksson, Carina Jönsson, Gunilla Gagnö.

Switzerland

Department of Intensive Care Medicine, Bern University Hospital (Inselspital): Luca Cioccari*, Stephan Jakob, David Zacharias, Margaret Lynn Jong, Marianne Roth, Rachelle Mader, Laureta Fazlija, Manuela Akaltan, Anja Eichenberger, Gion Rüegg, Tatjana Dill, Bruno Schoenmaekers.

#### Acknowledgements

We would like to thank all patients and relatives for agreeing to participate in the COVID STEROID 2 trial; all clinical and research staff at the participating hospitals; the regulatory authorities in the participating countries for the expedite handling of the protocol; and the funding sources.

We would also like to thank all clinical and research staff involved in the design and conduct of the 'Low-dose hydrocortisone in patients with Covid-19 and severe hypoxia (COVID STEROID) trial' which preceded the COVID STEROID 2 trial'.[1]

.

#### Inclusion Criteria

- Adult patients (aged 18 years or above at the time of randomization)
- Confirmed severe acute respiratory syndrome coronavirus 2 (SARS-CoV-2) requiring hospitalisation**:** we accepted any detections of SARS-CoV-2 approved by the local health authorities in the participating countries (i.e. pharyngeal swab, tracheal secretion or bronchoalveolar lavage).
- Use of either of the following:
  - Invasive mechanical ventilation: use of mechanical ventilation via a cuffed endotracheal tube **OR**
  - Non-invasive ventilation or continuous use of continuous positive airway pressure (CPAP) for hypoxia: Non-invasive ventilation includes positive pressure ventilation via a tight mask or helmet, continuous use of CPAP (via mask, helmet or tracheostomy, this does not include intermittent use of CPAP) **OR**
  - Oxygen supplementation with an oxygen flow ≥10 L/min irrespective of system used (mask or nasal cannula) or the addition of atmospheric air.

#### Exclusion Criteria

- Use of systemic corticosteroids for other indications than coronavirus disease 2019 (Covid-19) in doses higher than 6 mg dexamethasone equivalents: systemic corticosteroids (IV, IM, oral or per GI tube; not including nebulized, inhaled or transdermal corticosteroids) in doses higher than 6 mg dexamethasone / 6 mg betamethasone / 200 mg cortisone / 160 mg hydrocortisone / 32 mg methylprednisolone / 40 mg prednisolone / 40 mg prednisone. Other indications include adrenal insufficiency (i.e. primary, secondary or tertiary), anti-emetic treatment (i.e. post-operative or chemotherapy-induced nausea and vomiting), and immunosuppressive treatment (i.e. rheumatic diseases, allergic diseases, chronic obstructive pulmonary disease, haematological diseases, chronic kidney diseases, autoimmune hepatitis, inflammatory bowel disease, chronic neurological diseases).
- Use of systemic corticosteroids for Covid-19 for 5 days or more: use of systemic corticosteroids for Covid-19 for 5 consecutive preceding days or more up to the day of screening.
- Invasive fungal infection: any of the following:
    - Suspected invasive fungal infection: presence of plasma markers in blood (e.g. candida mannan antigen and galactomannan antigen).
    - Confirmed invasive fungal infection: positive culture from blood, peritoneal fluid or tissue.
- Active tuberculosis: either microbiologically confirmed or diagnosed based on epidemiological, clinical and radiographic data.
- Pregnancy: confirmed by positive urine human gonadotropin (hCG) or plasma-hCG. A negative urine or plasma-HCG test was mandatory for all women below 60 years of age.
- Known hypersensitivity to dexamethasone: history of any hypersensitivity reaction to dexamethasone, including but not limited to urticaria, eczema, angioedema, bronchospasm and anaphylaxis.
- Previously randomized in the COVID STEROID 2 trial: patients who had previously been randomized in the COVID STEROID 2 trial, either during current or former hospitalisation.
- Informed consent not obtainable: patients where the clinician or investigator was unable to obtain the necessary consent according to the national regulations, including patients with no relatives or patients who were hospitalized against their will.

#### Baseline Characteristics

- Sex: the genotypic sex of the participant.
- Age at enrolment: the age of the participant in whole years at the time of randomization. The age will be calculated from the date of birth and date of enrolment in the COVID STEROID 2 trial.
- Date of admission to hospital: the date of admission to the first hospital the participant was admitted to during the current hospital admission.
- Number of days with symptoms of COVID-19 before hospital admission: the total number of days (as integer) with symptoms of COVID-19 before hospital admission. If the symptoms started on the same day as hospital admission, the number of days with symptoms before hospital admission will be registered as 1. If the patient was infected while in hospital, the number of days with symptoms before hospital admission will be registered as 0.
- Department at which participant was included:
    - Emergency department: accident/emergency/casualty/acute department at COVID STEROID 2 trial sites.
    - Hospital ward: medical or surgical ward at COVID STEROID 2 trial site, including dedicated COVID-19 hospital wards.
    - Intermediate care unit: area of the hospital with higher resources to monitor patients as defined by the site, but invasive mechanical cannot be given.
    - Intensive care unit: area of the hospital where invasive mechanical can be given.
      Other: any location in the same or another hospital not covered in the other categories.
- Use of respiratory support at randomization:
    - Closed system (y/n): Use of invasive mechanical ventilation or non-invasive ventilation or continuous use of continuous positive airway pressure (CPAP) for hypoxia (defined in Supporting Information S2). If yes, latest $FiO_2$ prior to randomization.
    - Open system with an oxygen flow ≥10 L/min: defined in Supporting Information S2. If yes, the maximum supplemental oxygen flow on an open system at randomization (+/- 1 h) will be registered.
- Limitations of care (y/n): participant with limitation(s) in use of life support (i.e. invasive mechanical ventilation, circulatory support, renal replacement therapy) and/or cardio-pulmonary resuscitation at the time of randomization.

#### Outcome definitions

##### Primary outcome

The days alive without life support at day 28 was calculated as the total number of days alive without any of the 3 life supporting interventions (listed below) within 28 days after randomization. Use of life support was defined as the need for:

- Invasive mechanical ventilation: use of mechanical ventilation via a cuffed endotracheal tube.
- Circulatory support: infusion of any vasopressor/inotrope agent for a minimum of 1 hour (i.e. norepinephrine, epinephrine, phenylephrine, vasopressin analogues, angiotensin, dopamine, dobutamine, milrinone or levosimendan).
- Renal replacement therapy: any form of acute or chronic intermittent or continuous renal replacement therapy (RRT), including days between intermittent RRT. We defined periods with up to 3 days between intermittent RRT as days with RRT (e.g. if a patient received RRT on day 1, did not receive RRT on days 2-4, and received RRT again on day 5, the patient was considered on RRT on days 1-5).

##### Secondary outcome

1. Number of patients with one or more serious adverse reactions (SARs) at day 28 was assessed as at least one of the following on each day from randomization to day 28:

    - New episodes of septic shock: we defined septic shock according to the Sepsis-3 criteria:[2]

        - Suspected or confirmed superinfection.
        - New infusion (or 50% increase) of vasopressor/inotropic agent (i.e. norepinephrine, epinephrine, phenylephrine, vasopressin analogues, angiotensin, dopamine, dobutamine, milrinone or levosimendan) to maintain a mean arterial blood pressure of 65 mmHg or above.
        - Lactate of 2 mmol/L or above in any plasma sample performed on the same day

    - Invasive fungal infection: any of the following:

- Suspected invasive fungal infection: presence of plasma markers in blood (e.g. candida mannan antigen and galactomannan antigen).
- Confirmed invasive fungal infection: positive culture from blood, peritoneal fluid or tissue.

- Clinically important gastrointestinal (GI) bleeding was assessed as any GI bleeding **AND** the use of at least 2 units of red blood cells on the same day. GI bleeding defined as hematemesis, coffee ground emesis, melena, haematochezia or bloody nasogastric aspirate on this day.
- Anaphylactic reaction to IV dexamethasone: anaphylactic reactions was only recorded during the intervention period (i.e. for up to 10 days depending on the number of days with corticosteroid use before randomization). An anaphylactic reactions was defined as urticarial skin reaction **AND** at least one of the following observed after randomization
    - Worsened circulation (>20% decrease in blood pressure or >20% increase in vasopressor dose).
    - Increased airway resistance (>20% increase in the peak pressure on the ventilation)
    - Clinical stridor or bronchospasm.
    - Subsequent treatment with bronchodilators.

2. All-cause mortality at day 28 after randomization: death from any cause within 28 days post-randomization.

The following secondary longer-term outcomes are also registered in the COVID STEROID 2 trial[3] but will be reported later when follow-up has completed.

1. All-cause mortality at day 90 after randomization.
2. Days alive without life support (i.e. invasive mechanical ventilation, circulatory support or renal replacement therapy) at day 90.
3. Days alive and out of hospital at day 90.
4. All-cause mortality at 180 days after randomization.
5. Health-Related Quality of Life (HRQoL) at 180 days after randomization assessed by EQ-5D-5L.

6. Health-Related Quality of Life (HRQoL) at 180 days after randomization assessed by EQ-VAS.

#### Criteria for Discontinuation of Trial Medication and Withdrawal

The procedure for handling withdrawal of consent from a patient followed national regulations.

**Discontinuation and withdrawal at the choice of the patient or the proxy**

The patient or next of kin could withdraw his/her consent or proxy consent at any time without further explanations and without consequences for further treatment. In these instances, the investigator asked the patient or next of kin if they allowed continued data registration and follow-up until day 180. Already collected data up until the time of withdrawal were used according to national regulations.

If the patient was withdrawn before the first dose of trial medication was administered, the patient was not considered a part of the intention-to-treat (ITT) population. If the patient was withdrawn after one or more doses of trial medication were administered, and the patient or next of kin allowed continued data registration, the patient remained in the ITT population. If the patient was withdrawn after one or more doses of trial medication were administered, and the patient or next of kin did not allow continued data registration, the patient remained in the ITT population but with missing data from the time of withdrawal. For patients withdrawn in Switzerland due to withdrawal of consent, all registered data were deleted, including the data registered before the time of withdrawal as required by the Swiss Ethics Committee. Any such Swiss patient was not part of the ITT population.

**Discontinuation and withdrawal at the choice of the investigator**

A patient could have the intervention stopped by the clinician or investigator at any time if:

- The patient experienced intolerable adverse reactions or events (including SARs or suspected unexpected serious adverse reactions (SUSARs)) suspected to be related to the trial intervention.
- The clinicians in conjunction with the coordinating investigator decided it to be in the interest of the patient.
- The patient was withdrawn from active therapy.
- The patient was subject to compulsory hospitalisation.

In these patients, the collection of data and the follow-up continued, and the patient remained in the ITT population.

#### Trial Populations

##### Definition of the ITT population

The ITT population was defined as all randomized patients and for whom there was consent for the use of data.

##### Definition of the per-protocol population

The per-protocol population was defined as the ITT population except those having one or more major protocol violations during the intervention period (for up to 10 days depending on the number of consecutive days with corticosteroid use before randomization) defined as:

- Use of open-label systemic corticosteroids.
- Trial medication not administered as per protocol (1 bolus injection of either 12 mg or 6 mg dexamethasone according to the allocation on each day during the intervention period).

Patients who discontinued trial medication due to withdrawal of consent or for clinical reasons were considered as part of the per-protocol population.

#### Handling of Missing Data

##### Complete missingness of data

We had complete missingness of data for 5 patients who withdrew consent and had all registered data deleted, including the data registered before the time of withdrawal as required by the Swiss Ethics Committee.

##### Missing baseline variables

We had missing source data for the time from onset of symptoms to hospital admission for 32 patients allocated to dexamethasone 12 mg and for 18 patients allocated to dexamethasone 6 mg; for the fraction of inspired oxygen for patients receiving invasive mechanical ventilation for 1 patient allocated to dexamethasone 12 mg; for the fraction of inspired oxygen for patients receiving non-invasive ventilation or continuous use of CPAP for 4 patients allocated to dexamethasone 12 mg and for 8 patients allocated to dexamethasone 6 mg; for the most recent partial pressure of oxygen (PaO2) prior to randomization for 28 patients allocated to dexamethasone 12 mg and for 23 patients allocated to dexamethasone 6 mg; for the most recent arterial oxygen saturation prior to randomization for 5 patients allocated to dexamethasone 12 mg and for 9 patients allocated to dexamethasone 6 mg; and for the highest plasma lactate value in the last 24 hours prior to randomization for 57 patients allocated to dexamethasone 12 mg and for 49 patients allocated to dexamethasone 6 mg. The missing data in these patients were not included in the baseline characteristics.

##### Missing outcome data

Primary outcome

We had data for the primary outcome for 971 of 982 (98.9%) patients in the ITT population (98.8% in patients allocated to dexamethasone 12 mg; 99.0% in patients allocated to dexamethasone 6 mg). 11 patients had missing data due to withdrawal of consent from the time of withdrawal; no patients were lost to follow-up.

Secondary outcomes

We had data for 28-day mortality for 971 of 982 (98.9%) patients in the ITT population (98.8% in patients allocated to dexamethasone 12 mg; 99.0% in patients allocated to dexamethasone 6 mg). 11 patients had missing data due to withdrawal of consent from the time of withdrawal; no patients were lost to follow-up.

We analyzed data for SARs for all 982 patients in the ITT population. For the 11 patients who withdrew consent before 28-days follow up, the occurrence of serious adverse reactions was analyzed until the day of withdrawal of consent

**Logical imputation**

We used logical imputation for one patient that likely left the country of enrolment before 28-days follow-up. The patient was last seen alive 3 days before 28-days follow up, where the patient was assessed to be fit-to-fly by a medical doctor. Consequently, the patient was assumed to be alive, without the need for life support, and without any serious adverse reactions after discharge and until day 28 after randomization.

We used logical imputation for one patient for whom the relatives could inform that the patient had received renal replacement therapy after discharge, but not the number of days with renal replacement therapy after discharge. The patient died on day 46 after randomization. For this patient, we assumed that the patient received renal replacement therapy (including days in between intermittent renal replacement therapy) at all days from discharge to 28-days follow-up.

#### Abstract Written before Breaking the Randomization Code

#### v. 1.3, June 26, 2021, approved by the Management Committee

**Background**

Dexamethasone 6 mg daily is recommended for up to 10 days in patients with severe and critical Covid-19. A higher dose of dexamethasone may benefit those with more severe disease.

**Methods**

In this international, blinded trial, we randomly assigned adults with confirmed Covid-19 receiving at least 10 L/min of oxygen or mechanical ventilation to intravenous dexamethasone 12 mg or dexamethasone 6 mg daily for up to 10 days. The primary outcome was the number of days alive without life support (i.e. invasive mechanical ventilation, circulatory support or renal replacement therapy) at 28 days.

**Results**

We randomized 1000 patients, included 982 in the intention-to-treat-population and had primary outcome data for 971, among whom 491 patients were assigned to dexamethasone A mg and 480 patients assigned to dexamethasone B mg. The median number of days alive without life support at 28 days was 22.0 days (interquartile range 6.0-28.0) in the A mg group and 20.5 days (4.0-28.0) in the B mg group (adjusted mean difference 1.3 days, 95% confidence interval (CI), 0.0-2.6; P=0.066). The 28-day mortality was 27.1% and 32.3% in patients assigned to A mg and B mg, respectively, (adjusted relative risk 0.86, 99% CI, 0.68-1.09). The 28-day occurrence of serious adverse reactions were similar in the two groups (adjusted relative risk 0.85, 99% CI, 0.56-1.31).

**Conclusions**

Among patients with Covid-19 and severe hypoxia, dexamethasone A mg probably led to more days alive without life support at 28 days than dexamethasone B mg.

**Note, 28 June 2021.** The upper confidence interval for 28-day mortality differs between the abstract written before breaking the randomization code and the final one due to simulation uncertainty and rounding; these estimates were calculated using logistic regression and g-computation, which involved a bootstrap resampling procedure to estimate confidence intervals, with 5,000 samples used in the analyses conducted before unblinding and 50,000 samples used in the final analyses.

**Note, 21 July 2021.** Four serious adverse reactions were refuted when reviewed before final reporting to institutional ethics committees. The adjusted relative risk therefore differs between the abstract written before breaking the randomization code and the final one.

### eFigures

#### eFigure 1. Masking of Trial Medication

We used shelf-medications from the hospital department’s pharmacy for the trial interventions. To ensure blinding, the trial medications were prepared by a team of unblinded trial staff; all others, including the patients and clinical staff, remained blinded to the treatment allocation. For each patient, the trial medication was be prepared once daily and administered as a bolus injection intravenously.

Dexamethasone phosphate (used at all hospitals expect in the one Swedish hospital where bethamethasone phosphate was used) is a clear colourless solution. To ensure blinding, the dexamethasone phosphate solution was mixed with isotonic saline (0.9%) to a bolus volume of 5 ml for both trial interventions. Consequently, the 12 mg and 6 mg dexamethasone bolus injections appeared identical upon visual inspection (eFigure 1). The preparation of trial medication is described in detail in the Supporting Information S5 in the primary protocol.[3]

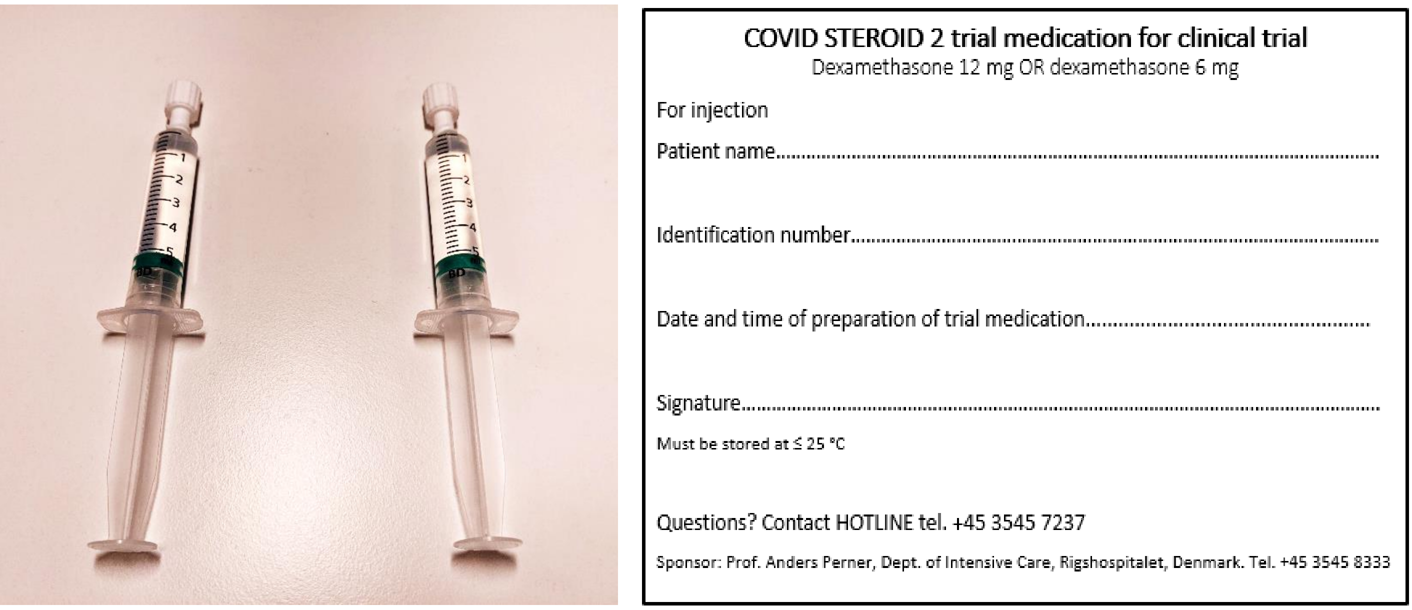

**eFigure 1. Masking of trial medication.** Trial interventions (12 mg and 6 mg of dexamethasone) without labels (to the left). Both interventions were labelled with identical labels (to the right).

**Local trade names**

The local trade names used in the COVID STEROID 2 trial are listed below.

Denmark

Dexavit™, Vital Pharma Nordic, Denmark, ATC code: H02AB02.

Sweden

Betapred™, Alfasigma S.p.A., Italy, ATC code: H02AB01.
Dexavit™, Vital Pharma Nordic, Denmark, ATC code: H02AB02.

Switzerland

Mephameson™, Mepha Pharma AG, Switzerland, ATC code: H02AB02.

India

Daksone™, Daksh Pharmaceuticals Pvt. Ltd, India, ATC code: H02AB02.
Dacdac™, Wockhardt Limited (Merind), India, ATC code: H02AB02.
Decmax™, GLS Pharma Ltd., India, ATC code: H02AB02.
Demisone™, Cadila Pharmaceuticals Ltd. (Genvista), India, ATC code: H02AB02.
Dexona™, Zydus Cadila Healthcare Ltd. (Alidac), India, ATC code: H02AB02.
Dex-V™, Vensat Bio, India, ATC code: H02AB02.
Intradex™, Intra Labs India Pvt. Ltd, India, ATC code: H02AB02.

**eFigure 2. Number of Enrolments per Site**

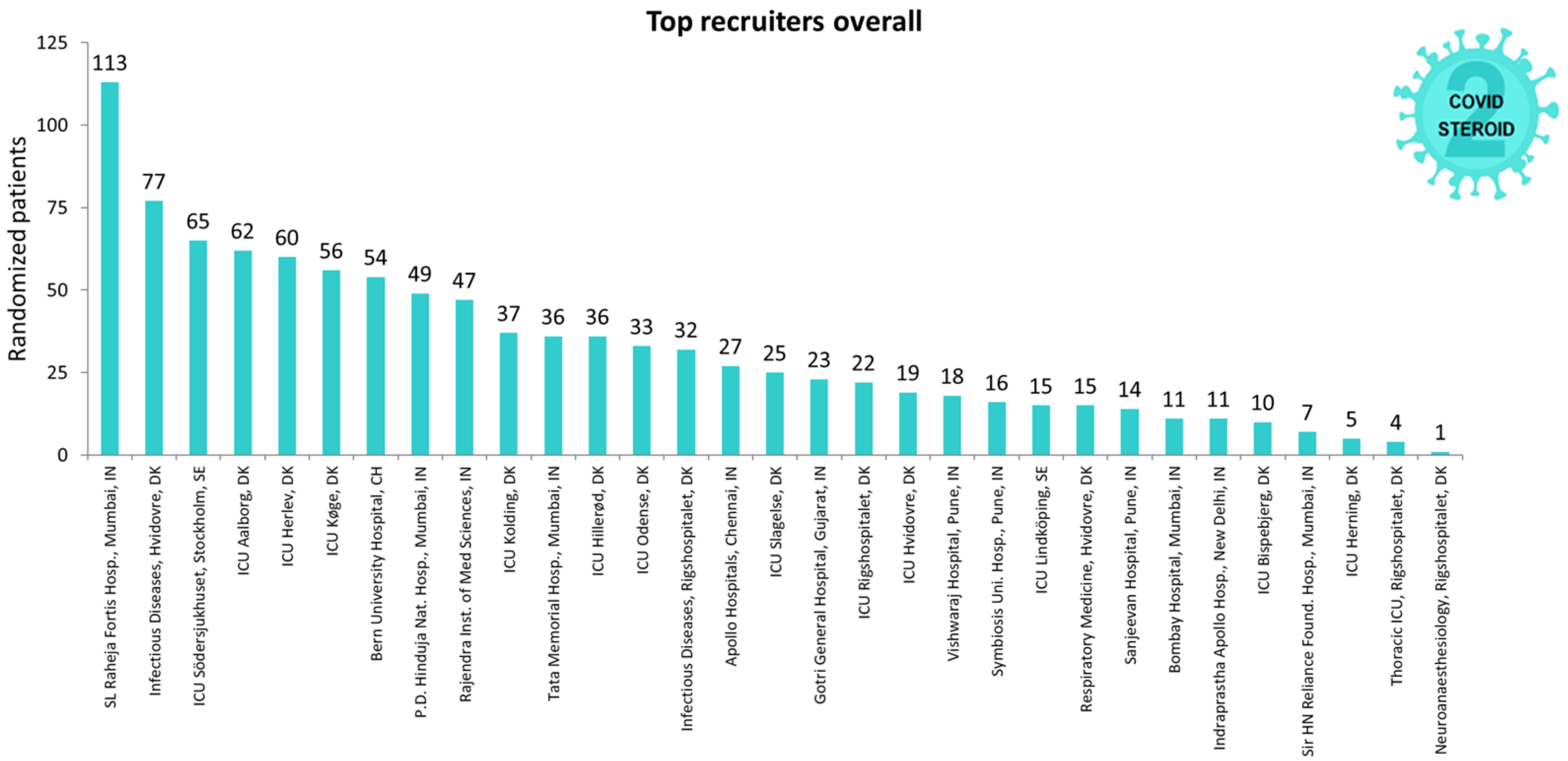

The 4 sites at Rigshospitalet, Denmark, was run by one research staff team; so was the 3 sites at Hvidovre Hospital, Denmark.

#### eFigure 3. Number of Enrolments per Month

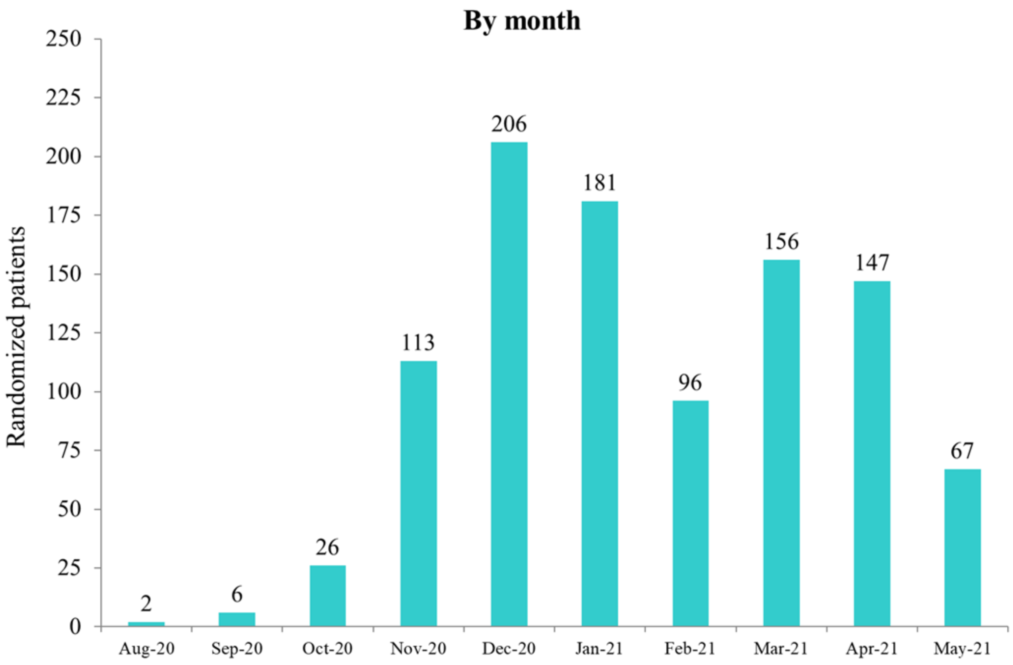

**eFigure 4. Distributions of the Primary Outcome in the Post Hoc Analyses of the Primary Outcome Assigning Patients Who had Died at Day 28 the Worst Possible Outcome (i.e., 0 Days Alive Without Life Support)**

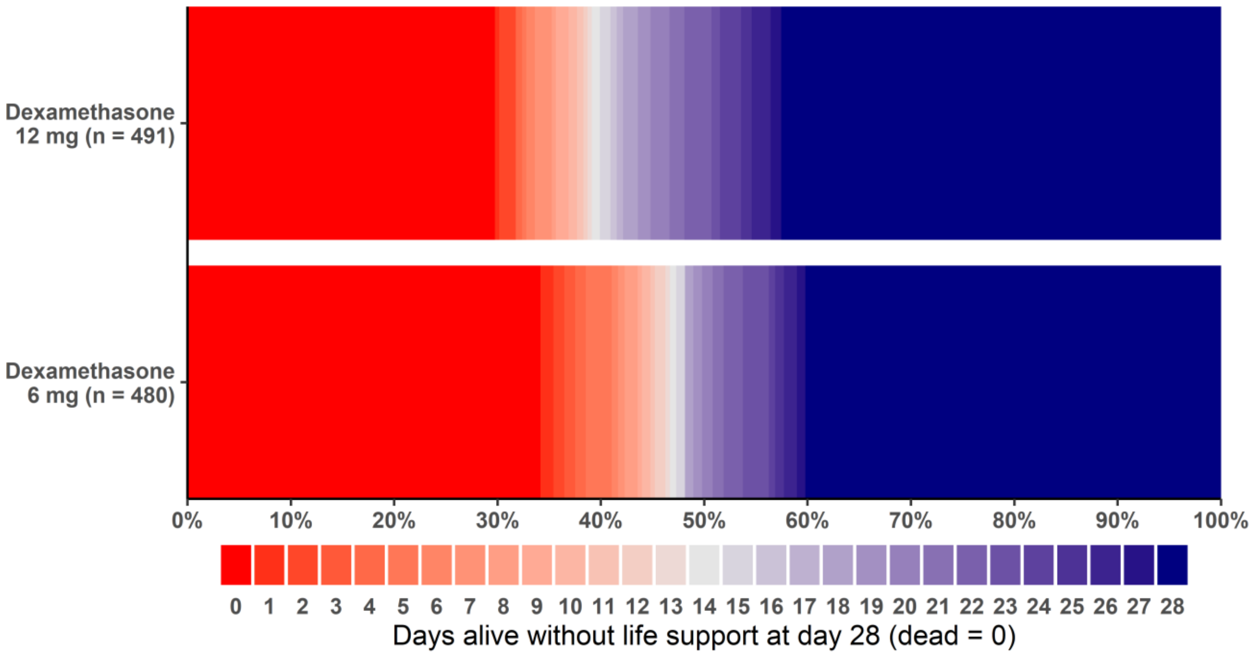

eFigure 4 shows the number of days alive without life support as horizontally stacked proportions in the two intervention groups in the intention-to-treat population in the *post hoc* analysis assigning patients who had died at day 28 the worst possible outcome of 0 days alive without life support at day 28 after randomization (n=971; 11 patients had missing primary outcome data). Red represents worse outcomes, and blue represents better outcomes.

### eTables

#### eTable 1a and 1b. Randomizations in Error and Wrongly Entered Stratification Variables

##### eTable 1a: Randomizations in error

| Reason for randomization in error | Number of patients |
|---|---|
| Ineligible at the time of randomization[a] | 8 |
| Did not fulfill inclusion criteria[b] | 4 |
| Fulfilled one or more exclusion criteria[c] | 4 |
| Duplicate randomizations[d] | 1 |

[a] All 8 patients were included in the ITT population.
[b] 3 patients received less than 10 L/min of supplementary oxygen and 1 was PCR-negative for SARS-CoV-2.
[c] 3 had received corticosteroids for Covid-19 for 5 consecutive days or more, and 1 had an indication for systemic corticosteroids in a dose higher than dexamethasone 6 mg or equivalent.
[d] The patient received trial medication according to the allocation at the first randomization and was only included once in the ITT population.

##### eTable 1b: Wrongly entered stratification variables

| Stratification variable[a] | Number of patients |
|---|---|
| Site[b] | 6 |
| Invasive mechanical ventilation | 7 |
| Age below 70 years | 18 |

[a] The wrongly entered stratification variables were corrected for all these 31 patients before the adjusted analyses were conducted.
[b] Some investigators had access to more than one site in the electronic case report form where screening was performed. As a result, 6 patients were administratively allocated at a wrong site.

#### eTable 2. Trial Medication Administration and Protocol Violations

| | Dexamethasone 12 mg (n=497) | Dexamethasone 6 mg (n=485) |
|---|---|---|
| **Trial medication administration** | | |
| Treatment duration, median (IQR), days | 7 (5 - 9) | 7 (6 - 9) |
| **Major protocol violation, no. (%)** | 44 (8.9%) | 46 (9.5%) |
| Use of open-label systemic corticosteroids | 10 (2.0%) | 9 (1.9%) |
| Trial medication not administered as per protocol | 36 (7.2%) | 39 (8.0%) |

Abbreviations: mg, milligrams; IQR, interquartile range.

#### eTable 3. Results of the Analysis of the Primary Outcome in the Per Protocol Population[a]

| Primary outcome | Dexamethasone 12 mg (N=453) | Dexamethasone 6 mg (N=439) | Mean difference (95% CI)[b] | Median difference (95% CI)[b] |
|---|---|---|---|---|
| Number of days alive without life support, median (IQR)[c] | 23.0 (6.0-28.0) | 20.0 (4.0-28.0) | 1.3 (0.0 to 2.7) | 0 (-0.9 to 0.9) |

Abbreviations: mg, milligrams; CI, confidence interval; IQR, interquartile range.

[a] Defined as the ITT population except those having one or more major protocol violations (i.e. use of open-label systemic corticosteroids and/or trial medication not administered as per protocol) during the intervention period (N = 892).

[b] Significance level is set to 5% including the interim analysis. Accordingly, we have employed 95.08% confidence intervals for the final analysis.

[c] Analyzed using the Kryger Jensen and Lange test adjusted for the stratification variables (site, invasive mechanical ventilation, and age below 70 years).

#### eTable 4. Best-Worst/Worst-Best Case Analyses of the Primary Outcome

| | Dexamethasone 12 mg (N=497) | Dexamethasone 6 mg (N=485) | Mean difference (95% CI)[a] | Median difference (95% CI)[a] |
|---|---|---|---|---|
| **Best-worst case analysis[b]** | | | | |
| Number of days alive without life support, median (IQR) | 22.0 (6.0-28.0) | 20.0 (4.0-28.0) | 1.6 (0.3 to 2.9) | 0 (-0.8 to 0.8) |
| **Worst-best case analysis[c]** | | | | |
| Number of days alive without life support, median (IQR) | 22.0 (5.0-28.0) | 21.0 (4.0-28.0) | 1.1 (-0.2 to 2.4) | 0 (-0.5 to 0.5) |

Abbreviations: mg, milligrams; CI, confidence interval; IQR, interquartile range.
[a] Significance level is set to 5% including the interim analysis. Accordingly, we have employed 95.08% confidence intervals for the final analysis. Analyzed using the Kryger Jensen and Lange test adjusted for the stratification variables (site, invasive mechanical ventilation, and age below 70 years).
[b] Analyzed assuming all patients lost to follow-up in the 12 mg group have had a beneficial outcome (i.e. assuming that the patient was alive and without life support for all days with missing data) and all patients lost to follow-up in the 6 mg group have had a harmful outcome (i.e. assuming that the patient was dead and/or received life support on all days with missing data).
[c] Analyzed assuming all patients lost to follow-up in the 12 mg group have had a harmful outcome ((i.e. assuming that the patient was dead and/or received life support on all days with missing data) and all patients lost to follow-up in the 6 mg group have had a beneficial outcome (i.e. assuming that the patient was alive and without life support for all days with missing data).

#### eTable 5. *Post Hoc* Analyses of the Primary Outcome Assigning Patients Who Had Died at Day 28 the Worst Possible Outcome (i.e., 0 Days Alive Without Life Support)

| **Outcome** | **Dexamethasone 12 mg (N=497)** | **Dexamethasone 6 mg (N=485)** | **Mean difference (95% CI)[a]** | **Median difference (95% CI)[a]** |
|---|---|---|---|---|
| **Primary outcome[b]** | | | | |
| Number of days alive without life support, median (IQR) | 22 (0 - 28) | 20 (0 - 28) | 1.5 (0.1 to 3.0) | 0 (-1.1 to 1.1) |
| Number of days alive without invasive mechanical ventilation, median (IQR) | 23 (0 - 28) | 21 (0 - 28) | | |
| Number of days alive without circulatory support, median (IQR) | 26 (0 - 28) | 25 (0 - 28) | | |
| Number of days alive without renal replacement therapy, median (IQR)[c] | 28 (0 - 28) | 28 (0 - 28) | | |

Abbreviations: mg, milligrams; CI, confidence interval; IQR, interquartile range.
Before the lock of the trial database, we decided to do this *post hoc* sensitivity analysis assigning dead patients the worst possible outcome (i.e. 0 days alive without life support) as this as previously been proposed and is frequently used in other, similar trials.[4,5]

[a] Analyzed using the Kryger Jensen and Lange test adjusted for the stratification variables (site, invasive mechanical ventilation, and age below 70 years. Significance level is set to 5% including the interim analysis for the primary outcome. Accordingly, we have employed 95.08% confidence interval for the primary outcome.

[b] Defined as days alive and without the use of invasive mechanical ventilation, circulatory support or renal replacement therapy. Data were missing for 6 patients assigned to dexamethasone 12 mg and 5 patients assigned to dexamethasone 6 mg.

[c] Any form of acute or chronic intermittent or continuous renal replacement therapy, including days between intermittent dialysis (i.e. we have considered up to 3 days in between renal replacement therapy as days with renal placement therapy).

#### eTable 6. Analyses of the Single Components of Composite Outcomes

| Outcomes at day 28 | Dexamethasone 12 mg (N=497) | Dexamethasone 6 mg (N=485) | Mean difference (95% CI)[a] | Median difference (95% CI)[a] |
|---|---|---|---|---|
| **Primary outcome: days alive without life support[b]** | | | | |
| Number of days alive without invasive mechanical ventilation, median (IQR) | 23 (7 - 28) | 22 (5 - 28) | 1.0 (-0.3 to 2.2) | 0 (-0.5 to 0.5) |
| Number of days alive without circulatory support, median (IQR) | 26 (13 - 28) | 25 (9 - 28) | 1.0 (-0.2 to 2.2) | 0 (-0.1 to 0.1) |
| Number of days alive without renal replacement therapy, median (IQR)[c] | 28 (18 - 28) | 28 (14 - 28) | 0.7 (-0.4 to 1.8) | -[d] |
| **Secondary outcome: serious adverse reactions, no. (%)[e]** | | | **Relative risk (99% CI)[a]** | **Risk difference (99% CI)[a]** |
| New episodes of septic shock | 42 (8.5%) | 49 (10.1%) | 0.85 (0.50 to 1.43) | -1.5 (-6.2 to 3.2) |
| Invasive fungal infection | 15 (3.0%) | 21 (4.3%) | 0.70 (0.27 to 1.64) | -1.3 (-4.2 to 1.7) |
| Clinically important gastrointestinal bleeding | 9 (1.8%) | 5 (1.0%) | -[f] | -[f] |
| Anaphylactic reaction to intravenous dexamethasone[g] | 0 (0.0%) | 0 (0.0%) | - | - |

Abbreviations: mg, milligrams; CI, confidence interval; IQR, interquartile range.
[a] Significance level is set to 5% including the interim analysis. Accordingly, we have employed 95.08% confidence interval for the primary outcome, and 99% confidence intervals for the secondary outcomes. Risk differences are reported in percentage points.
[b] Analyzed using the Kryger Jensen and Lange test adjusted for the stratification variables (site, invasive mechanical ventilation, and age below 70 years). Data were missing for 6 patients assigned to dexamethasone 12 mg and 5 patients assigned to dexamethasone 6 mg.
[c] Any form of acute or chronic intermittent or continuous renal replacement therapy, including days between intermittent dialysis (i.e. we have considered up to 3 days in between renal replacement therapy as days with renal placement therapy).
[d] Not applicable as too many observations are identical.
[e] Analyzed using logistic regression and g-computation adjusted for the stratification variables (site, invasive mechanical ventilation, and age below 70 years).
[f] Too few events to be meaningfully analyzed.
[g] Only recorded during the intervention period (for up to 10 days).

#### eTable 7. Results of the Unadjusted Analyses of Secondary Outcomes

| Secondary outcomes[a] | Relative risk (99% CI) | Risk difference (99% CI) |
|---|---|---|
| Death by day 28 | 0.84 (0.65 to 1.08) | -5.2 (-12.7 to 2.3) |
| Patients with one or more serious adverse reactions[b] | 0.85 (0.61 to 1.19) | -1.9 (-6.1 to 2.2) |

Abbreviations: CI, confidence interval.
[a] Data were missing for 6 patients assigned to dexamethasone 12 mg and 5 patients assigned to dexamethasone 6 mg. Both outcomes are analyzed using generalized linear models with log/identity links and binomial error distributions. Risk differences are reported in percentage points.
[b] Defined as new episodes of septic shock, invasive fungal infection, clinically important gastrointestinal bleeding or anaphylactic reaction to intravenous dexamethasone.

#### eTable 8. All Serious Adverse Reactions and Serious Adverse Events[a]

| | Dexamethasone 12 mg (N=497) | Dexamethasone 6 mg (N=485) |
|---|---|---|
| **SARs registered in addition to those in the SAR-outcome, no. (%)[b]** | | |
| Sepsis | 2 (0.4%) | 5 (1.0%) |
| Bacteraemia | 4 (0.8%) | 1 (0.2%) |
| Pneumonia | 6 (1.2%) | 7 (1.4%) |
| *C. difficile* | 0 (0.0%) | 1 (0.2%) |
| Other | 3 (0.6%) | 3 (0.6%) |
| Total[c] | 14 (2.8%) | 17 (3.5%) |
| **SAEs, no. (%)[d]** | | |
| Thromboembolic events | 4 (0.8%) | 10 (2.1%) |
| Bleeding | 7 (1.4%) | 7 (1.4%) |
| Acute kidney injury | 52 (10.5%) | 62 (12.8%) |
| Pulmonary complications, including pneumothorax | 3 (0.6%) | 3 (0.6%) |
| Circulatory failure, including cardiac arrest | 9 (1.8%) | 9 (1.9%) |
| Other | 7 (1.4%) | 3 (0.6%) |
| Total[e] | 70 (14.1%) | 85 (17.5%) |
| **Patients with one or more SARs or SAEs, no. (%)[f]** | 108 (21.7%) | 130 (26.8%) |

Abbreviations: mg, milligrams; SAE, serious adverse event; SAR, serious adverse reaction.

[a] SAE and any SAR (both those included in the SAR-outcome (i.e. new episodes of septic shock, invasive fungal infection, clinically important gastrointestinal bleeding, or anaphylactic reaction to intravenous dexamethasone) and those reported directly to Sponsor or his delegate by e-mail or using a standardized report form).

[b] Other registered SARs, i.e. adverse reactions registered in the Summary Product Characteristic for dexamethasone considered serious and not registered in the secondary SAR outcome.

[c] Total number of patients with at least one SAR not included in the secondary SAR outcome.

[d] Any SAE not considered a SAR.

[e] Total number of patients with at least one SAE.

[f] Including the reactions reported in the secondary SAR outcome.

#### eTable 9. Use of Extracorporeal Membrane Oxygenation within 28 Days of Randomization[a]

| | Dexamethasone 12 mg | Dexamethasone 6 mg |
|---|---|---|
| Extracorporeal membrane oxygenation, no. (%) | 3 (0.6%) | 14 (2.9%) |

Abbreviations: mg, milligrams.
[a] Data were missing for 6 patients assigned to dexamethasone 12 mg and 5 patients assigned to dexamethasone 6 mg.

#### eTable 10. Discharge Against Medical Advice within 28 Days of Randomization

| | Dexamethasone 12 mg | Dexamethasone 6 mg |
|---|---|---|
| Discharged against medical advice, no. (%) | 9 (1.8%) | 11 (2.3%) |
| Use of life support at the time of discharge | 1 (0.2%) | 1 (0.2%) |
| Invasive mechanical ventilation | 1 (0.2%) | 1 (0.2%) |
| Circulatory support | 0 (0.0 %) | 0 (0.0 %) |
| Renal replacement therapy[a] | 0 (0.0 %) | 0 (0.0 %) |
| Use of supplementary oxygen at the time of discharge | 5 (1.0%) | 8 (1.7%) |
| ≥ 10 l/min | 2 (0.4%) | 3 (0.6%) |
| 0-9 l/min | 3 (0.6%) | 5 (1.0%) |

Abbreviations: mg, milligrams; l, liters; min, minutes.
[a] Any form of acute or chronic intermittent or continuous renal replacement therapy, including days between intermittent dialysis (i.e. we have considered up to 3 days in between renal replacement therapy as days with renal placement therapy).
